# Genome-wide association study identifies new locus associated with OCD

**DOI:** 10.1101/2021.10.13.21261078

**Authors:** Nora I. Strom, Dongmei Yu, Zachary F. Gerring, Matthew W. Halvorsen, Abdel Abdellaoui, Cristina Rodriguez-Fontenla, Julia M. Sealock, Tim Bigdeli, Jonathan R. I. Coleman, Behrang Mahjani, Jackson G. Thorp, Katharina Bey, Christie L. Burton, Jurjen J. Luykx, Gwyneth Zai, Kathleen D. Askland, Cristina Barlassina, Judith Becker Nissen, Laura Bellodi, O. Joseph Bienvenu, Donald Black, Michael Bloch, Julia Boberg, Rosa Bosch, Michael Breen, Brian P. Brennan, Helena Brentani, Joseph D. Buxbaum, Jonas Bybjerg-Grauholm, Enda M. Byrne, Beatriz Camarena, Adrian Camarena, Carolina Cappi, Angel Carracedo, Miguel Casas, Maria C. Cavallini, Valentina Ciullo, Edwin H. Cook, Vladimir Coric, Bernadette A. Cullen, Elles J. De Schipper, Bernie Devlin, Srdjan Djurovic, Jason A. Elias, Lauren Erdman, Xavier Estivil, Martha J. Falkenstein, Bengt T. Fundin, Maiken E. Gabrielsen, Fernando S. Goes, Marco A. Grados, Jakob Grove, Wei Guo, Jan Haavik, Kristen Hagen, Alexandra Havdahl, Ana G. Hounie, Donald Hucks, Christina Hultman, Magdalena Janecka, Michael Jenike, Elinor K. Karlsson, Julia Klawohn, Lambertus Klei, Janice Krasnow, Kristi Krebs, Jason Krompinger, Nuria Lanzagorta, Fabio Macciardi, Brion Maher, Evonne McArthur, Nathaniel McGregor, Nicole C. McLaughlin, Sandra Meier, Euripedes C. Miguel, Maureen Mulhern, Paul S. Nestadt, Erika L. Nurmi, Kevin S. O’Connell, Lisa Osiecki, Teemu Palviainen, Fabrizio Piras, Federica Piras, Ann E. Pulver, Raquel Rabionet, Alfredo Ramirez, Scott Rauch, Abraham Reichenberg, Jennifer Reichert, Mark A. Riddle, Stephan Ripke, Aline S. Sampaio, Miriam A. Schiele, Laura G. Sloofman, Jan Smit, Janet L. Sobell, María Soler Artigas, Laurent F. Thomas, Homero Vallada, Jeremy Veenstra-VanderWeele, Nienke N. C. C. Vulink, Christopher P. Walker, Ying Wang, Jens R. Wendland, Bendik S. Winsvold, Yin Yao, Pino Alonso, Götz Berberich, Cynthia M. Bulik, Danielle Cath, Daniele Cusi, Richard Delorme, Damiaan Denys, Valsamma Eapen, Peter Falkai, Thomas V. Fernandez, Abby J. Fyer, Daniel A. Geller, Hans J. Grabe, Benjamin D. Greenberg, Gregory L. Hanna, Ian M. Hickie, David M. Hougaard, Norbert Kathmann, James Kennedy, Liang Kung-Yee, Mikael Landén, Stéphanie Le Hellard, Marion Leboyer, Christine Lochner, James T. McCracken, Sarah E. Medland, Preben B. Mortensen, Benjamin Neale, Humberto Nicolini, Merete Nordentoft, Michele Pato, Carlos Pato, David L. Pauls, Nancy L. Pedersen, John Piacentini, Christopher Pittenger, Danielle Posthuma, Josep A Ramos-Quiroga, Steven A. Rasmussen, Kerry J. Ressler, Margaret A. Richter, Maria C. Rosário, David R. Rosenberg, Stephan Ruhrmann, Jack F. Samuels, Sven Sandin, Paul Sandor, Gianfranco Spalletta, Dan J. Stein, S. Evelyn Stewart, Eric A. Storch, Barbara E. Stranger, Maurizio Turiel, Thomas Werge, Ole A. Andreassen, Anders D. Børglum, Susanne Walitza, Bjarne K. A. Hansen, Christian P. Rück, Nicholas G. Martin, Lili Milani, Ole Mors, Ted Reichborn-Kjennerud, Marta Ribasés, Gerd Kvale, David Mataix-Cols, Katharina Domschke, Edna Grünblatt, Michael Wagner, John-Anker Zwart, Gerome Breen, Gerald Nestadt, Andres Metspalu, Jaakko Kaprio, Paul D. Arnold, Dorothy E. Grice, James A. Knowles, Helga Ask, Karin J. H. Verweij, Lea K. Davis, Dirk J. A. Smit, James J. Crowley, Carol A. Mathews, Eske M. Derks, Jeremiah M. Scharf, Manuel Mattheisen

## Abstract

Obsessive-compulsive disorder (OCD) is a heritable disorder, but no definitive, replicated OCD susceptibility loci have yet been identified by any genome-wide association study (GWAS). Here, we report results from a GWAS in the largest OCD case-control sample (N = 14,140 OCD cases and N = 562,117 controls) to date. We explored the genetic architecture of OCD, including its genetic relationships to other psychiatric and non-psychiatric phenotypes. In the GWAS analysis, we identified one SNP associated with OCD at a genome-wide significant level. Subsequent gene-based analyses identified additional two genes as potentially implicated in OCD pathogenesis. All SNPs combined explained 16% of the heritability of OCD. We show substantial positive genetic correlations between OCD and a range of psychiatric disorders, including anxiety disorders, anorexia nervosa, and major depression. We thus for the first time provide evidence of a genome-wide locus implicated in OCD and strengthen previous literature suggesting a polygenic nature of this disorder.

## Introduction

Obsessive-compulsive disorder (OCD) is a chronic mental illness that affects approximately 2-3% of the general population^1;2^. It is characterized by obsessions and compulsions that vary in type and severity across patients, with most showing a waxing and waning illness course. Age at onset is usually in early to late adolescence but diagnosis and initiation of treatment are often delayed by several years^3;4^. OCD frequently co-occurs with other disorders, particularly tic disorders, eating disorders, depressive disorders, and anxiety disorders^5–8^. Although anxiety is a common symptom of OCD, OCD is currently considered to be distinct from anxiety disorders in terms of its epidemiological, clinical and pathophysiological presentation. This uniqueness is recognized in DSM-5 and ICD11, which separate OCD, along with several related disorders, into a separate disorder category, Obsessive Compulsive and Related Disorders (OCRDs)^2^.

Although its pathogenesis is yet to be fully elucidated, it is now clear that complex genetic factors play a role in the susceptibility to and/or development of OCD. The heritability of obsessive-compulsive (OC) symptoms as estimated by twin studies is between 27 and 47% in adults and between 45 and 65% in children^9–11^. SNP-based heritability of OCD, estimated using genome complex trait analysis (GCTA) is between 28-37%, with higher estimates for childhood-onset OCD, in line with heritability estimates from twin studies^12;13^. GCTA analyses further indicate that much of the genetic risk for OCD arises from common variation in multiple loci, each with a small effect, acting in concert^12^. Although rare variants exerting larger effects may also play a role in OCD development^14–17^, less work has been done in this area.

Two genome-wide association studies (GWAS) of OCD as well as a combined meta-analysis of 2,688 cases and 7,037 controls have been published to date^13;18;19^. However, all three studies were significantly under-powered based on estimates that suggest a need for tens to hundreds of thousands of individuals (cases and controls) for definitive gene identification in psychiatric disorders with complex inheritance^20^. Accordingly, no definitive, replicated OCD susceptibility loci have yet been identified by any study. Nevertheless, while small, these prior GWASs did demonstrate significant SNP-based heritability, indicating that a well-powered OCD GWAS with a much larger sample size would likely identify genome-wide significant loci and more accurate SNP effects, similar to what has been demonstrated for other major psychiatric disorders^21–23^. Such findings will provide insight into the biological underpinnings of OCD and may improve follow-up analyses and contribute to (multimodal) models for risk prediction. In addition, the frequent co-occurrence seen between OCD and other psychiatric illnesses appears to be due, at least in part, to shared genetic susceptibility. Crossdisorder studies demonstrate that OCD is genetically correlated with a range of other psychiatric disorders, most notably Tourette syndrome (TS), anorexia nervosa (AN), and major depressive disorder (MDD)^24;25^, all of which are highly comorbid with OCD.

Here, we performed a GWAS in a substantially expanded case-control sample (N = 14,140 OCD cases and N = 562,117 controls). We explored the genetic architecture of OCD, including its genetic relationships to other psychiatric and non-psychiatric phenotypes, such as TS, AN, and MDD. In the GWAS analysis, we identified one SNP associated with OCD at a genome-wide significant level. Subsequent gene-based analyses nominated two additional genes as potentially implicated in OCD pathogenesis. All SNPs combined explained 16% of the heritability of OCD. We also show substantial positive genetic correlations between OCD and a range of psychiatric disorders, including those with anxiety disorders and MDD.

## Methods

### Samples

We analyzed genomic data from 17 OCD case-control cohorts (11,312 OCD cases and 557,230 controls in total) not included in any previous OCD GWAS publications. In addition, three previously published GWAS datasets (two from the International OCD Foundation-Genetics Consortium (IOCDF-GC)^18^ and one from The OCD Collaborative Geneticws Association Study (OCGAS)^19^) were re-analyzed using newly matched control samples that were genotyped with the same microarrays as the cases (2,828 cases and 4,887 controls). In total, 20 cohorts comprising 14,140 OCD cases and 562,117 controls of European ancestry were included in the analyses. Among these, 323 cases were part of a parent-proband trio; in these cases, parents were used as pseudo controls. A total of 12,607 cases met DSM-IV^26^ or ICD10 criteria for OCD, while the remaining 1,533 cases were based on self-reported OCD diagnosis. Supplementary Table S1 provides an overview of the individual cohorts, cohort-specific sample and analytic details can be found in the Supplementary data. Data collections were approved by the relevant institutional review boards at all participating sites, and all participants provided written informed consent.

### Genetic data formatting, cleaning, alignment, and individual GWASs

First, the data of each participating cohort were analyzed individually (see Supplementary Methods for details). Genetic data were imputed using either the Haplotype Reference Consortium (HRC)^27^ or 1000 Genomes Project Phase 3 reference panels^28^. The resulting GWAS summary statistics were then harmonized before a conjoint meta-analysis was conducted. Each summary statistic data set was transformed to ‘daner’ file format following ricopili^29^ specifications. Next, each dataset was cleaned of variants that were likely to have poor underlying genotype data. All variants had to meet the following criteria for inclusion: minor allele frequency (MAF) *>* 1% in cases and controls, imputation-quality (INFO) score *>* 0.8 and *<* 1.2. If the effect measure, p-value or standard error (SE) was missing or was out of bounds (infinite), the SNP was removed. Only biallelic SNPs were retained in the data. Once cleaned summary statistics were produced, all datasets were aligned to the HRC reference panel^27^. If variants were reported on different strands, they were flipped to the orientation in the HRC-reference. Furthermore, strand-ambiguous A/T and C/G SNPs were removed if their MAF was *>* 0.4. In case A/T and C/G SNPs showed a MAF *<* 0.4 allele frequencies were compared to frequencies in the HRC-reference. If an allele frequency match was found, i.e., the respective allele was also the minor allele in the HRC reference, same strand orientation was assumed. If an allele mismatch was found, i.e. the allele had a frequency > 0.5 in HRC, it was assumed that alleles were reported on different strands and alleles were flipped subsequently. Marker-names were uniformly switched to those present in the HRC reference. If a variant did not overlap with the variants in the HRC reference, it was removed.

### Genome-wide association meta-analysis

Inverse variance weighted meta-analysis was conducted on 20 European cohorts using METAL^30^. Heterogeneity was assessed with Cochran’s *I*^2^ statistic. The genomic control factor (*λ*_1000_) was calculated for each individual GWAS and for the overall meta-analysis to identify residual population stratification or systematic technical artifact. GWAS summary statistics were subjected to linkage disequilibrium (LD) score regression (LDSC) analyses on high-quality common SNPs (INFO score *>* 0.9) to examine the LDSC intercept as an alternative evaluation of test statistic inflation due to residual artifact or population stratification, and to estimate the genetic heritability from the meta-analysis and genetic correlations between cohorts. The genome-wide significance threshold for the GWAS was set at a p-value of 5.0×10^−8^. Sign tests on the top SNPs (inclusion threshold of *p* = 0.0001, *p* = 0.00001, *p* = 0.000001, and *p* = 5.00×10^−8^) were performed between each individual cohort as well as ‘leave-one-out’ meta-analyses to identify any cohort in which the summary statistics significantly deviated from the rest of the cohorts.

### Polygenic risk scoring

OCD polygenic risk scores (PRS) were calculated to assess the predictive value of the new meta-analysis. For this purpose, we conducted three ‘leave-one-out’ GWAS metaanalyses that each excluded one of three samples (IOCDF-GC, OCGAS, and Psych_Broad) and tested how well the PRS predicted case-control status in the excluded sample. We selected these three datasets based on their previous involvement in a recent meta-analysis (IOCDF-GC and OCGAS; see above) and their availability at the site that conducted the analysis (Psych_Broad). PRS were generated at eight p-value thresholds (0.001, 0.05, 0.1, 0.2, 0.3, 0.4, 0.5, and 1) as a weighted sum of the risk allele dosages. The contribution of PRS was measured by comparing Nagelkerke’s preudo-*R*^2^ of the full model (including principal components (PCs), sex, and PRS) and of the reduced model (adjusted for PCs and sex).

### Gene-based analyses

#### Conventional MAGMA (C-MAGMA)

We used conventional MAGMA (C-MAGMA v1.07)^31^ to test genetic associations at the gene level for the combined effect of SNPs in or near protein-coding genes while adjusting for LD between the SNPs and the size of the gene. We used the 1000 Genomes reference panel (Phase 3)^28^ to control for LD. SNPs were mapped to genes if they were located within 10 kb of the gene coding region. The significance threshold was set using Bonferroni correction (i.e. 0.05/20,031 tests performed: *p <* 2.50×10^−6^).

#### Expression informed analysis (E-MAGMA, S-PrediX-can, S-MultiXcan)

We conducted gene-based tests informed by expression quantitative trait loci (eQTL) to identify genes that are enriched for association signal. We used a novel eQTL-informed gene-based test (E-MAGMA)^32^ that assigns SNPs to genes based on significant associations (FDR-corrected *p <* 0.05) with tissue-specific eQTLs in 48 GTEx tissues (v8). E-MAGMA is conducted in MAGMA (v1.07) by modifying gene annotation files by integration of eQTL information and uses the 1000 Genomes reference panel (Phase 3) to model LD. The significance threshold was set using Bonferroni correction across all tissues at *p <* 2.51×10^−7^ (i.e. 0.05/199,421 tests performed).

We used S-PrediXcan^33^ to integrate expression quantitative trait loci (eQTL) information with our GWAS summary statistics and identify genes whose genetically predicted expression levels were associated with OCD. S-PrediXcan estimates gene expression weights by training a linear prediction model in samples with both gene expression and SNP genotype data. The weights are then used to predict gene expression from GWAS summary statistics, while incorporating the variance and covariance of SNPs from an LD reference panel. We used expression weights for 48 tissues from the GTEx Project (v8)^34^ and LD information from the 1000 Genomes Project (Phase 3)^28^ (see web resources). These data were integrated with *β* values and standard errors from the OCD GWAS meta-analysis to test the association between imputed levels of gene expression and OCD risk. The significance threshold was set at *p <* 1.67×10^−7^ using Bonferroni correction (i.e. 0.05/299,996). Next, we used S-MultiXcan, which integrates the Predixcan associations across the 48 GTEx tissues into a single test, thus maximizing statistical power. We applied a transcriptome-wide significance threshold of *p <* 2.31×10^−6^ (i.e. 0.05/21,601 tests).

#### Chromatin informed (H-MAGMA)

We further conducted gene-based tests that integrated genetic results with Hi-C information using H-MAGMA^35^. SNPs were assigned to genes by leveraging regulatory (chromatin interaction) relationships in human brain tissue. H-MAGMA is conducted in MAGMA (v1.07) using the 1000 Genomes reference panel (Phase 3) to model LD. The significance threshold was set using Bonferroni correction (i.e. 0.05/24,358: *p <* 2.05×10^−6^).

### Functional annotation of GWAS findings

We also performed an extensive functional annotation for all SNPs in genomic areas identified by our lead SNPs using FUMA (see web-resources)^36^ to identify the most likely causal variants. In addition, the list of genes significantly associated with OCD, as indicated by at least one of the four gene-based methods (Table 2; except *LOC101928274* as it is neither contained in the gene catalog of FUMA nor in the expression data) was used to create expression heatmaps to visualize the average expression value of the associated genes. Further, a differential gene expression analysis (DEG) was performed to test the expression of associated genes against all other genes for each expression data set (tissue and developmental stage) using GTEx v8 and BrainSpan data^37^.

### Tissue and cell-type enrichment analysis

To determine if there are specific tissues or cell-types whose gene expression profiles are enriched for OCD risk variation, we used an analysis protocol recently described in Bryois et al.^38^. Consistent with this we utilized their code-base for this analysis, and in particular the sets of genes that mark different highlighted tissue and cell-type datasets (see web-resources for data source).

We selected 3 datasets that had been preprocessed by Bryois et al.^38^ for inclusion in this analysis. The first features tissue-specific gene expression data derived from GTEx^39^, with a total of 37 tissues represented. The second and third datasets are derived from Zeisel et al.^40^, and represent 1) *broad cell-type groups* across the entirety of the mouse nervous system, and 2) a *high-resolution single cell-type map* of the same data. In Zeisel et al.^40^, a total of 39 broad cell-type groups and 265 individual cell-types are represented.

We followed the analysis protocol described in Bryois et al.^38^ for the analyses of 37 tissues from GTEx and 39 broad cell-type groups from Zeisel et al.^40^, and utilized a simplified approach for the analysis of each of the 265 individual cell-types from Zeisel et al.^40^. For the tissue and broad cell-type group datasets, we conducted the full protocol from Bryois et al^38^ which included analyzing tissue/cell-type enrichment using both LDSC^41^ and MAGMA^31^. For each dataset, p-values were adjusted for a false discovery rate of 5%. We only considered a tissue or cell-type significantly enriched if the FDR-adjusted p-value was less than 0.05 in both the LDSC and MAGMA-based tests. Due to the high computational demands of analysing 265 individual cell-types across the mouse nervous system in LDSC, we limited the assessment protocol to using MAGMA only, and considered a cell-type as significant if it had an FDR-adjusted p-value of less than 0.05. All statistical analyses downstream of LDSC and MAGMA (namely, p-value adjustment) were done using R v3.6.1, and all plotting was done inside of R v3.6.1 using the package ggplot2 v3.2.1^42^.

### Overlap of the genome-wide significant locus with high-confidence chromatin interactions

To derive chromatin interactions found in brain tissue that overlap with the genome-wide significant region(s) from the OCD GWAS, we submitted a corresponding analysis job to FUMA^36^ on May 3rd 2020 using the OCD GWAS sumstats as input. We selected the following chromatin interaction datasets for inclusion: 1) Promoter anchored loops from the PsychENCODE project^43^, 2) loops from fetal cortex tissue, from Giusti-Rodriguez et al. 2019^44^, and 3) loops from adult cortex tissue, also from Giusti-Rodriguez et al. 2019^44^. All other parameters were set to default settings (SNP2GENE: a) maximum p-value of lead SNPs *<* 5*x*10^−8^, maximum p-value cutoff *<* .05, *r*^2^ threshold to define independent significant SNPs. *≥* 6, second *r*^2^ threshold to define lead SNPs. *≥* 1, reference population: 1000G phase 3 EUR, maximum distance between LD blocks to merge into a locus *<* 250kb. b) Positional gene mapping: distance to genes or functional consequences of SNPs on genes to map: 50 kb, SNP filtering: minimum CADD score *≥* 12.37, maximum RegulomeDB score = 7, annotation datasets eQTL catalogue: PsychENCODE, FANTOM5, Brain Open Chromatin Atlas. c) Gene eQTL mapping: Tissue types from Brain-Seq Brain, BRAINEAC, GTEx v8 Brain, eQTL; eQTL p-value threshold: only significant snp-gene pairs with FDR *<* .05. GENE2FUNC: all background genes used, gene expression datasets: GTEx v8:54 tissue types, 30 general tissue types, 29 different ages of brain samples, and 11 general developmental stages of brain samples, Benjamin-Hochberg (FDR) correction was used to test for multiple testing in gene-enrichment analyses, maximum adjusted p-value for gene set association *<* .05, minimum overlapping genes with gene sets *≥* 2).

### SNP-based heritability and genetic correlation with other traits

The proportion of variance in liability to OCD that could be explained by the aggregated effect of all included SNPs (SNP-based heritability) was estimated using LDSC^41^. The SNP heritability is based on the estimated slope from the regression of the SNP effect sizes from the GWAS on the LD score. Genetic correlations between OCD and 82 other disorders/traits of potential relevance to OCD were estimated in cross-trait LDSC, a method that computes genetic correlations between GWASs without bias from ancestry differences or sample overlap^45^. The genetic correlation between traits is based on the estimated slope from the regression of the product of Z-scores from two GWASs on the LD score and represents the genetic covariation between two traits based on all polygenic effects captured by the included SNPs. The source studies of the GWAS summary statistics used are summarized in Supplementary Table S9. The genome-wide LD information used by these methods was based on European populations from the HapMap 3 reference panel^45;46^ (see web resources). The GWAS summary statistics were filtered to only include SNPs that were part of the 1,290,028 million genome-wide HapMap 3 SNPs used in the original LD score regression studies^45;46^. As a follow-up, we repeated the same analyses separately for three OCD subsets: 1) including only clinical samples, 2) including only (large-scale) biobank samples, and 3) including only data sets that were not primarily ascertained for OCD (AUS and iPSYCH).

## Results

### Genome-wide association results

Prior to conducting the GWAS meta-analysis, QQ plots and genomic inflation factors (reported in Supplementary Table S1 as λ1000) from each of the individual GWAS data sets were evaluated, and no evidence for significant residual population stratification was observed. The final GWAS meta-analysis contained 20 data sets consisting of 14,140 OCD cases, 562,117 controls and 7,027,156 autosomal SNPs. No significant residual stratification effects were observed (Supplementary Figure S1 QQ plot; λ = 1.151; λ_1000_ = 1.005). One SNP (rs2581789, *p* = 2.1×10^−8^, OR = 0.92) exceeded the genome-wide threshold for significance (See Figure 1 for Manhattan plot, Figure 2 for regional plot of rs2581789, and Figure 3 for a forest plot showing the effect sizes in each individual cohort for rs2581789). This SNP is located in the intergenic region between *SFMBT1* and *RFT1* on chromosome 3p21.1. The LD block tagged by rs2581789 spans 325.8 kb (LD r^2^ > 0.6) and encompasses 12 genes, including *NEK4, ITIH1, ITIH3, ITIH4, ITIH4-AS1, MUSTN1, STIMATE-MUSTN1, STIMATE, MIR8064, SFMBT1, RFT1*, and *PRKCD*. Eight additional independent GWAS loci with p-values < 1.0×10^−5^ were identified (corresponding association data, genomic regions, and genes in LD with the lead SNP are reported in Table 1, see Supplementary Figure S2-S9 for regional plots and Supplementary Figures S10-S17 for forest plots of each SNP).

**Fig. 1.**
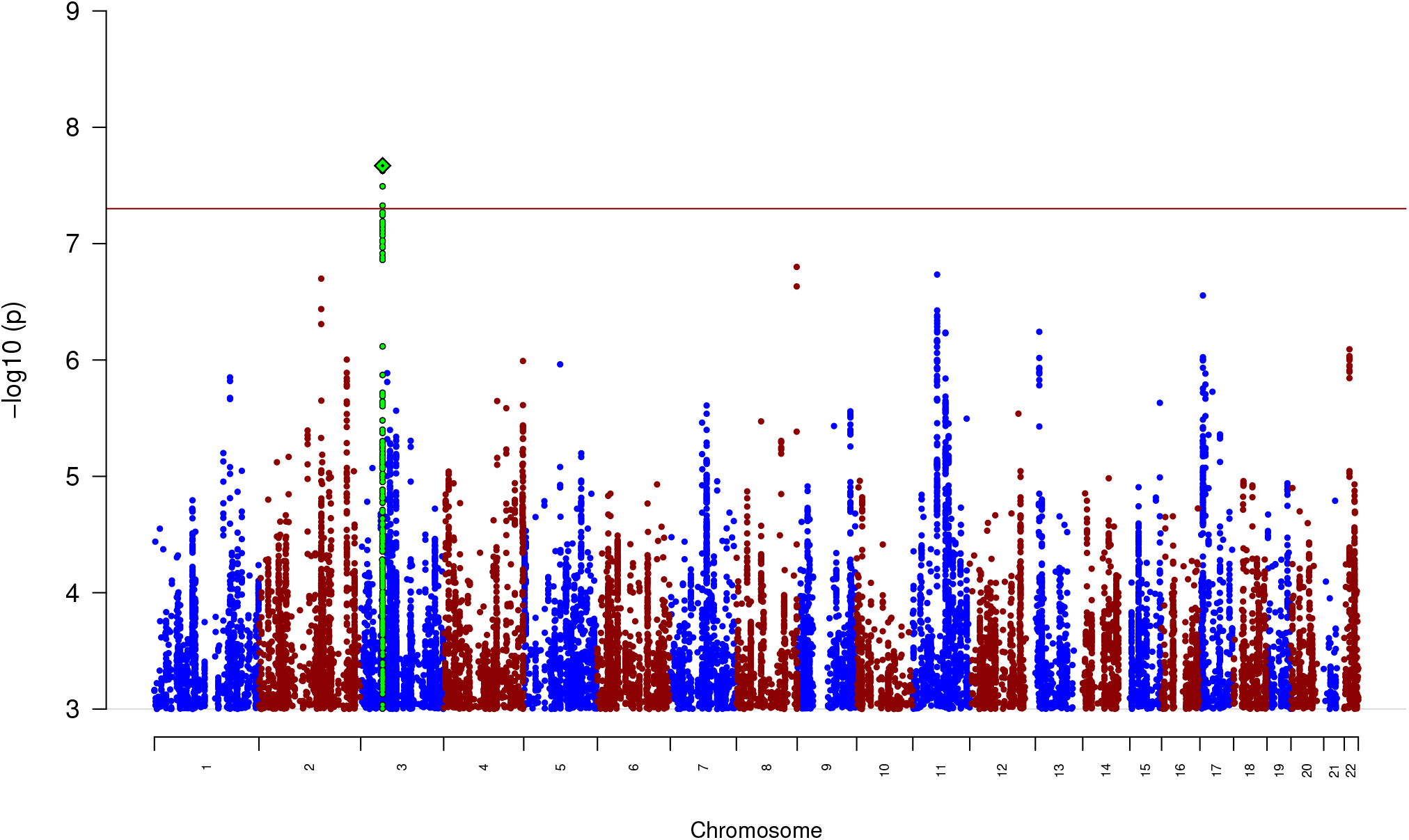
Manhattan plot of the results from the GWAS meta-analysis of OCD: The y-axis represents –log10 p values for association of variants with OCD, from metaanalysis using an inverse-variance weighted fixed effects model. The x-axis represents chromosomes 1 to 22. The horizontal red line represents the threshold for genome-wide significance. The index variant of the one genome-wide significant locus is highlighted as a green diamond.

**Fig. 2.**
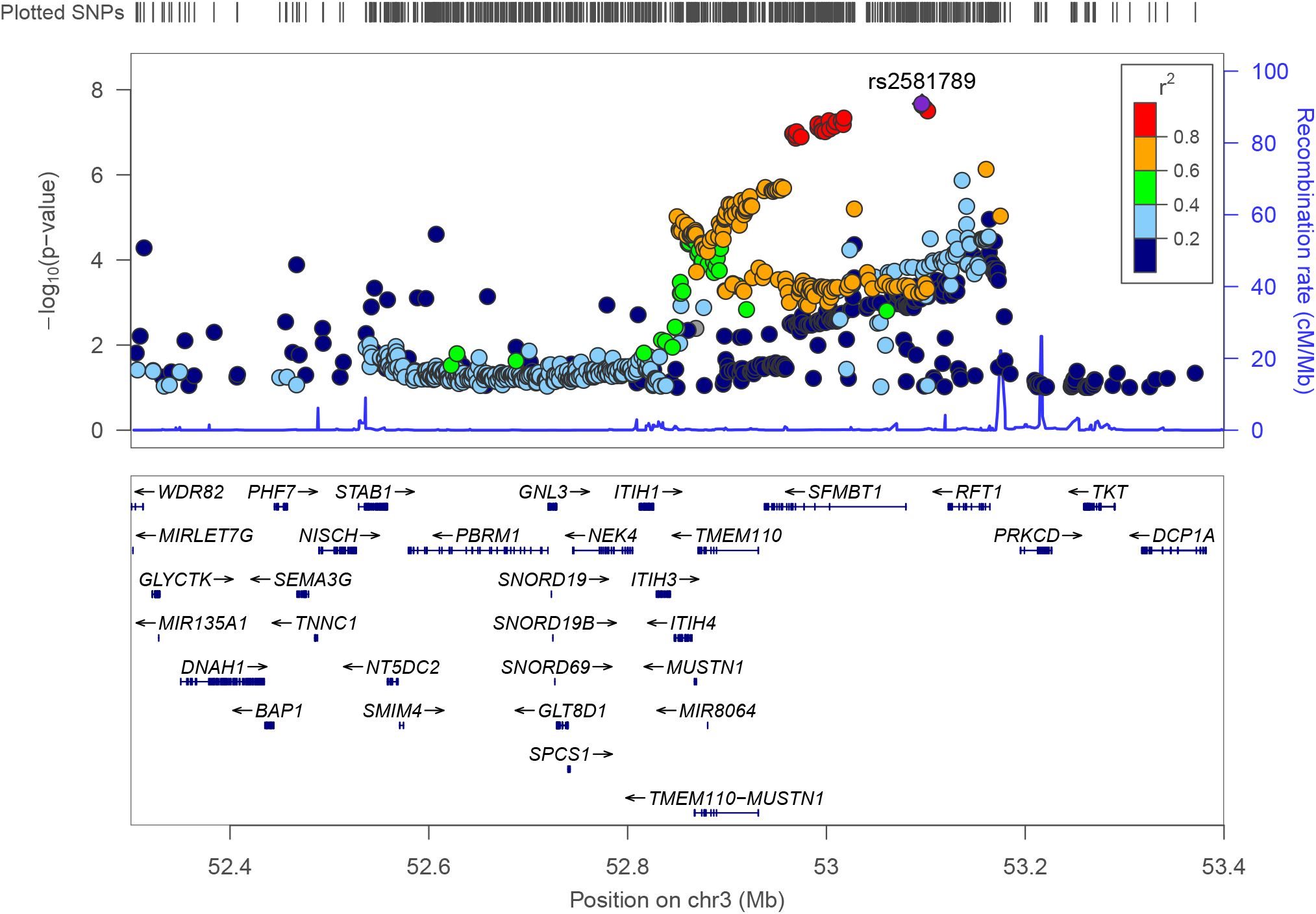
Regional plot of the genome-wide significant locus rs2581789. The –log_10_ (p-value) of SNPs in the OCD meta-analysis GWAS is shown on the left y axis. The recombination rates expressed in centimorgans (cM) per Mb (Megabase) (blue line) are shown on the right y axis. Position in Mb is on the x axis. Only the SNPs with association p-value less than 0.1 were plotted. The most associated SNP is shown as a purple diamond.

**Fig. 3.**
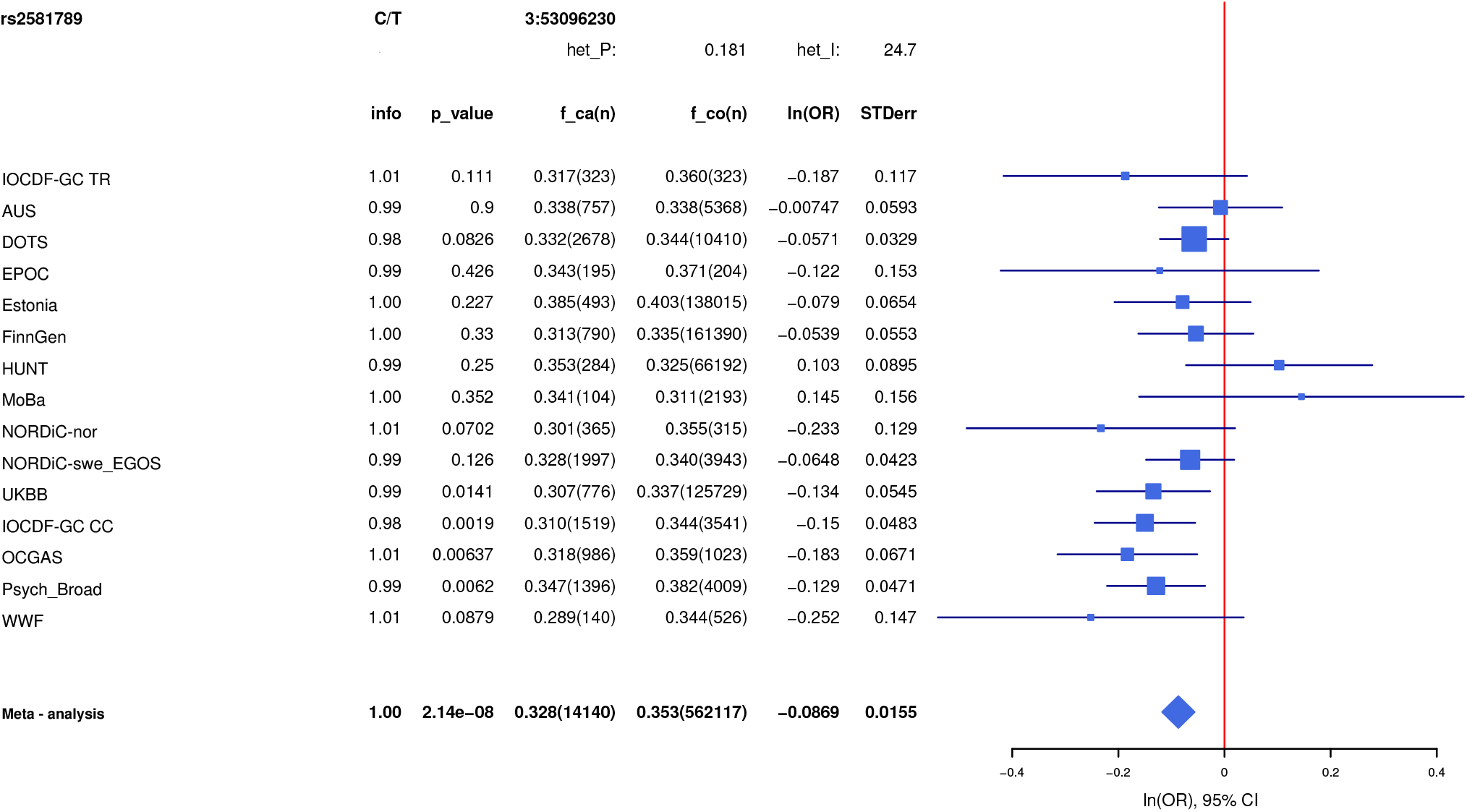
Forest plot of SNP rs13262595. The plot shows the effect estimate with 95%-confidence interval for each cohort contributing to the meta-analysis and for the inverse variance weighted meta analysis. The table lists INFO (imputation score), p-value, f_ca(n) (frequency cases), f_co(n) (frequency controls), ln(OR), and STDerr (standard error) for each of the contributing cohorts and for the meta-analysis. At the top, + indicates a positive direction of effect, - a negative direction of effect while ? indicates that the SNP was not contained in the respective cohort.

**Table 1.** LD-independent genomic regions with *p* < 1×10^−6^ in the OCD meta-analysis and their associated genes

| SNP | CHR | BP | P | OR | SE | A1/A2 | FRQ_A | FRQ_U | INFO | genes.6.50kb(dist2index) |
| --- | --- | --- | --- | --- | --- | --- | --- | --- | --- | --- |
| rs2581789 | 3 | 53096230 | **2.138x10<sup>-8</sup>** | 0.91677 | 0.0155 | C/T | 0.328 | 0.353 | 0.996 | NEK4(-241.3), ITIH1(-220.2), ITIH3(-203.2), ITIH4(-181.5), ITIH4-AS1(-186.9), MUSTN1(-177.2), STIMATE-MUSTN1(-114.7), STIMATE(-114.7), MIR8064(-165.7), SFMBT1(0.0), RFT1(0.0), PRKCD(49.0) |
| rs13262595 | 8 | 143316970 | 1.585x10<sup>-7</sup> | 1.07573 | 0.0139 | A/G | 0.454 | 0.424 | 0.993 | LINC00051(0.0), TSNARE1(0.0) |
| rs674094 | 11 | 57665336 | 1.844x10<sup>-7</sup> | 1.08622 | 0.0159 | A/C | 0.319 | 0.332 | 0.99 | SERPING1(-233.0), MIR130A(-206.6), YPEL4(-197.9), CLP1(-186.0), ZDHHC5(-146.7), MED19(-135.5), TMX2(-106.9), TMX2-CTNND1(-28.7), SELENOH(-104.3), BTBD18(-96.1), CTNND1(-28.3) |
| rs79712033 | 2 | 147846501 | 1.999x10<sup>-7</sup> | 1.17975 | 0.0318 | A/G | 0.945 | 0.937 | 0.971 | - |
| rs7219489 | 17 | 8121622 | 2.79x10<sup>-7</sup> | 1.08405 | 0.0157 | G/T | 0.732 | 0.727 | 0.994 | ALOXE3(-49.4), HES7(-44.2), PER1(-15.9), MIR6883(-23.2), VAMP2(-5.4), TMEM107(0.0), SNORD118(0.0), MIR4521(0.0), BORCS6(0.0), AURKB(0.0), LINC00324(0.0), CTC1(0.0), PFAS(0.0), SLC25A35(19.5), RANGRF(20.4), ARHGEF15(41.9), ODF4(71.5) |
| rs9535127 | 13 | 31606961 | 5.725x10<sup>-7</sup> | 1.07659 | 0.0148 | T/C | 0.487 | 0.49 | 0.993 | TEX26(-7.3) |
| rs7128224 | 11 | 77360928 | 5.817x10<sup>-7</sup> | 1.0835 | 0.016 | T/G | 0.316 | 0.308 | 0.989 | LOC646029(-24.2), AQP11(0.0), CLNS1A(0.0), RSF1(0.0), AAMDC(121.2), INTS4(178.8), KCTD14(315.8), NDUFC2-KCTD14(315.8) |
| rs424541 | 22 | 29863922 | 8.089x10<sup>-7</sup> | 0.91439 | 0.0181 | C/T | 0.818 | 0.789 | 0.997 | AP1B1(-29.4), RFPL1S(0.0), RFPL1(0.0), NEFH(0.0), THOC5(0.0), NIPSNAP1(36.9), NF2(85.6) |
| rs34289388 | 2 | 208280861 | 9.928x10<sup>-7</sup> | 1.08741 | 0.0171 | G/A | 0.8 | 0.788 | 0.998 | - |

Notes: SNP: single-nucleotide polymorphism; CHR: chromosome; BP: base pair position (hg19); P: p value; OR: odds ratio for allele1; A1/A2: effect allele/ reference allele; FRQ\_A: frequency of allele1 in affected cases; FRQ\_U: frequency of allele1 unaffected controls; INFO: imputation INFO score; genes.6.50kb(dist2index): list of genes within the region of LD-friends.6 (variants with LD-r<sup>2</sup> > 0.6 to index SNP) (±50 kb), in brackets distance to index SNP in kb.

This SNP (rs2581789) has previously been reported to be associated with schizophrenia (SCZ)^47^ and a combined SCZ/bipolar disorder (BP) phenotype^48^, as well as with anthropomorphic traits such as body mass index (BMI)^49^ and waist-hip-ratio^49^, and with psychological traits such as worry^50–52^ and well-being^53^. Other top-ranked SNPs identified in this analysis have also been previously reported to be significantly associated with psychological traits such as anxiety^54^, depressive symptoms^53^, neuroticism^50;52;53;55^, hurt^50^, worry too long after embarrassment^50;52^, nervous feelings^50^ and worry/vulnerability^54^, as well as with cognitive performance and educational attainment^56^ (see Supplementary Table S2 for a full list of previously reported significant associations of the top nine SNPs identified in the OCD GWAS).

### Polygenic risk scoring

In the ‘leave-one-out’ PRS analyses we observed significantly higher OCD PRS among OCD cases compared to the controls in each target sample, explaining 3.9%, 3.5%, and 3.3% of overall phenotypic variance of IOCDF-GC, OCGAS, and Psych_Broad, respectively (Supplementary Figure S18).

### Gene-based analyses

We identified 18 genes that were significantly associated with OCD in at least one of the four gene-based methods (C-MAGMA, E-MAGMA, H-MAGMA, and S-PrediXcan) following correction for multiple hypothesis testing at the experiment-wide or transcriptome-wide level (see Table 2 for a list of all significant genes; full results can be found in Supplementary Tables S3, S4, S5, and S6). Genes identified by the four methods partially overlapped (Venn-diagram, Supplementary Figure S19). C-MAGMA revealed five genes significantly associated with OCD: *PER1, SFMBT1, CTC1, TMEM107*, and *NEFH* at a Bonferroni-corrected significance threshold of *p* < 2.50×10^−6^ (Supplementary Table S3). H-MAGMA identified 14 risk genes across 7 genomic regions associated with OCD (Bonferroni-corrected *p* < 2.05×10^−6^; Supplementary Table S4). Using E-MAGMA, we identified one gene that was significantly associated with OCD (*CTC1* in whole blood, *p* < 2.51×10^−7^, Supplementary Table S5). *CTC1* was also the most significantly associated gene using S-PrediXcan (whole blood, *p* = 3.04×10^−7^), but did not survive multiple testing correction in this analysis (*p* < 1.78×10^−7^) (Supplementary Table S6). We therefore meta-analyzed the tissue-specific S-PrediXcan associations (i.e. combining association signals across 48 GTEx tissues) using S-MultiXcan to generate a single test statistic (see Supplementary Table S7) for each gene. This approach yielded two significant (*p <* 2.31*x*10^−6^) genes: *TMEM107* (*p* = 2.44×10^−7^) and *RPL35* (*p* = 1.01×10^−6^).

**Table 2.**
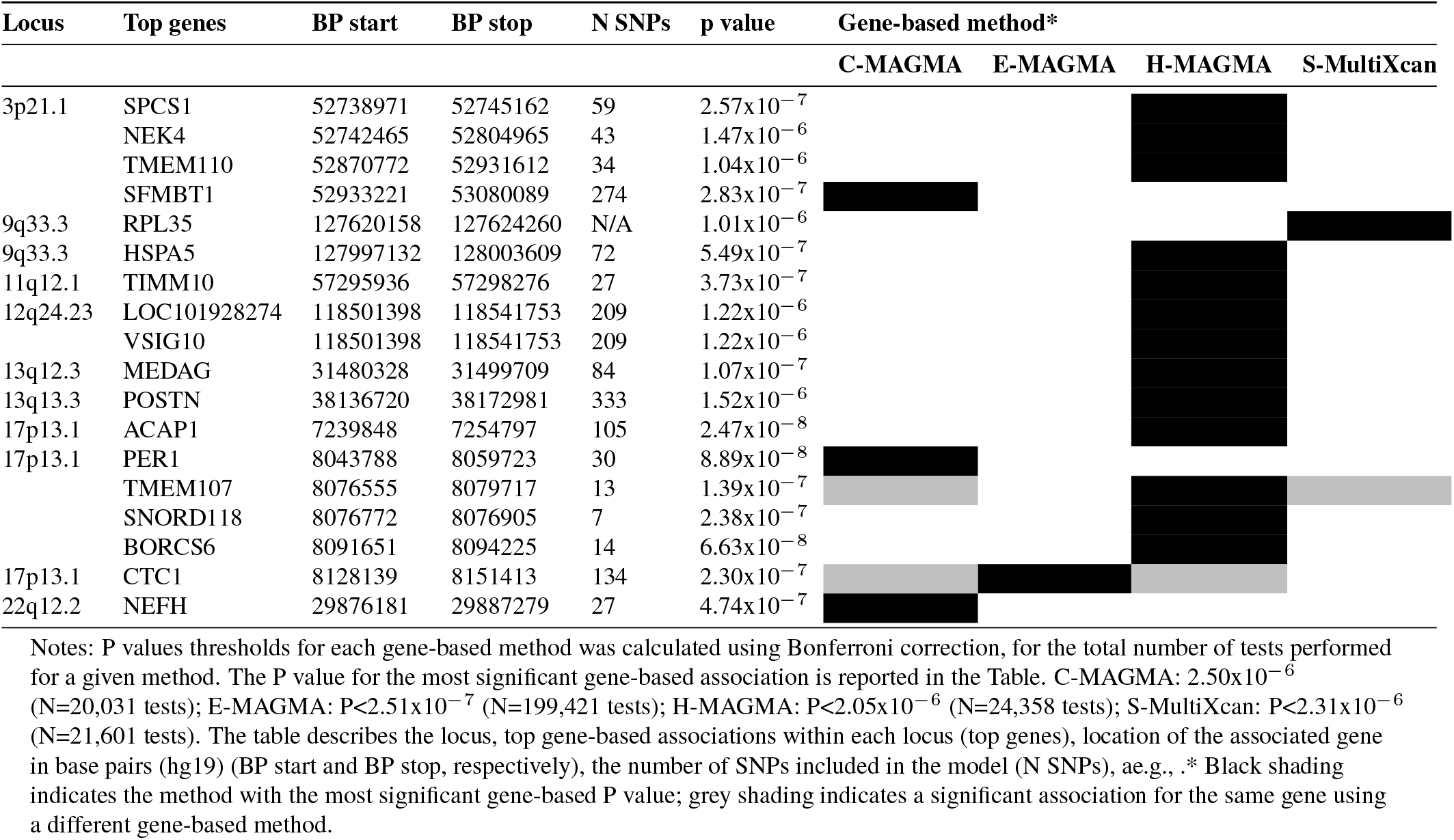
Genes significantly associated with OCD, as indicated by 4 gene-based methods

### Functional annotation of GWAS findings

The functional annotation of individual GWAS findings (FUMA) is presented in Supplementary Table S8) and in the regional annotation plot for the genome-wide significant chromosome 3 locus (see Supplementary Figure S20). We explored tissue-specific expression patterns for 17 of the 18 genes resulting from the gene-based tests (excluding *LOC101928274* for which no information was available). Results are visualized for the 54 GTEx tissues (see Supplementary Figure S21) and for Brainspan gene expression data (Supplementary Figures S22 and S23). Deferentially Expressed Gene (DEG) set analyses were conducted to test whether the set of 17 genes implicated by one or more genebased tests are significantly up-regulated or down-regulated in any of the tissues. No significant differences were found (Supplementary Figures S24, S25, and S26).

### Tissue and cell-type enrichment analyses

We performed association tests for enrichment between OCD GWAS summary statistics and three gene expression profiles from human tissues and mouse cell-types. No tissue or cell-type was significantly associated with OCD after FDR correction for both the LDSC and MAGMA methods (Supplementary Figure S27A), thus failing to reach our criterion for significance. Nevertheless, tissues derived from brain regions clustered preferentially at the top of the results distribution. Also, no broad cell-type category reached significance for both LDSC and MAGMA (Supplementary Figure S27B). Here, the top-ranking result was ‘telencephalon projecting excitatory neurons’. A further eight types included three types of di- and mesencephalon inhibitory neurons from the midbrain, two types of di- and mesencephalon excitatory neurons (one from thalamus and one from midbrain), and two types of telencephalon inhibitory interneurons from the hippocampus/cortex. In the high-resolution single cell-type analysis using MAGMA, nine specific cell-types were significantly enriched for OCD after FDR-correction (Supplementary Figure S28).

### Overlap of the genome-wide significant locus with high-confidence chromatin interactions

Genome-wide significant hits in other psychiatric GWAS studies have been shown to be enriched in overlap with brain-specific chromatin interactions^44;57^, and we sought to determine if this overlap (and potential subsequent mechanism of effect) existed for the genome-wide significant locus. We utilized FUMA^36^ to determine if the genome-wide significant locus from the GWAS (lead SNP located at hg19 chr3:53096230) overlapped with any high-confidence chromatin interactions derived from either PsychENCODE^57^ or fetal and adult cortex from Giusti-Rodriguez et al.^44^. While several loops connected the locus with a distal gene on the same chromosome (see Supplementary Figure S29), follow-up analyses did not reveal any genes with an obvious dosage sensitivity.

### SNP-based heritability and genetic correlations with other traits

The SNP-based heritability of OCD as obtained from LD-score regression was estimated to be 0.164 (SE = 0.012). In genetic correlation analyses between OCD and summary statistics from 82 behavioral, cognitive, psychiatric, neurological, allergic/immunologic, metabolic, and anthropomorphic traits 31 of the 82 traits investigated had significant genetic correlations after correction for multiple testing (Figure 4 and Supplementary Table S9). In particular, OCD showed significant positive genetic correlations with all psychiatric disorders, with especially high correlations with anxiety disorder (*r*_*g*_ = 0.627, *SE* = 0.057, FDR-corrected *p* = 6.11×10^−27^, AN (*r*_*g*_ = 0.588, *SE* = 0.04, FDR-corrected *p* = 1.91×10^−48^), and MDD (*r*_*g*_ = 0.54, *SE* = 0.042, FDR-corrected *p* = 6.58×10^−36^). OCD was also positively correlated with alcohol dependence, while cannabis use disorder and other substance-use traits did not show significant correlations. Moreover, OCD was significantly genetically correlated with several cognitive/SES related traits, including a positive correlation with memory and negative correlations with intelligence, income, and job satisfaction. Also, some of the anthropomorphic traits (BMI, waist-hip-circumference, and hip-circumference) and auto-immune disorders (Crohn’s disease, ulcerative colitis, and inflammatory bowel disease) showed a significant negative correlation with OCD. In addition, significant correlations of substantial effect (r > |0.25|) were found with neuroticism, suicidality, tiredness (all positive), and with subjective well-being (negative). Somewhat less prominent, but also significant was the positive correlation with childhood maltreatment. Genetic correlations with neurological, substance use, cardiovascular, and fertility traits were weak and non-significant.

**Fig. 4.**
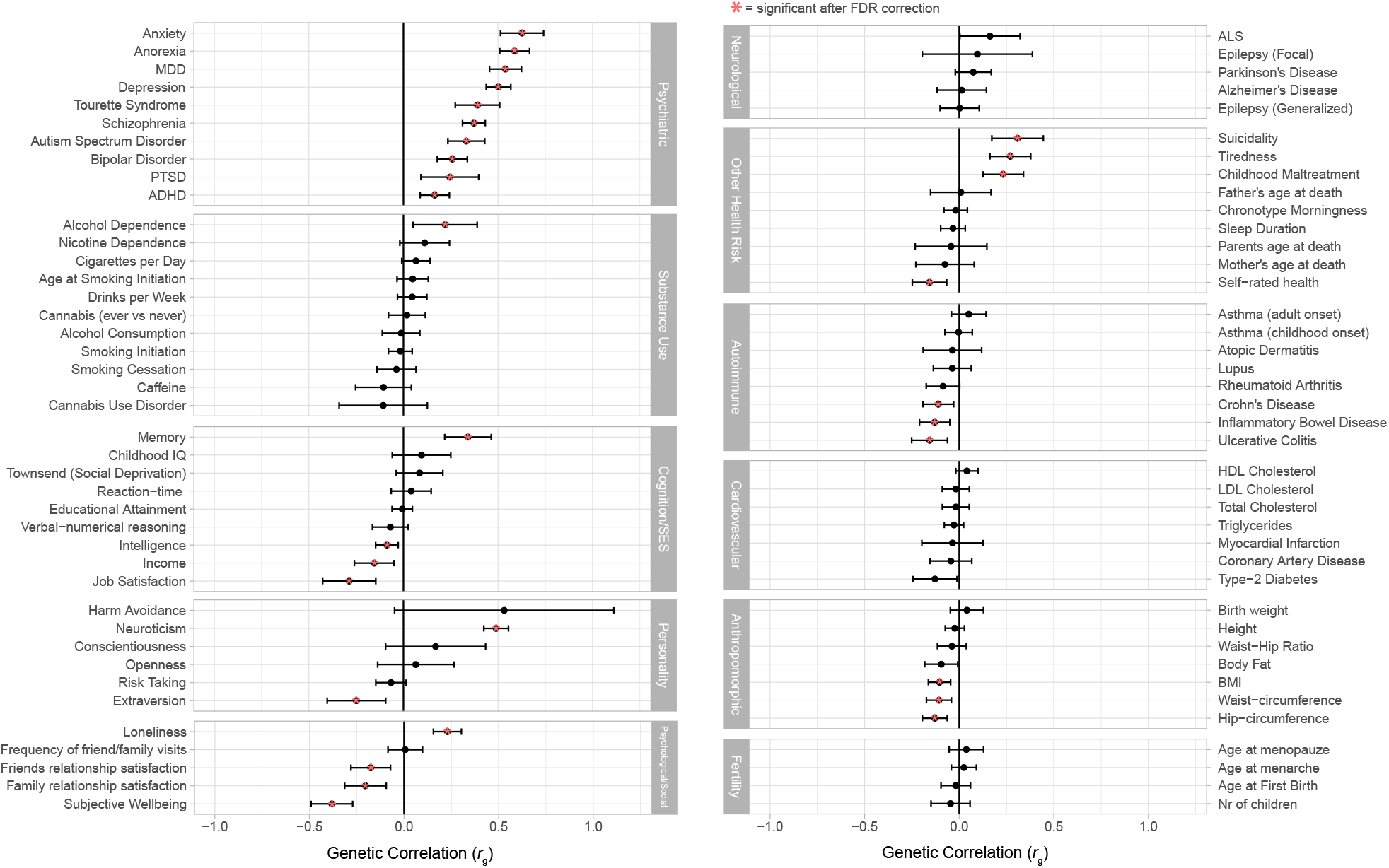
Genetic correlations (rg) between OCD and a broad range (N=82) of other phenotypes. Error bars represent 95% confidence intervals and asterisks indicate significant associations after FDR correction for multiple testing. Non-significant correlations with SE *>* 0.5 are excluded from display.

As follow-up sensitivity analyses, we repeated the genetic correlation analyses for clinically-ascertained samples and biobank samples separately, as well as for the full OCD GWAS sample excluding the two datasets that were not primarily ascertained for OCD (AUS and DOTS). The resulting correlations were similar, though not identical, across these three OCD sub-samples (see Supplementary Figure S30). As an example for differences in the sub-samples, only the clinically-ascertained OCD samples had significant genetic correlations with ADHD and PTSD. Generally, the biobank-derived OCD samples showed less pronounced correlations than the other two subsets, which might be expected due to their smaller case sample sizes.

## Discussion

In this GWAS, containing approximately five times the number of individuals diagnosed with OCD than previous studies^13;18;19^, we report the first definitive genome-wide significant locus for OCD (rs2581789 on chromosome 3p21.1). All SNPs combined in our analysis explained 16% of the heritability of OCD. We also showed substantial positive genetic correlations between OCD and a range of psychiatric disorders, including anxiety disorders, AN, and MDD.

Our top SNP rs2581789 resides in a gene-rich genomic region (see Figure 2 for a regional association plot and a list of genes in the region). This region has previously been associated with a broad range of other psychiatric disorders and related traits, including SCZ^47^, well-being^52^, and the worry-subcluster of neuroticism^50^. Of note, this SNP was also identified in a recent cross-disorder meta-analysis of eight psychiatric disorders from the Psychiatric Genomics Consortium (PGC)^24^ (*p* = 6.51×10^−14^ without the 23andMe data). Although the PGC cross-disorder analysis used a subset of the samples included in the current study, these previously published OCD cases^13^ represent only 20% of the cases in the current analysis, and did not have the power to identify any individual locus with genome-wide significance for OCD alone^13^. Despite being under-powered in the crossdisorder analysis^24^, OCD was among only three of the eight phenotypes that were shown to contribute to the rs2581789 association results with m-values > 0.9 (SCZ and BP being the other two). These findings indicate that the genome-wide significant SNP in this study is not solely associated with OCD, but rather with multiple psychiatric phenotypes, reflecting a pleiotropic effect, possibly contributing to a shared underlying neurobiological susceptibility across a subset of psychiatric disorders and other neurobehavioral traits. These findings add to the confidence that this SNP, and the encompassing 3p21.1 region, is robustly, but not uniquely associated with OCD. Notably, another recent manuscript^58^, reported one genome-wide significant region for a compulsive disorder factor (rs9821797 on chromosome 3p21.31, P = 3.61×10^−9^, r^2^ with rs2581789 < 0.01 and D < 0.2 in European ancestry individuals), using structural equation modeling and summary statistics from 11 major psychiatric disorders. This locus reached a p-value of 4.32×10^−4^ in the present OCD GWAS, although it should be noted that there was partial overlap between samples using the same sample as the PGC CDG study (see above).

Gene-based analyses, incorporating multiple types of functional annotations, including eQTLs, fetal and adult brain Hi-C information, and enrichment of tissue and single-cell RNAseq data, revealed no consistent associations. This is likely due to the absence of multiple, independent genomewide association loci from the primary GWAS, as these are typically necessary to have enough statistical power for such studies. However, two genes were identified by three of the four gene-based methods: the Transmembrane Protein 107 gene (*TMEM107*) and the CST Telomere Replication Complex Component 1 gene (*CTC1*), both located in the same locus on chromosome 17 (see Table 2 for the respective results of the four different gene-based methods). Given that SNPs in these genes were not identified as genome-wide significant in the primary GWAS meta-analysis, there is not yet sufficient evidence to consider the potential role(s) of genes or SNPs in these loci in the etiology of OCD.

Our analyses of the shared genetic risk between OCD and other psychiatric disorders provides deeper insights into the etiology of OCD. Previous work, including the cross-disorder analyses discussed above^24;58^, have confirmed the shared genetic risk between OCD and two neuropsychiatric disorders that are highly co-morbid with OCD - TS^12;59^ and AN^60;61^. The above mentioned cross-disorder analyses also identified other psychiatric disorders as being correlated with OCD, albeit less strongly (SCZ, BP, and MDD). The present study, with its substantially increased sample size, confirms the genetic relationships between OCD and all of these disorders (TS, AN, SZ, BP, and MDD), and for the first time also shows significant genetic correlations between OCD and anxiety disorders, ADHD, as well as ASD (see Figure 4, Supplementary Figure S30 and Supplementary Tables S9 and S10).

In addition, our analyses of genome-wide genetic correlations between OCD and a wide range of additional brain-and non-brain-based traits identified a number of significant results that warrant a more in-depth study in the future (see also below for potential limitations of our study). The significant negative association between genome-wide genetic risk for OCD and BMI, body-fat and hip circumference may be reflective of the shared genetic risk between OCD and AN, while the positive genetic correlation between OCD and tiredness, loneliness, neuroticism, and suicidality may represent the known clinical overlap of OCD, anxiety and MDD at a genetic level. Less clear regarding its interpretation is the significant genetic correlation between OCD and childhood maltreatment. Prior clinical studies have reported increased rates of childhood maltreatment in individuals with OCD^62;63^, which may correlate with co-occurring depression and anxiety. However, further studies are needed to shed light on the genetic relationship and the overlap in genetic contributions to their etiology.

One potential study limitation is the fact that, due to potentially higher than normal rates of psychiatric co-morbidity in many of our OCD datasets (e.g. through inclusion of individuals diagnosed with more severe OCD), our results may be biased towards identifying genes that contribute to multiple comorbid psychiatric disorders, or alternatively to an underlying trait related to general psychopathology, rather than to OCD specifically. A substantial proportion of new samples in the current study were derived from large-scale biobanks and national registers, two of which were drawn from studies that originally aimed to study other psychiatric disorders, such as depression (AUS), or that included a higher-than-average number of cases with depression, ADHD, and ASD (iPSYCH). As such, there is a larger degree of heterogeneity in our meta-analysis which likely influenced the genetic correlation results. We conducted sensitivity analyses to address and to mitigate these potential biases by dividing the OCD meta-analysis into three subsets including a) only samples from large-scale biobanks, b) only clinical samples, and c) excluding the two studies with comorbid ascertainment (AUS and iPSYCH) from the analysis. We observed in part substantial changes in the point estimates of the genetic correlations with some disorders and traits (e.g. depression, anxiety disorders, and ADHD). We are currently conducting additional analyses to further address these limitations and will update the current version of this manuscript as soon as these results become available. Our observations echo similar results in other disorders (e.g. MDD) that call for more sophisticated analytical approaches (e.g. through inclusion of sensitivity meta-analytic methods based on structural equation modelling and other techniques) in order to combine samples with heterogeneous ascertainment. As with these other disorders the limitations inherent in our study design and in the available samples should be addressed in future genetic studies of OCD. In light of the above highlighted results, it is of note that we also performed leave-one-out polygenic risk score analyses using, among others, the previously published OCD meta-analysis of clinically-ascertained cases^13^, which was used in all previous cross-disorder studies mentioned above. We found significant overlap in genetic liability between the prior OCD GWAS meta-analysis and leave-one-out GWAS data using predominantly samples that were newly added (mostly from large-scale biobanks) to our current meta-analysis (see Supplementary Figure S18). Once sample sizes sufficiently large to identify and replicate clear OCD susceptibility genes are collected, further analyses such as functional characterization, Mendelian randomization, and additional genetic correlation analyses may prove to be useful with the goal of further elucidating the underlying genetic etiology of OCD.

In sum, the present study provides insights into the current state of the largest GWAS for OCD to date. Our study identified the first definite genome-wide significant association of a locus with OCD (on chromosome 3p21.1). We are confident that with inclusion of additional samples (in the near future) we will be able to add new genome-wide significant regions to our current findings and further our understanding of OCD genetics. With these new meta-analysis results we will then also be able to use approaches (e.g., Mendelian Randomization and other) that will allow us to study the causal relationship between the genetic underpinnings of closely related disorders.

## Data Availability

Results from this study can be made available upon request to the authors as part of our standardized secondary analysis proposal program (please contact the corresponding author).

https://drive.google.com/file/d/1R465fjW4yH2cnx1EOLw6LqaZt-ZStIdc/view?usp=sharing

## ACKNOWLEDGEMENTS

A.A. and KJHV were supported by the Foundation Volksbond Rotterdam. MoBa is supported by the Norwegian Ministry of Health and Care Services and the Ministry of Education and Research. We are grateful to all the participating families in Norway who take part in this on-going cohort study. We thank the Norwegian Institute of Public Health for generating high-quality genomic data. This research is part of the HARVEST collaboration, supported by the Research Council of Norway (RCN) (229624). We also thank the NORMENT Centre for providing genotype data, funded by the RCN (223273), South East Norway Health Authority (SENHA) and KG Jebsen Stiftelsen. We further thank the Center for Diabetes Research, the University of Bergen for providing genotype data and performing quality control and imputation of the data funded by the ERC AdG project SELECTionPRE-DISPOSED, Stiftelsen Kristian Gerhard Jebsen, Trond Mohn Foundation, the NRC, the Novo Nordisk Foundation, the University of Bergen, and the Western Norway health Authorities. The Anorexia Nervosa Genetics Initiative (ANGI), an initiative of the Klarman Family Foundation. JK has been supported by the Academy of Finland (grant 336823). This study is funded by “Gentransmed” Centre of Excellence, ERDF nr 2014-2020.4.01.15-0012. MSA is a recipient of a Juan de la Cierva Incorporación contract (IJC2018-035346-I) from the Ministry of Science, Innovation and Universities, Spain. This investigation was supported by Instituto de Salud Carlos III (PI19/00721, P19/01224 and PI20/00041), and cofinanced by the European Regional Development Fund (ERDF), “la Marató de TV3” (092330/31), the Agència de Gestió d’Ajuts Universitaris i de Recerca-AGAUR, Generalitat de Catalunya (2014SGR1357 and 2017SGR1461) and the Pla estratègic de recerca i innovació en salut (PERIS), Generalitat de Catalunya (MENTAL-Cat; SLT006/17/287). This project has also received funding from the European Union’s Horizon 2020 Research and Innovation Programme under the grant agreements No 667302 (CoCA) and 728018 (Eat2beNICE). J.A.R.Q was on the speakers’ bureau and/or acted as consultant for Janssen-Cilag, Novartis, Shire, Takeda, Bial, Shionogi, Sincrolab, Novartis, BMS, Medice, Rubió, Uriach and Raffo in the last 3 years. He also received travel awards (air tickets + hotel) for taking part in psychiatric meetings from Janssen-Cilag, Rubió, Shire, Takeda, Shionogi, Bial and Medice. The Department of Psychiatry chaired by him received unrestricted educational and research support from the following companies in the last 3 years: Janssen-Cilag, Shire, Oryzon, Roche, Psious, and Rubió. Some of the data were collected as part of the following NIH grant: 1R01MH09338. The EPOC study was funded by the German Research Foundation, grants RA1971/8-1, WA731/10-1, WA731/15-1, and KA815/6-1. The GENOS study was supported by the German Research Foundation (GR 1912/1-1). The research at EstBB was supported by the European Union through the European Regional Development Fund (Project No. 2014-2020.4.01.16-0125), the Estonian Research Council grant PUT (PRG184), and through the CoMorMent project. CoMorMent has received funding from the European Union’s Horizon 2020 Research and Innovation Programme under Grant agreement 847776. SEM is supported by an Australian NHMRC Investigator Grant (APP1172917). The AGDS was primarily funded by National Health and Medical Research Council (NHMRC) of Australia grant 1086683. This work was further supported by NHMRC grants 1145645, 1078901 and 1087889. LCC is supported by a QIMR Berghofer Institute fellowship. NORDiC is funded by NIMH R01 MH110427 (PI Crowley), NIMH R01 MH105500 (PI Crowley) and the Swedish Research Council grant 2015–02271 (PI Mataix-Cols). Funding support for some of the Swedish controls was provided by the Klarman Family Foundation, the Swedish Research Council (Vetenskapsrådet, award: 538-2013-8864, and the NIMH Center for Collaborative Genomics Research on Mental Disorders (award U24 MH068457). This study was also supported by NIMH R01MH085321, NIMH R01MH101493, NIMH R01MH58376, and NIMH K20MH01065. It was furthermore funded by NIH R21 MH109938. Support was also provided by the German Research Foundation through grant KA815/6. The Research Council of Norway supported H. Ask, and T. Reichborn-Kjennerud (27611). A. Havdahl was supported by South East Norway Health Authority (2020022). Grant support was also provided from RCN (273291, 262656, 248778, 223273) and the KG Jebsen Stiftelsen. LifeGene was supported by grants from the Ragnar and Torsten Söderberg Foundation, AFA Insurance, the Swedish Research Council and Karolinska Institutet. It is currently a core facility at Karolinska Institutet. The Trøndelag Health Study (HUNT) is a collaboration between HUNT Research Centre (Faculty of Medicine and Health Sciences, Norwegian University of Science and Technology NTNU), Trøndelag County Council, Central Norway Regional Health Authority, and the Norwegian Institute of Public Health. The genotyping was financed by the National Institute of health (NIH), University of Michigan, The Norwegian Research council, and Central Norway Regional Health Authority and the Faculty of Medicine and Health Sciences, Norwegian University of Science and Technology (NTNU). The genotype quality control and imputation has been conducted by the K.G. Jebsen center for genetic epidemiology, Department of public health and nursing, Faculty of medicine and health sciences, Norwegian University of Science and Technology (NTNU). EGOS was supported by a grant from the Beatrice and Samual A. Seaver Foundation to DEG. The OCD Collaborative Genetics Association Study (OCGAS) is a collaborative research study and was funded by the following NIMH Grant Numbers: MH071507 (G N), MH079489 (DAG), MH079487 (JM), MH079488 (AF), and MH079494 (JK). Yao Shugart and Wei Guo were also supported by the Intramural Research Program of the NIMH (MH002930-06).

The International Obsessive Compulsive Foundation Genetics Collaborative (IOCDF-GC) was supported by a grant from the David Judah Foundation (a private, non-industry related foundation established by a family affected by OCD), MH079489 (DLP), MH073250 (DLP), S40024 (JMS), MH 085057 (JMS), and MH087748 (CAM). We are deeply grateful for the participation of all subjects contributing to this research. In Sweden we would further like to thank the local collection team: Anders Juréus, Jessica Pege, Malin Rådström, Radja Satgunanthan-Dawoud, Milka Krestelica, and Birgitta Ohlander, as well as data manager Bozenna Iliadou. We also wish to thank the National Quality Registry for Eating Disorders (RIKSÄT) for help with recruiting patients. We finally wish to thank the BBMRI.se and KI Biobank at Karolinska Institutet for professional biobank service. Data analysis for EstBB was partly carried out in the High-Performance Computing Center of University of Tartu. AGDS would like to thank all the people who helped in the conception, implementation, beta testing, media campaign and data cleaning. They would specifically like to acknowledge Dale Nyholt for advice on using the PBS for research; Ken Kendler, Patrick Sullivan, Andrew McIntosh and Cathryn Lewis for input on the questionnaire; Lorelle Nunn, Mary Ferguson, Lucy Winkler and Natalie Garden for data and sample collection; Natalia Zmicerevska, Alissa Nichles and Candace Brennan for participant recruitment support. Jonathan Davies, Luke Lowrey and Valeriano Antonini for support with IT aspects; Vera Morgan and Ken Kirkby for help with the media campaign. AGDS would like to thank VIVA! Communications for their effort in promoting the study. They also acknowledge David Whiteman and Catherine Olsen from QSkin. The authors thank the Psychiatric Genomics Consortium (PGC) for the use of their servers for data integration and analysis, the many families who have participated in the study, as well as the clinicians, study managers, and clinical interviewers at the respective study sites for their efforts in participant recruitment and clinical assessments. The views expressed here do not reflect the view of the National Institutes of Health, the Department of Health and Human Services, or the United States government.

## WEB RESOURCES

**1000G reference:** www.internationalgenome.org/category/phase-3/

**FUMA:** https://fuma.ctglab.nl

**ggplot2:** www.ggplot2.tidyverse.org

**HapMap3 reference:** www.sanger.ac.uk/resources/downloads/human/hapmap3.html

**Single-cell RNAseq data:** www.github.com/jbryois/scRNA_disease

