## Supplemental Information and Figures (Figures S1-S30) for "Genome-wide association study identifies new locus associated with OCD"

### Supplementary Material - PGC OCD2

**Nora I. Strom[1,2,3,4,182], Dongmei Yu[5,6,182], Zachary F. Gerring[7], Matthew W. Halvorsen[8], Abdel Abdellaoui[9], Cristina Rodriguez-Fontenla[10], Julia M. Sealock[11], Tim Bigdeli[12], Jonathan R. I. Coleman[13,14], Behrang Mahjani[15,16], Jackson G. Thorp[17], Katharina Bey[18], Christie L. Burton[19], Jurjen J. Luykx[20,21], Gwyneth Zai[22,23], Kathleen D. Askland[24], Cristina Barlassina[25], Judith Becker Nissen[26,27], Laura Bellodi[28], O. Joseph Bienvenu[29], Donald Black[30], Michael Bloch[31], Julia Boberg[32], Rosa Bosch[33], Michael Breen[15,34,35], Brian P. Brennan[36], Helena Brentani[37], Joseph D. Buxbaum[15], Jonas Bybjerg-Grauholm[38], Enda M. Byrne[39,40], Beatriz Camarena[41], Adrian Camarena[42], Carolina Cappi[15,37], Angel Carracedo[43], Miguel Casas[44,45], Maria C. Cavallini[46], Valentina Ciullo[47], Edwin H. Cook[48], Vladimir Coric[31], Bernadette A. Cullen[29], Elles J. De Schipper[3], Bernie Devlin[49], Srdjan Djurovic[50,51], Jason A. Elias[36], Lauren Erdman[52], Xavier Estivil[53], Martha J. Falkenstein[54], Bengt T. Fundin[16], Maiken E. Gabrielsen[55], Fernando S. Goes[29], Marco A. Grados[29], Jakob Grove[56,57], Wei Guo[58,59], Jan Haavik[60,61], Kristen Hagen[62,63], Alexandra Havdahl[64,65,66], Ana G. Hounie[37], Donald Hucks[11,67], Christina Hultman[16], Magdalena Janecka[15,34], Michael Jenike[68], Elinor K. Karlsson[69,70], Julia Klawohn[1], Lambertus Klei[71], Janice Krasnow[72], Kristi Krebs[73], Jason Krompinger[36], Nuria Lanzagorta[74], Fabio Macciardi[75], Brion Maher[76], Evonne McArthur[11], Nathaniel McGregor[77], Nicole C. McLaughlin[78], Sandra Meier[79], Euripedes C. Miguel[37], Maureen Mulhern[15,34], Paul S. Nestadt[29], Erika L. Nurmi[80], Kevin S. O'Connell[81,82], Lisa Osiecki[5,83], Teemu Palviainen[84], Fabrizio Piras[47], Federica Piras[47], Ann E. Pulver[29], Raquel Rabionet[53], Alfredo Ramirez[85,86,87,88], Scott Rauch[54], Abraham Reichenberg[89], Jennifer Reichert[15,34], Mark A. Riddle[29], Stephan Ripke[6,90,91], Aline S. Sampaio[37,92], Miriam A. Schiele[93], Laura G. Sloofman[15], Jan Smit[94], Janet L. Sobell[95], María Soler Artigas[96,97,98,99], Laurent F. Thomas[100,101], Homero Vallada[37,102], Jeremy Veenstra-VanderWeele[103], Nienke N.C.C. Vulink[9], Christopher P. Walker[104], Ying Wang[29], Jens R. Wendland[105], Bendik S. Winsvold[106,107,108], Yin Yao[109], Pino Alonso[110], Götz Berberich[111], Cynthia M. Bulik[16,112,113], Danielle Cath[114,115], Daniele Cusi[116], Richard Delorme[117], Damiaan Denys[118], Valsamma Eapen[119], Peter Falkai[120], Thomas V. Fernandez[31], Abby J. Fyer[121,122], Daniel A. Geller[5,123], Hans J. Grabe[124], Benjamin D. Greenberg[77,78,125], Gregory L. Hanna[126], Ian M. Hickie[127], David M. Hougaard[38,57], Norbert Kathmann[1], James Kennedy[23], Liang Kung-Yee[128,129], Mikael Landén[16,130], Stéphanie Le Hellard[131,132], Marion Leboyer[133], Christine Lochner[134], James T. McCracken[80], Sarah E. Medland[7], Preben B. Mortensen[57,135,136], Benjamin Neale[83,137,138], Humberto Nicolini[139,140], Merete Nordentoft[141,142], Michele Pato[143], Carlos Pato[143], David L. Pauls[144], Nancy L. Pedersen[16], John Piacentini[80], Christopher Pittenger[145], Danielle Posthuma[146], Josep A. Ramos-Quiroga[147,148,149,150], Steven A. Rasmussen[78], Kerry J. Ressler[36], Margaret A. Richter[23,151], Maria C. Rosário[152], David R. Rosenberg[153], Stephan Ruhrmann[85], Jack F. Samuels[29], Sven Sandin[15,16], Paul Sandor[23], Gianfranco Spalletta[47,154], Dan J. Stein[155], S Evelyn Stewart[156], Eric A. Storch[154], Barbara E. Stranger[157,158], Maurizio Turiel[159], Thomas Werge[57,142,160,161], Ole A. Andreassen[82,162], Anders D. Børglum[56,57], Susanne Walitza[163,164,165], Bjarne KA. Hansen[131,166], Christian P. Rück[3], Nicholas G. Martin[17], Lili Milani[73], Ole Mors[167], Ted Reichborn-Kjennerud[64,162], Marta Ribasés[97,168,169,170], Gerd Kvale[131,166], David Mataix-Cols[3], Katharina Domschke[93,171], Edna Grünblatt[163,164,165], Michael Wagner[18], John-Anker Zwart[106,107,172], Gerome Breen[13,14], Gerald Nestadt[29], Andres Metspalu[73], Jaakko Kaprio[173], Paul D. Arnold[174,175], Dorothy E. Grice[15], James A. Knowles[176], Helga Ask[64], Karin J.H. Verweij[9], Lea K. Davis[67], Dirk JA. Smit[177], James J. Crowley[3,8,112], Carol A. Mathews[178], Eske M. Derks[17], Jeremiah M. Scharf[5,6,183], and Manuel Mattheisen[4,180,181,183]**

[1-181]see below

**AFFILIATIONS**

[1]Department of Psychology, Humboldt-Universität zu Berlin, Berlin, Germany [2]Departments of Psychiatry, Psychosomatics, and Psychotherapy, University Hospital Würzburg, Würzburg, Germany [3]Department of Clinical Neuroscience, Karolinska Institutet, Stockholm, Sweden [4]Department of Biomedicine, Aarhus University, Aarhus, Denmark [5]Department of Psychiatry, Massachusetts General Hospital, Boston, MA, USA [6]Stanley Center for Psychiatric Research, Broad Institute of MIT and Harvard, Cambridge, MA, USA [7]Department of Mental Health, QIMR Berghofer Medical Research Institute, Brisbane, QLD, Australia [8]Department of Genetics, University of North Carolina At Chapel Hill, Chapel Hill, NC, USA [9]Department of Psychiatry, Amsterdam UMC, University of Amsterdam, Amsterdam, The Netherlands [10]CIMUS (centre for Research In Molecular Medicine and Chronic Diseases), University of Santiago De Compostela, Santiago De Compostela, A Coruña, Spain [11]Vanderbilt Genetics Institute, Vanderbilt University Medical Center, Nashville, TN, USA [12]Departments of Psychiatry and Behavioral Sciences, SUNY Downstate Health Sciences University, Brooklyn, NY, USA [13]Social, Genetic and Developmental Psychiatry Centre, King's College London, London, United Kingdom [14]Departments of South London and Maudsley NHS Trust, NIHR Maudsley Biomedical Research Centre, London, United Kingdom [15]Department of Psychiatry, Icahn School of Medicine At Mount Sinai, New York, NY, USA [16]Departments of Medical Epidemiology and Biostatistics, Karolinska Institutet, Stockholm, Sweden [17]Departments of Genetics and Computational Biology, QIMR Berghofer Medical Research Institute, Brisbane, QLD, Australia [18]Departments of Psychiatry and Psychotherapy, University Hospital Bonn, Bonn, Germany [19]Departments of Neurosciences and Mental Health, Hospital for Sick Children, Toronto, ON, Canada [20]Department of Psychiatry, University Medical Center Utrecht, Utrecht, The Netherlands [21]Second Opinion Outpatient Clinic, Ggnet, Warnsveld, The Netherlands [22]Molecular Brain Science Department, Campbell Family Mental Health Research Institute, Centre for Addiction and Mental Health, Toronto, ON, Canada [23]Department of Psychiatry, University of Toronto, Toronto, ON, Canada [24]Waypoint Research Institute AND Outpatient Assessment and Treatment Services,, Waypoint Centre for Mental Health Care, Penetanguishene, ON, Canada [25]Department of Heath Sciences, University of Milano, Milano, Milano, Italy [26]Departments of Child and Adolescent Psychiatry, Aarhus University Hospital, Psychiatry, Denmark, Aarhus University Hospital, Psychiatry, Aarhus, Denmark [27]Institute of Clinical Medicine, Health, Aarhus University, Health, Aarhus University, Aarhus, Danmark [28]Department of Neuropsychiatric Sciences, Università Vita-salute San Raffaele Milano Italy, Milano, Italy [29]Departments of Psychiatry and Behavioral Sciences, Johns Hopkins University, Baltimore, MD, USA [30]Departments of Roy J. and Lucille A. Carver College of Medicine, University of Iowa, Iowa City, IA, USA [31]Child Study Center and Psychiatry, Yale University, New Haven, CT, USA [32]Center for Psychiatric Research, Institution of Clinical Neuroscience, Stockholm, Sweden [33]Department of MIND SCHOOLS, HOSPITAL SANT JOAN DE DEU, ESPLUGUES DE LLOBREGAT, BARCELONA, Spain [34]Seaver Autism Center for Research and Treatment, Icahn School of Medicine At Mount Sinai, New York, NY, USA [35]The Mindich Child Health and Development Institute, Icahn School of Medicine Mount Sinai, New York, NY, USA [36]Department of Mclean Hospital, Harvard Medical School, Belmont, MA, USA [37]Department of Psychiatry, Universidade De São Paulo, São Paulo, Brazil [38]Department of Congenital Disorders, Statens Serum Institut, Copenhagen, Denmark [39]Institute for Molecular Bioscience, University of Queensland, Brisbane, Queensland, Australia [40]Child Health Research Centre, University of Queensland, Brisbane, QLD, Australia [41]Department of Pharmacogenetics, Instituto Nacional De Psiquitria Ramon De La Fuente Muñiz, Mexico City, Mexico [42]Department of Surgery, Duke University, Durham, NC, USA [43]Genomics Group, University of Santiago De Compostela, Santiago De Compostela, A Coruña, Spain [44]Programa MIND Es-

DRAFT

coles, Hospital Sant Joan De Déu, ESPLUGUES DE LLOBREGAT, Barcelona, Spain [45]Department of Departamento De Psiquiatría Y Medicina Legal, Universitat Autònoma De Barcelona, Bellaterra, Barcelona, Spain [46]Department of Psychiatry, Ospedale San Raffaele, Milano, Italy [47]Laboratory of Neuropsychiatry, IRCCS Santa Lucia Foundation, Rome, Italy [48]Department of Psychiatry, University of Illinois At Chicago, Chicago, IL, USA [49]Department of Psychiatry, University of Pittsburgh School of Medicine, Pittsburgh, PA, USA [50]Department of Medical Genetics, Oslo University Hospital, Oslo, Norway [51]NORMENT, Clinical Science, University of Bergen, Bergen, Norway [52]Departments of Genetics and Genome Biology, The Hospital for Sick Children, Toronto, ON, Canada [53]Department of Genetics, University of Barcelona, Barcelona, Spain [54]Department of Psychiatry, Mclean Hospital, Harvard Medical School, Belmont, MA, USA [55]Departments of Public Health and Nursing, Norwegian University of Science and Technology, Trondheim, Norway [56]Biomedicine and The iSEQ Center, Aarhus University, Aarhus, Denmark [57]The Lundbeck Foundation Initiative for Integrative Psychiatric Research, Ipsych, , Denmark [58]National Institute of Mental Health, National Institutes of Health, Bethesda, MD, USA [59]Genetic Epidemiology Research Branch, National Institute of Mental Health, National Institutes of Health, Bethesda, MD, Bethesda, MD, USA [60]Department of Biomedicine, University of Bergen, Bergen, Norway [61]Department of Division of Psychiatry, Haukeland University Hospital, Bergen, Norway [62]Department of Molde Hospital, Møre Og Romsdal Hospital Trust, Molde, Norway [63]Department of Mental Health, Norwegian University of Science and Technology, Trondheim, Norway [64]Department of Mental Disorders, Norwegian Institute of Public Health, Oslo, Norway [65]Nic Waals Institute, Lovisenberg Diaconal Hospital, Oslo, Norway [66]PROMENTA Research Center, Psychology, University of Oslo, Oslo, Norway [67]Department of Medicine, Vanderbilt University Medical Center, Nashville, TN, USA [68]Department of Psychiatry, Massachusetts General Hospital and Harvard Medical School, Boston, MA, USA [69]Departments of Bioinformatics and Integrative Biology, University of Massachusetts Medical School, Worcester, MA, USA [70]Department of Vertebrate Genomics, Broad Institute of MIT and Harvard, Cambridge, MA, USA [71]Department of Psychiatry, University of Pittsburgh, Pittsburgh, PA, USA [72]Department of Psychiatry, Johns Hopkins University, Baltimore, MD, USA [73]Institute of Genomics, University of Tartu, Tartu, Estonia [74]Carracci Medical Group, Mexico City, Mexico [75]Departments of Psychiatry and Human Behavior, University of Californiairvine (UCI), Irvine, CA, USA [76]Department of Mental Health, Johns Hopkins Bloomberg School of Public Health, Baltimore, MD, USA [77]COBRE Center for Neuromodulation, Butler Hospital, Providence, RI, USA [78]Departments of Psychiatry and Human Behavior, Alpert Medical School, Brown University, Providence, RI, USA [79]Department of Psychiatry, Dalhousie University, Halifax, NS, Canada [80]Departments of Psychiatry and Biobehavioral Sciences, University of California, Los Angeles, Los Angeles, CA, USA [81]NORMENT Center of Excellence (coe), Institute of Clinical Medicine, University of Oslo, Oslo, Norway [82]Departments of Division of Mental Health and Addiction, Oslo University Hospital, Oslo, Norway [83]Psychiatric and Neurodevelopmental Genetics Unit, Harvard Medical School, Boston, MA, USA [84]Institute for Molecular Medicine Finland - FIMM, University of Helsinki, Helsinki, Finland [85]Departments of Psychiatry and Psychotherapy, University of Cologne, Cologne, Germany [86]Departments of Neurodegenerative Diseases and Geriatric Psychiatry, University of Bonn, Bonn, Germany [87]DZNE Bonn, German Center for Neurodegenerative Diseases (DZNE), Bonn, Germany [88]Psychiatry and Glenn Biggs Institute for Alzheimer's and Neurodegenerative Diseases, UT Health San Antonio, San Antonio, TX, USA [89]Department of Mental Disorders, Norwegian Institute of Public Health, New York, NY, USA [90]Department of Medicine, Massachusetts General Hospital and Harvard Medical School, Boston, MA, USA [91]Departments of Psychaitry and Psychotherapy, Charité Universitätsmedizin, Berlin, Germany [92]Department of University Health Careservices - SMURB, Federal University of Bahia, Salvador, Brazil [93]Departments of Psychiatry and Psychotherapy, Medical Center - University of Freiburg, Freiburg, Germany [94]Department of Psychiatry, Amsterdam UMC Location Vumc, Amsterdam, The Netherlands [95]Departments of Psychiatry and The Behavioral Sciences, Keck School of Medicine of USC, Los Angeles, CA, USA [96]Psychiatric Genetics Unit, Group of Psychiatry, Mental Health and Addiction, Vall D'hebron Research Institute (VHIR), Barcelona, Spain [97]Department of Psychiatry, Hospital Universitari Vall D'hebron, Barcelona, Spain [98]Department of Instituto De Salud Carlos III, Biomedical Network Research Centre On Mental Health (CIBERSAM), Madrid, Spain [99]Department of Biology, Universitat De Barcelona, Barcelona, Spain [100]Departments of Clinical and Molecular Medicine, Norwegian University of Science and Technology, Trondheim, Norway [101]K. G. Jebsen Center for Genetic Epidemiology, Norwegian University of Science and Technology, Trondheim, Norway [102]Departments of Molecular Medicine and Surgery, Karolinska Institutet, Stockholm, Sweden [103]Departments of Psychiatry, Pediatrics, and Pharmacology, Vanderbilt University Medical Center, Nashville, TN, USA [104]Department of Precision Medicine, City of Hope, Monrovia, CA, USA [105]Laboratory of Clinical Science, NIMH Intramural Research Program, Bethesda, MD, USA [106]Departments of Research, Innovation and Education, Oslo University Hospital, Oslo, Norway [107]Departments of Medicine and Health Sciences, Norwegian University of Science and Technology, Trondheim, Norway [108]Department of Neurology, Oslo University Hospital, Oslo, Norway [109]Department of Computional Biology,, Fudan University., Fudan, PRC [110]Department of Bellvitge University, University of Barcelona, Barcelona, Spain [111]Psychosomatic Department, Windach Hospital of Neurobehavioural Research and Therapy, Windach, Germany [112]Department of Psychiatry, University of North Carolina At Chapel Hill, Chapel Hill, NC, USA [113]Department of Nutrition, University of North Carolina At Chapel Hill, Chapel Hill, NC, USA [114]Departments of Rijksuniversiteit Groningen and Psychiatry, University Medical Center Groninge, Groningen, The Netherlands [115]Department of Specialized Training, Drenthe Mental Health Care Institute, , [116]Institute of Biomedical Technologies, Italian National Centre for Research, Segrate, Milano, Italy [117]Child and Adolesccent Psycchiatry Department, APHP, Paris, France [118]Department of Psychiatry, Institute of The Royalnetherlands Academy of Arts and Sciences (NIN-KNAW), Amsterdam, The Netherlands [119]Department of Medicine, University of New South Wales, Randwick, NSW, Australia [120]Departments of Psychiatry and Psychotherapy, University of Göttingen, Göttingen, Germany [121]Department of Psychiatry, New York State Psychiatric Institute, New York, NY, USA [122]Department of Psychiatry, Columbia University Medical Center, New York, NY, USA [123]Department of Psychiatry, Harvard Medical School, Boston, MA, USA [124]Departments of Psychiatry and Psychotherapy, University Medicine Greifswald, Greifswald, Greifswald [125]RRD Center for Neurorestoration and Neurotechnology, Providence VA Medical Center, Providence, RI [126]Department of Psychiatry, University of Michigan, Ann Arbor, MI, USA [127]Brain and Mind Center, The University of Sydney, Sydney, NSW, Australia [128]Department of Biostatistics, Johns Hopkins Bloomberg School of Public Health, Baltimore, MD, USA [129]Institute of Population Health Sciences, National Health Research Institutes, , Taiwan [130]Institute of Neuroscience and Physiology, University of Gothenburg, Gothenburg, Sweden [131]Bergen Center for Brain Plasticity, Haukeland University Hospital, Bergen, Norway [132]Department of Clinical Science, University of Bergen, Bergen, Norway [133]Departments of Addictology and Psychiatry, Univ Paris Est Créteil, AP-HP, Inserm, Paris, France [134]Department of Psychiatry, Stellenbosch University, Stellenbosch, South Africa [135]National Centre for Register-based Research, Aarhus University, Aarhus, Denmark [136]Centre for Integrated Register-based Research, Aarhus University, Aarhus, Denmark [137]Analytic and Translational Genetics Unit, Massachusetts General Hospital, Boston, MA, USA [138]Program In Medical and Population Genetics, Broad Institute of Harvard and MIT, Cambridge, MA, USA [139]Department of Psychiatric Genetics, Grupo Médico Carracci INMEGEN, Mexico City, México [140]Department of Psychiatric Genetics, Instituto Nacional De Medicina Genómica, México City, Mexico [141]CORE - Copenhagen Research Center for Mental Health, Copenhagen University Hospital, Copenhagen, Denmark [142]Department of Clinical Medicine, University of Copenhagen, Copenhagen, Denmark [143]Department of Psychiatry, Rutgers University, Piscataway, NJ, USA [144]Department of Psychiatry, Harvard University, Boston, MA, USA [145]Department of Psychiatry, Yale University, New Haven, CT, USA [146]Department of Clinical Genetics, VU University Medical Center Amsterdam, Amsterdam, The Netherlands [147]Department of Psychiatry, Hospital Universitari Vall D'hebron, Barcelona, Catalonia, Spain [148]Group of Psychiatry, Mental Health and Addictions, Vall D'hebron Research Institute (VHIR), Barcelona, Spain [149]Group 27, Biomedical Network Research Centre On Mental Health (CIBERSAM), Barcelona, Spain [150]Departments of Psychiatry and Forensic Medicine, Universitat Autònoma De Barcelona, Barcelona, Spain [151]Department of Psychiatry, Sunnybrook Health Sciences Centre, , [152]Department of Psychiatry, Federal University of São Paulo (UNIFESP), São Paulo, Brazil [153]Departments of Psychiatry and Behavioral Neurosciences, Wayne State University School of Medicine, Detroit, MI, USA [154]Departments of Psychiatry and Behavioral Sciences, Baylor College of Medicine, Houston, TX, USA [155]Psychiatry and Neuroscience Institute, University of Cape Town, Cape Town, Western Cape, South Africa [156]Department of Psychiatry, University of British Columbia, Vancouver, BC, Canada [157]Department of Pharmacology, Northwestern University Feinberg School of Medicine, Chicago, IL, USA [158]Center for Genetic Medicine, Northwestern University Feinberg School of Medicine, Chicago, IL, USA [159]Department of Cardiology, University of Milan, Milan, Italy [160]Department of Mental Health Services, Copenhagen University Hospital, Copenhagen, Denmark [161]GLOBE Institute, University of Copenhagen, Copenhagen, Denmark [162]Institute of Clinical Medicine, University of Oslo, Oslo, Norway [163]Departments of Child and Adolescent Psychiatry and Psychotherapy, University of Zurich, Zürich, Switzerland [164]Neuroscience Center Zurich, University of Zurich and The ETH Zuric, Zurich, Switzerland [165]Zurich Center for Integrative Human Physiology, University of Zurich, Zurich, Switzerland [166]Department of Psychology, University of Bergen, Bergen, Norway [167]Psychosis Reasearch Unit, Aarhus University Hospital - Psychiatry, 8200 Aarhus N, Denmark [168]Psychiatric Genetics Unit, Vall D'hebron Research Institute (VHIR), Universitat Autònoma De Barcelona, Barcelona, Spain [169]Biomedical Network Research Centre On Mental Health (CIBERSAM), Barcelona, Spain. 3 Biomedical Network Research Centre On Mental Health (CIBERSAM), Instituto De Salud Carlos III, Madrid, Spain [170]Departments of Genetics, Microbiology and Statistics, University of Barcelona, Barcelona, Spain [171]Center for Basics In Neuromodulation, University of Freiburg, Freiburg, Germany [172]Department of Medicine, University of Oslo, Oslo, Norway [173]Helsinki Institute of Life Science, University of Helsinki, Helsinki, Finland [174]Department of Psychiatry, Cumming School of Medicine, University of Calgary, Calgary, AB, Canada [175]Program In Genetics and Genome Biology, Hospital for Sick Children, Toronto, ON, Canada [176]Department of Cell Biology, SUNY Downstate Health Sciences University, Brooklyn, NY, USA [177]Department of Psychiatry, Amsterdam UMC Location AMC, Amsterdam, The Netherlands [178]Psychiatry and Genetics Institute, Center for OCD, Anxiety and Related Disorders, University of Florida, Gainesville, FL, USA [179]Department of Psychiatry, Center for Genomic Medicine, Massachusetts General Hospital, Harvard Medical School, Boston, MA, USA [180]Departments of Psychiatry and Community Health and Epidemiology, Dalhousie University, Halifax, NS, Canada [181]Institute of Psychiatric Phenomics and Genomics (IPPG), University Hospital of Munich, Munich, Germany [182]Contributed equally [183]Contributed equally

### Methods

#### Sample Descriptions of individual participating studies

DRAFT

In the following we describe each individual sample that was included in the OCD meta-analysis. The header of each sample lists study identifier, study PI(s), country or site name, and if there has been a previous publication in connection with the data, also PubMed ID(s). See Supplementary Table S1 for an overview of sample sizes, number of included SNPs, reference panel and GWAS analysis tool used, and Lambda1000 estimate for each individual cohort.

*OCGAS_all | Multiple | USA | 22889921*

Results of *The OCD Collaborative Genetics Association Study* (OCGAS) have been published previously[1]. For detailed sample- and analysis description refer to the primary article. In brief: The OCGAS sample consists of 986 cases from OCD Collaborative Genetics Association Study (OCGAS)[1?] , a family based sample, and 1023 controls from Genomic Psychiatry Cohort (GPC)[2], a population based sample. The inclusion of control samples was slightly altered in this publication compared to the original study. For study inclusion, probands were required to meet DSM-IV criteria for OCD with onset of obsessions and/or compulsions before the age of 18 years (*mean*= 9.4 years; *SD* = 6.35), as evaluated by a PhD-level clinical psychologist using the Structured Clinical Interview for DSM-IV modified and extended to include additional symptom an diagnostic information. Several mental- and brain disorders were reason for exclusion. Genotyping was performed at the Johns Hopkins SNP Center using Illumina's HumanOmniExpress bead chips (Illumina, San Diego, CA, USA). A stringent quality control protocol was followed, including checking the relatedness of samples, sex-comparison, Mendelian inconsistencies etc. One sample was removed per related pairs with pihat>0.2. Multidimensional scaling analyses were performed on singleton OCD cases and unselected controls, as implemented in PLINK. Samples were removed when they significantly deviated in the first two multi- dimensional scaling dimensions (> 4 *SD* from the mean). The genotyped data were phased by SHAPEIT2[3] and imputed by Minimac3[4] using the HRC release 1.1 as the reference panel. GWAS was performed on SNPs with info score >0.8 and MAF>0.01, using logistic regression model in PLINK2[5] with the first four and the 9th MDS components as covariates. Ethics approvals for the OCGAS study were obtained from the Hopkins Medicine Institutional Review Boards, the Butler Institutional Review Board, the UCLA Institutional Review Boards, the Mass General Brigham Human Research Committee, the Columbia University Institutional Review Boards, and the National Institutes of Health Institutional Review Board (NIH IRB).

*IOCDF-GC and 610k_trio | Multiple | Multiple | 22889921, 28761083*

The results of the International OCD Foundation-Genetics Consortium (IOCDF-GC) study was previously published[6;7]. IOCDF-GC case-control cohort (IOCDF-GC) consists of 1519 European ancestry cases and 3541 matching controls from IOCDF-GC and three cohorts previously genotyped, including the Alzheimer's Disease Genetics Initiative[8], the Center for Applied Genomics (CAG) at Children's Hospital of Philadelphia (CHOP)[9], and the Breast and Prostate Cancer Cohort Consortium (BPC3)[10]. The IOCDF-GC trio sample (610k_trio) consist of 323 European ancestry complete trios. All cases and trios were recruited predominantly from OCD specialty clinics, and controls were recruited from Bonn, Germany and from Capetown, South Africa. This work was approved by the relevant IRBs at all participating sites, and all participants provided written informed consent. For study inclusion, all cases and trio probands were required to have a DSM-IV diagnosis of OCD. The controls from Bonn had an absent lifetime history of all axis I disorders and the South African controls were diagnostically unscreened. The sample description of the three cohorts refer to the primary article. All samples were genotyped on Illumina Human610-Quadv1_B SNP array (Illumina, San Diego, CA, USA). Standard quality control (QC) protocol was conducted with PLINK2[5]. Samples were removed for call rates<98%, sex discrepancy and ambiguous genomic sex, related samples with pihat>0.2. SNP QC included removing monomorphic SNPs, CNV-targeted SNP probes, SNPs with genotyping rate<98%, SNPs with minor allele frequency (MAF)<0.01, strand-ambiguous SNPs with significant allele frequency differences or aberrant LD correlations with adjacent SNPs based on the entire HapMap2 reference panel, SNPs with $P$<1x$10^{-6}$ in Hardy Weinberg Equilibrium (HWE) test among controls or $P$<1x$10^{-10}$ among cases, SNPs with differential missing rate between cases and controls (> 0.02), and SNPs with batch effect (P<1x$10^{-5}$) between different control cohorts. Multidimensional scaling (MDS) analyses were performed in PLINK2 and samples were removed when they were significant outliers in the first five MDS dimensions or when there were no matching cases or controls on these MDS dimensions. The genotyped data were phased by SHAPEIT2[3] and imputed by Minimac3[4] using the HRC release 1.1 as the reference panel. GWAS was performed on SNPs with info score>0.8 and MAF>0.01, using logistic regression model in PLINK2 with the first five and the 7th MDS components as covariates. Same QC was conducted on the 323 trios, with additional filter of removing SNPs with Mendelian errors. In each trio, the transmitted alleles and untransmitted alleles were converted into one case and one pseudo-control. Phasing and imputation were conducted on the cases and pseudo-controls in the same way as the case-control cohort. GWAS was performed without covariates.

*Psych_Broad | Multiple | Netherlands, Italy, USA, Spain*

Psych_Broad sample consists of 1396 European ancestry cases and 4009 population matched controls. The cases were recruited predominantly from OCD specialty clinics in US, Spain, The Netherlands, and Italy. The Spanish controls were part of the Mental-Cat clinical sample or the IN-School population-based cohort. Both studies have been approved by the Clinical Research Ethics Committee (CREC) of Hospital Universitari Vall d'Hebron. A total of 1,757 controls from the Mental-Cat cohort (60.3% males) were evaluated and recruited prospectively from a restricted geographic area at the Hospital Universitari Vall d'Hebron of

DRAFT

Barcelona (Spain) and consisted of unrelated healthy blood donors. The INSchool sample consisting of 765 children (76.2% males) from schools in Catalonia. Genomic DNA samples were obtained either from peripheral blood lymphocytes by the salting out procedure or from saliva using the Oragene DNA Self-Collection Kit (DNA Genotek, Kanata, Ontario Canada). DNA concentrations were determined using the Pico- Green dsDNA Quantitation Kit (Molecular Probes, Eugene, OR) . The study was approved by the Clinical Research Ethics Committee (CREC) of Hospital Universitari Vall d'Hebron, all methods were performed in accordance with the relevant guidelines and regulations and written informed consent was obtained from participant parents before inclusion into the study. Detailed information has been published previously (doi: 10.1038/s41386-020-0664-5 and DOI: 10.1017/S0033291720005115). All cases were required to have a DSM-IV diagnosis of OCD. The population based unscreened controls were recruited from the same countries. This work was approved by the relevant IRBs at all participating sites, and all participants provided written informed consent. All samples were genotyped on Infinium PsychArray-24 at Broad Institute (Cambridge, MA, USA). Standard quality control (QC) protocol was conducted with PLINK2[5]. Samples were removed for call rates<98%, sex discrepancy and ambiguous genomic sex, related samples with pihat>0.2. SNP QC included removing monomorphic SNPs, SNPs with genotyping rate<98%, SNPs with minor allele frequency (MAF)<0.01, SNPs with $P<1\text{x}10^{-6}$ in HWE test among controls or $P<1\text{x}10^{-10}$ among cases, SNPs with differential missing rate between cases and controls (> 0.02), and SNPs with batch effect ($P<1\text{x}10^{-5}$). Multidimensional scaling (MDS) analyses were performed in PLINK2 and samples were removed when they were significant outliers in the first six MDS dimensions or when there were no matching cases or controls on the MDS dimensions. The genotyped data were phased by SHAPEIT2[3] and imputed by Minimac3[4] using the HRC release 1.1 as the reference panel. GWAS was performed on SNPs with info score>0.8 and MAF>0.01, using logistic regression model in PLINK2 with the first six and the 8th MDS PCA components as covariates. The GWAS result of this data set has not been previously published.

*AUS | Derks, E. | QIMR, Brisbane, Australia*

The *Australian Genetics of Depression study* (AGDS) was established to recruit a large cohort of individuals who have been diagnosed with depression at some point in their lifetime. The purpose of establishing this cohort is to investigate genetic and environmental risk factors for depression and response to commonly prescribed antidepressants. For the present study, we combined information from the *Obsessive-Compulsive Inventory-Revised* (OCI-R) with self-reported clinical diagnosis to identify cases (diagnosed with depression and OCD) and controls (diagnosed with depression). A total of 20,689 participants were recruited through the Australian Department of Human Services and a media campaign, 75% of whom were female. Participants were recruited to the Australian Genetics of Depression Study (www.geneticsofdepression.org.au) between 2016 and 2019. All study protocols were approved by the QIMR Berghofer Medical Research Institute Human Research Ethics Committee. The protocol for approaching participants through the DHS, enrolling them in the study, and consenting for all phases of the study (including invitation to future related studies) and accessing MBS and PBS records was approved by the Ethics Department of the Department of Human Services. The average age of participants was 43 years ± 15 years. Participants completed an online questionnaire that consisted of a compulsory module that assessed self-reported psychiatric history, clinical depression using the *Composite Interview Diagnostic Interview Short Form*, and experiences of using commonly prescribed antidepressants. Further voluntary modules assessed a wide range of traits of relevance to psychopathology. Participants who reported they were willing to provide a DNA sample (75%) were sent a saliva kit in the mail. In the present study, we included 757 cases and 5368 controls for whom genotype data were available. Cases were defined by self-reported diagnosis of OCD. Controls were defined by no self-reported diagnosis of OCD plus a sumscore less than 10 on the Obsessive Compulsive Inventory-Revised (OCI-R). Samples from the AGDS were genotyped in three different genotyping centers using the same array (GSAMD-24v1-0_20011747). Genotype calling was performed with GenomeStudio. A common set of high QC markers between the different genotyping batches was obtained prior to joint imputation. Marker exclusion criteria (prior to imputation) included: unknown or ambiguous map position and strand alignment in a BLAST search, missingness>5%, HWE test $P< 10^{-6}$, MAF<1%, GenTrain score<0.6. The Michigan imputation server was used to impute the genotypes using the HRCr1.1 as a reference panel. Individuals were excluded based on a high missingness (missing rate>3%), inconsistent (and unresolvable) sex, or if deemed ancestry outliers from the European population (6 standard deviations from the first two genetic principal components). The GWAS was done employing a logistic regression using PLINK 1.9 and imputed dosage genotypes while correcting for the genotyping center and the first twenty ancestry principal components as covariates.

*iPSYCH | Borglum, A.D.; Mors, O.; Mattheisen, M. | Denmark*

In the scope of the *Danish OCD and Tourette Study* (DOTS) within *The Lundbeck Foundation Initiative for Integrative Psychiatric Research* (iPSYCH), Danish nation-wide population-based case-cohort samples were collected and genotyped. The study was approved by the *Regional Scientific Ethics Committee* in Denmark and the *Danish Data Protection Agency*. All analyses of the samples were performed on the secured national GenomeDK high performance-computing cluster in Denmark (https://genome.au.dk). See Pedersen et al.[11] for a detailed description of the overall cohort, array genotyping and quality control. The iPSYCH sample comprises 2938 individuals with a diagnosis of OCD. Eligible were singletons that had been a Danish resident on their first birthday. All OCD patients that are included in the iPSYCH sample are either comorbid with one of the

DRAFT

primary disorders in iPSYCH or stem from the population based pool of controls. Genetic information was obtained by the Statens Serum Institut (SSI) at the Danish Neonatal Screening Biobank (DNSB) from heel prick blood samples that had been collected from all newborn babies in Denmark. Genotyping was performed on the PsychChip v 1.0 array (Illumina, San Diego, CA, USA) at the Broad Institute of MIT and Harvard (Cambridge, MA, USA). Genotyping and data processing was carried out in 25 waves. Genetic information was coupled with other registers via the Danish Civil Registration System (CPR). Cases were identified by the Danish Psychiatric Central Research Register (DPCRR) which collects data on all individuals treated in Denmark either in psychiatric hospitals or in outpatient psychiatric clinics that met ICD10 (F42) criteria. Controls were randomly selected from the same birth cohorts, and excluded individuals with a diagnosis of F42. Genotypes were processed using the *Rapid Imputation and COmputational PIpeLIne for Genome-Wide Association Studies* (Ricopili; see https://sites.google.com/a/broadinstitute.org/ricopili/) performing stringent quality control of the data. Samples with call rates below 95% and individuals with a mismatch between sex obtained from genotyping and registered sex in the DPCRR were excluded. Related individuals were removed, principle component analyses were used to exclude ancestral outliers and the data was imputed using the HRC reference panel. The final dataset included 2678 cases and 10410 controls.

*EPOC | Wagner, M.; Kathmann, N. | Germany | 30744714, 30008679, 29890378, 29721727, 29159055, 28541065, 28481032, 28160276*

The EPOC (Endophenotypes of OCD) sample comprises (epi)genetic and deep phenotype data of OCD patients, unaffected first-degree relatives of OCD patients and healthy controls that were collected at two sites in Germany (Berlin and Bonn) between 2014 and 2017. In the present analysis, data from 195 patients with OCD (55.9 % female, 44.1 % male) and 204 controls (63.2 % female, 36.8 % male) were included. Mean age was 33.37 (*SD* = 10.76; *range*: 18–64) for patients and 34.72 (*SD* = 12.64; *range*: 18–64) for controls. Lifetime comorbidity rates of patients were 60.0% for depression; 11.3% for panic disorder or agoraphobia; 7.7% for tic disorder; 7.7% for specific phobia; 6.7% for social phobia; 4.6% for generalized anxiety disorder; 4.1% for PTSD; 4.1% for anorexia; 3.1% for hypochondria; and 2.6% for hoarding disorder. OCD patients were recruited via the outpatient clinics at the Department of Psychology of Humboldt University, Berlin and at the Department of Psychiatry and Psychotherapy of the University Hospital Bonn. Healthy volunteers were recruited from the general population via public advertisements in the same cities. All participants were examined by trained clinical psychologists using the Structured Clinical Interview for DSM-IV (SCID-I) to assess OCD diagnosis and potential comorbidities. To establish cross-site reliability of clinical ratings, all instructions were standardized and raters completed assessments of four training videos. Patients were only included if they: (a) met diagnostic criteria of OCD based on the SCID-I interview; (b) were free of past or present psychotic, bipolar or substance-related disorders; (c) did not take neuroleptic medication for the previous four weeks; and (d) did not use benzodiazepines in the prior two weeks. Healthy controls were excluded if they: (a) took any psychoactive medication in the previous three months; (b) had a current Axis I disorder; (c) had a lifetime diagnosis of OCD or tic disorder; or (d) had a family history of OCD. Written informed consent was obtained and participants were compensated for their time. The study was in accordance with the revised Declaration of Helsinki and approved by the local ethics committees of the Charité University Medicine Berlin and the University Hospital Bonn. Genotyping of cases and controls was performed on the Illumina Global Screen Array (GSA) at the Life Brain Center, Bonn. Genotype quality control was done using Plink-1.9, and R (version 3.5.1). We checked the data for sex inconsistencies and grossly failing markers (call rate$<$0.5). Individuals with a call rate of$<$0.95 were removed. The heterozygosity rate for each subject was calculated; outliers ($\pm$3 SD from the mean heterozygosity rate) were identified and removed. On marker level, SNPs were removed if at least one of the following conditions was true: significant difference of missing rate between cases and controls, call rate$<$0.95; deviation of Hardy-Weinberg equilibrium ($p<1x10^{-6}$); and minor allele frequency$<$0.05 (computed separately in cases and controls). Furthermore, all A/T or C/G SNPs were removed. To check for population stratification, principal component analysis (PCA) was performed, and the first two principal components were checked for outliers. Seven individuals, whose non-European ethnicity was additionally validated based on their demographic data, were excluded. The genotyped data were imputed on the Michigan Imputation Service using the 1000 Genomes Phase 3 (version 5) reference panel. GWAS was conducted using SNPTEST (version 2.5.2) with the first four PCA components as covariates.

DRAFT

*FinnGen | Kaprio, J. A. | Finland*

Finnish samples are population-based biobank samples collected between August 2017 and August 2019 (collection further ongoing), including legacy samples collected since the 1980s. The Ethical Review Board of the Hospital District of Helsinki and Uusimaa approved the FinnGen study protocol Nr. HUS/990/2017. The FinnGen project was approved by Finnish Institute for Health and Welfare (THL), approval numbers THL/2031/6.02.00/2017, amendments THL/341/6.02.00/2018, THL/2222/6.02.00/2018 and THL/283/6.02.00/2019. The here included data has not been published before but a general description of the FinnGen study can be found elsewhere[12]. Cases were recruited through hospital records (inpatient and outpatients) from 1970 onwards, based on clinical diagnoses used in patient care, supplemented by cause of death diagnoses (if clinical diagnosis was underlying and/or contributing cause). Case definitions were based on a diagnosis of ICD-10 F42 and ICD-8 3003, there were no case exclusion criteria. The mean age of cases was 45.36 (*SD* = 15.44) years with a minimum age of 16.5 and a maximum age of 91.4, including 330 males and 460 females. Controls were defined as all other participants in FinnGen with GWAS data. The mean age of controls was 60.02 (*SD* = 17.28 years with a minimum age of 0 and a maximum age of 105.73, including 71252 males and 90138 females. Genotyping of cases and controls was performed in multiple batches on multiple arrays in multiple centres. For genotype quality control the following filters were applied to the data: exclusion of chromosome X, exclusion of variants with INFO score<0.95, with missingness>0.01, or with MAF<0.05. To check for population stratification, principal component analysis (PCA) was performed and a bayesian algorithm was used to spot outliers. Individuals were excluded if there was a mismatch between imputed sex and sex in registry data. For calculating the genetic relationship matrix, we used the genotype dataset where genotypes with GP<0.95 have been set missing. Only variants imputed with an INFO score>0.95 in all batches were used. Variants with>3% missing genotypes were excluded as well as variants with MAF<1%. The remaining variants were LD pruned with a 1Mb window and $r^2$ threshold of 0.1. Imputation was performed using Eagle 2.4/Beagle 4.1 using Finnish WGS (depth up to 30x) samples as reference with a total amount of 16962023 variants. GWAS was performed using SAIGE (v0.35.8.8) including age, sex, the first 10 PCs, genotyping batch and kinship matrix as covariates.

*HUNT | Zwart, J.-A. | Norway*

The Trøndelag Health Study (HUNT) consists of three different population-based health surveys conducted in the county of Nord-Trøndelag, Norway over approximately 20 years (HUNT1: 1984-1986, HUNT2: 1995-1997 and HUNT3: 2006-2008). The HUNT study was approved by the Regional Committee for Medical and Health Research Ethics, Norway (2015/575). For each survey, the entire adult population ($\geq$ 20 years) was invited to participate by completing questionnaires, attending clinical examinations and interviews. Participation rates in HUNT1, HUNT2 and HUNT3 were 89.4% (*N* = 77,212), 69.5% (*N* = 65 237) and 54.1% (*N* = 50 807), respectively. Taken together, the study included more than 120,000 different individuals from Nord-Trøndelag County. Biological samples including DNA have been collected for approximately 70000 participants. The entire HUNT Study has been described in more detail elsewhere[13]. For the present study, we included participants from HUNT2 and HUNT3. Cases and controls were defined by linkage to hospital diagnostic registries from the time period 1987-2017. Cases were defined as those with a hospital diagnosis of obsessive-compulsive disorder (ICD-10 code F42). Controls were defined as those without ICD-10 code F42 and without ICD-9 code 300 (“anxiety, dissociative and somatoform disorders”). The study was approved by the Regional Committee for Medical and Health Research Ethics (ref. 2015/575). In total, DNA from 71,860 HUNT samples was genotyped at the Genomics Core Facility at the Norwegian University of Science and Technology using one of three different Illumina HumanCoreExome arrays (HumanCoreExome12 v1.0, HumanCoreExome12 v1.1 and UM HUNT Biobank v1.0). Samples that failed to reach a 99% call rate, had contamination>2.5% as estimated with BAF Regress[14], large chromosomal copy number variants, lower call rate of a technical duplicate pair and twins, gonosomal constellations other than XX and XY, or whose inferred sex contradicted the reported gender, were excluded. Samples that passed quality control were analysed in a second round of genotype calling following the Genome Studio quality control protocol described elsewhere[15]. Genomic position, strand orientation and the reference allele of genotyped variants were determined by aligning their probe sequences against the human genome (Genome Reference Consortium Human genome build 37 and revised Cambridge Reference Sequence of the human mitochondrial DNA; http://genome.ucsc.edu) using BLAT[16]. PLINK v1.90[17] was then used to exclude variants if their probe sequences could not be perfectly mapped, cluster separation was<0.3, Gentrain score<0.15, showed deviations from Hardy Weinberg equilibrium in unrelated samples of European ancestry with p-value<0.0001), had a call rate<99%, or another assay with higher call rate genotyped the same variant. Ancestry of all samples was inferred by projecting all genotyped samples into the space of the principal components of the Human Genome Diversity Project (HGDP) reference panel (938 unrelated individuals; downloaded from http://csg.sph.umich.edu/chaolong/LASER/)[18;19] using PLINK. Recent European ancestry was defined as samples that fell into an ellipsoid spanning exclusively European populations of the HGDP panel. The different arrays were harmonized by reducing to a set of overlapping variants and excluding variants that showed frequency differences>15% between data sets, or that were monomorphic in one and had MAF>1% in another data set. The resulting genotype data were phased using Eagle2 v2.3 47. Imputation was performed on the 69,716 samples of recent European ancestry using Minimac3 (v2.0.1, http://genome.sph.umich.edu/wiki/Minimac3)[4] with default settings (2.5 Mb reference based chunking with 500kb windows) and a customized Haplotype Reference consortium release 1.1 (HRC v1.1) for autosomal variants and HRC v1.1 for chromosome X variants[20]. The customized reference panel represented the merged panel of two reciprocally imputed reference panels: (a) 2201 low-coverage whole-genome sequences samples from the HUNT study and (b) HRC v1.1 with 1023 HUNT WGS samples removed before merging. We excluded imputed variants with Rsq<0.3 or minor allele count<1, resulting in 24.2 million well-imputed variants. After restricting to those with available phenotype information 66476 individuals (284 cases and 66192 controls) were included in the analysis. Association analyses were conducted using SAIGE[21], a generalized mixed effects model approach, to account for cryptic population structure and relatedness when modelling the association between genotype probabilities (dosages) and OCD. Models were adjusted for sex, birth year, genotyping batch and four principal components (PCs). PCs were computed using PLINK.

DRAFT

*MoBa | Ask, H. | Norway | 27063603*

The *Norwegian Mother, Father and Child Cohort Study* (MoBa) is a population based pregnancy cohort study conducted by the Norwegian Institute of Public Health[22]. The establishment of MoBa and initial data collection was based on a license from the Norwegian Data Protection Agency and approval from The Regional Committees for Medical and Health Research Ethics. The MoBa cohort is now based on regulations related to the Norwegian Health Registry Act. The current study was approved by The Regional Committees for Medical and Health Research Ethics. Participants were recruited from all over Norway between 1999 and 2008. The women consented to participation in 41% of the pregnancies. The cohort now includes 114,500 children, 95,200 mothers and 75,200 fathers. Blood samples were obtained from the mothers and fathers at 17–18 weeks of gestation and from mothers and children (umbilical cord) at birth[23]. Genotyping of the entire MoBa cohort is ongoing. For the current study we used the available genotype data from 17,000 randomly selectred trios, genotyped in three batches on three different arrays (harvest12: Illumina HumanCoreExome12v1.1, harvest24: Illumina HumanCoreExome24v1.0, rotterdam1: Illumina Global Screening Array MD v.1.0.). harvest12 and harvest24 were genotyped at the Genomics core facility in Trondheim, Norway while the rotterdam1 samples were genotyped at ERASMUS MC, Rotterdam, Netherlands. PLINK version 1.90 beta 3.36[5] was used to conduct quality control, details of QC have been previously described by Helgeland et al (2019)[24]. Individuals were excluded if they had a genotyping call rate below 95% or autosomal heterozygosity greater than four standard deviations from the sample mean. SNPs were excluded if they were ambiguous (A/T and C/G), had a genotyping call rate below 98%, minor allele frequency of less than 1%, or Hardy-Weinberg equilibrium P-value less than $1 \times 10^{-6}$. Population stratification was assessed using the HapMap phase 3 release 3 as a reference, by principal component analysis using EIGENSTRAT version 6.1.4. Visual inspection identified a homogenous population of European ethnicity and individuals of non-European ethnicity were removed. Phasing was conducted using Shapeit 2 release 837 and the duoHMM approach was used to account for the pedigree structure. Imputation was conducted using the Haplotype reference consortium (HRC) release 1-1 as the genetic reference panel. The Sanger Imputation Server was used to perform the imputation with the Positional Burrows-Wheeler Transform (PBWT). The phasing and imputation were conducted separately for each genotyping batch. A core homogeneous sample of European ethnicity across all batches and arrays were available for use in analysis (totals prior to analysis-specific exclusions for relatedness: N mothers = 14,804; N fathers =15,198). OCD diagnosis was ascertained through linkage to the Norwegian Patient Registry (ICD-10 codes from specialist health care registered from 2008-2018). Case inclusion criteria were ICD-10 code F42 diagnosed at least once. Control inclusion criteria were no F-diagnosis. Control individuals were excluded if they were related to any of the cases (pihat>0.2). Sex ratio in the cases was 40:60 (male:female) and in the controls 50:50. GWA analysis was based on 104 cases and 2193 controls. GWAS was performed using SAIGE, including the first 10 PCs and genotyping batch as covariates.

*OCD-WWF | Domschke, K; Berberich G. | Germany*

For the *Obsessive compulsive disorder - Windach Würzburg Freiburg* (OCD-WWF) study, 129 inpatients with OCD (age [$mean \pm SD$]: 34.47±11.86 years; 66 female) were recruited at the Psychosomatic Hospital Windach, Windach, Germany, between 2014 and 2017. OCD diagnosis was ascertained on the basis of a structured clinical interview according to DSM-IV criteria (SCID-I) by experienced psychiatrists and/or clinical psychologists. Inclusion criteria were age at inclusion between 18 and 80 years and Caucasian descent (self-report up to third generation). Exclusion criteria comprised severe somatic and neurological disorders, the consumption of illegal drugs, and pregnancy. Comorbid tic disorder, trichotillomania, skin-picking disorder or other current axis I diagnoses except for depression ($N$ = 71), specific phobias ($N$ = 10), generalized anxiety disorder ($N$ = 1), social phobia ($N$ = 5), panic disorder ($N$ = 2), agoraphobia ($N$ = 5) or post-traumatic stress disorder ($N$ = 3) were excluded. The study was approved by the ethics committee of the University of Würzburg, Germany and was conducted according to the ethical principles of the Helsinki Declaration. All patients gave written informed consent prior to participation. Cases and controls were genotyped on Illumina's Global screening array. GWAS analysis was performed using ricopili, employing standard parameters. First, we ran an automated round of pre-imputation QC. The pre-imputation QC step involved a series of hard filters on variant and sample level data, including removing variants with pre-sample pruning call rate $< 0.95$, samples with call rate $< 0.98$, FHET outside of $\pm$ 0.20, samples with discrepancies between reported and derived sex, and post sample-pruning variants that meet any of the following : 1) call rate $< 0.98$, 2) missing difference $>$ 0.02, 3) invariant positions, 4) MAF $> 0.01$, 5) HWE p $< 1E^{-6}$ in controls, and 6) HWE p $< 1E^{-10}$ in cases. Ricopili's *impute_dirsub* module was used to conduct imputation using the Haplotype Reference Consortium (HRC) reference panel. Ricopili's *impute_dirsub* module was used to conduct imputation using the 1000's genomes (1000G) reference panel. We conducted PCA on these samples across high-confidence imputed genotypes using the *pacer_sub* ricopili module and tested the first 20 PCs for significant association with sample case/control status. In a final step, we used ricopili's *postimp_navi* module to conduct the GWAS analysis.

*EGOS | Grice, D. E. | Sweden | 31907560*

The EGOS source population consists of individuals born in Sweden between January 1954 and December 1998 that presented at least two diagnoses of OCD or chronic tic disorders (CTD) at different time points in the Swedish National Patient Register (NPR), and were followed between January 1997 and December 2012 (N = 20,374). The International Classification of Disease (ICD) was used for the

DRAFT

identification of OCD cases (F49 ICD version 10). Detailed information for each individual was obtained through linkage to the Swedish national registers, e.g., family relatedness, identification of additional psychiatric diagnoses, medical diagnoses, birth-related variables, and relevant demographic and social data. To create an epidemiologically valid subset of the source cohort that also includes biospecimens and additional phenotyping, individuals were contacted from within the source population. Study participants could elect whether to donate blood or saliva. Individuals are aged 16-64 years. All samples were genotyped using the Global Screening Array (GSA). Further description of the EGOS cohort has been represented by Mahjani et al.[25]. The dataset that was used in this meta-analysis consists of 1026 OCD cases from EGOS and 1208 controls from LifeGene. GWAS of the EGOS cohort was performed together with the NORDiC-SWE cohort (see description of the NORDiC-SWE cohort for analysis details). EGOS was supported by a grant from the Beatrice and Samual A. Seaver Foundation to DEG. Ethical approvals were obtained from the Institutional Review Board (IRB) at the Icahn School of Medicine at Mount Sinai, New York, NY, and the Regional Ethical Review Board in Stockholm.

*NORDiC-SWE | Crowley, J; Mataix-Cols, D.; Rück, C. | Sweden | 31424634*

A paper describing the rationale, design and methods of the NORDiC study has been published previously[26]. NORDiC-SWE is the Swedish case-control arm of the study and all samples were collected in Sweden between 2015 and 2019. This study has been approved by a local ethics board (Stockholm Regional EPN) and all subjects provided informed consent. OCD Cases have a primary ICD-10 and/or DSM-5 diagnosis of OCD from a multidisciplinary specialist OCD team (established with a semi-structured instrument such as the MINI or the SCID). All patients are included in the study regardless of psychiatric comorbidity, as long as they fulfill strict diagnostic criteria for OCD. Patients are excluded in cases of diagnostic uncertainty, such as OCD secondary to a neurological disorder or CNS insult, or where the differential diagnosis between OCD and an alternative condition is unclear. Our cases had a mean age at symptom debut of 12.1 years and 59% were female. Approximately 58% of patients had a documented psychiatric comorbidity. Controls were unrelated to any OCD case to the third degree and unaffected with OCD. Controls were excluded if they had a lifetime history of anorexia nervosa (controls were inherited from an anorexia GWAS). The controls included 95% females. The NORDiC-SWE cohort consists of 971 OCD cases and 2735 controls (LIFEGENE control batch 1 $N$ = 1026) and LIFEGENE control batch 2 ($N$ = 1389)). Subjects provided either blood or saliva for DNA extraction. All samples were genotyped on the Illumina Global Screening Array at LIFE&BRAIN in Bonn, Germany. We formed a case/control dataset by merging PLINK filesets for five different cohorts (EGOS and NORDiC-SWE case-only, LIFEGENE 1, LIFEGENE 2 and EGOS control-only) using PLINK v1.90b3n. Using PLINK v1.90b3n[5], samples were filtered for duplicates (pi_hat $\geq$ 0.95) and cryptic relatedness (pi_hat$\geq$ 0.2). We selected individuals of European ancestry using principal component analysis implemented in PEDDY v0.4.3 .We defined variants as QC-failing if they met one of the following criteria: 1) maximum genotype missingness in a cohort $>$ 0.02; 2) allele frequency $<$ 0.001 in at least one cohort; 3) max – min allele frequency $>$ 0.1 across all five cohorts; 4) max – min allele frequency $>$ 0.03 across all three control cohorts; 5) genomewide significant in a control vs. control synthetic GWAS. A total of 154791 variants that met at least one of these criteria were excluded. We used the ricopili v2018_Dec_7.001 pipeline to run an automated round of pre-imputation QC. The pre-imputation QC step involved a series of hard filters on variant and sample level data, including removing variants with pre-sample pruning call rate $<$ 0.95, samples with call rate $<$ 0.98, FHET outside of $\pm$ 0.20, samples with discrepancies between reported and derived sex, and post sample-pruning variants that meet any of the following : 1) call rate $<$ 0.98, 2) missing difference $>$ 0.02, 3) invariant positions, 4) MAF $>$ 0.01, 5) HWE p $<$ $1E^{-6}$ in controls, and 6) HWE p $<$ $1E^{-10}$ in cases. The final dataset consisted of 647,335 variant calls across a total of 1997 cases and 3943 controls. Ricopili's *impute_dirsub* module was used to conduct imputation using the Haplotype Reference Consortium (HRC) reference panel. In our imputation run we used eagle v2.3.5 for pre-phasing, and minimac3 v2.0.1 for imputation. We derived 3 different imputed callsets from this process: 1) a set of high confidence imputed genotypes (2771425 SNPs), 2) 7112906 imputed best-guess genotypes with medium level accuracy, and 3) genotypes for variants where imputation accuracy is lowered in order to increase the total number of variants included in the imputation (resulting in 8995398 SNPs). We elected to run our GWAS across the largest dataset of imputed variants that were generated during the imputation process on a subset of samples that were of European ancestry. We conducted PCA on these samples across high-confidence imputed genotypes using the *pacer_sub* ricopili module and tested the first 20 PCs for significant association with sample case/control status (significant = p-value $<$ 0.05/20). We identified PCs 1, 3 and 14 as significant predictors and used them as covariates in the GWAS analysis. We used ricopili's *postimp_navi* module to conduct the final GWAS. Summary statistics were well controlled across the separate GWAS. We noted a lambda of 1.01 and a lambda1000 of 1.01 across a total of 7679714 tested SNPs. The NORDiC-SWE study was approved by the Regional Ethics Committee, Stockholm (EPN Stockholm) and the Institutional Review Board (IRB) at the University of North Carolina at Chapel Hill.

*NORDiC-NOR | Crowley, J.; Kvale, G.; Hansen, S. | Norway | 31424634*

NORDiC-NOR is the Norwegian case-control arm of the NORDiC study and all samples were collected in Norway between 2016 and 2019. This study has been approved by a local ethics board (REK West) and all subjects provided informed consent. OCD Cases have a primary ICD-10 and/or

DRAFT

DSM-5 diagnosis of OCD from a multidisciplinary specialist OCD team (established with a semi-structured instrument such as the MINI or the SCID). All patients are included in the study regardless of psychiatric comorbidity, as long as they fulfill strict diagnostic criteria for OCD. Patients are excluded in cases of diagnostic uncertainty, such as OCD secondary to a neurological disorder or CNS insult, or where the differential diagnosis between OCD and an alternative condition is unclear. Our cases had a mean age at symptom debut of 17 years and 65% were female. Approximately 51% of patients had a documented psychiatric comorbidity. Controls were unrelated to any OCD case to the third degree and unaffected with OCD. Among the controls, 50% were female. Subjects provided either blood or saliva for DNA extraction. All samples were genotyped on the Illumina Global Screening Array at LIFE&BRAIN in Bonn, Germany. The pre-GWAS QC applied to the NORDiC-NOR dataset (482 cases, 343 controls in the raw data) was nearly identical to that applied to NORDIC-SWE data after the merging of separate genotype data, consisting of pruning of cryptic relatedness, marking of samples that are of likely European ancestry and pruning of the dataset for single variants where there was suggestive evidence of technical biases or batch effects. In our cryptic relatedness QC step we identified 4 samples that had a mean pi_hat with other samples $\geq 0.1$, 74 samples that had evidence of being a sample duplicate (pi_hat $\geq 0.95$) and 8 samples from the remaining cohort with evidence of cryptic relatedness (pi_hat $\geq 0.2$). We found a total of 6263 variants overlapped between the merged PLINK fileset and the 1000 genomes data included in PEDDY, and identified a total of 368 cases and 315 controls with likely European ancestry. We performed variant-level QC on a PC-pruned subset of 340 cases and 307 controls, defining variants as failing if they met one of the following criteria: 1) maximum genotype missingness in a cohort $> 0.02$; 2) allele frequency of 0 in at least one cohort; 3) max – min allele frequency $> 0.1$. A total of 136,525 variants that met at least one of these criteria were excluded, leaving us with a final case/control dataset consisting of genotype calls across 479,358 variants. We used calls across these variants in samples that had been pruned for relatedness issues (not including standard pairwise relatedness issues as ricopili can detect these) as input for the GWAS (407 cases, 340 controls). Imputation and GWAS was performed analogously to the NORDiC-SWE and EGOS analysis. The final data set consisted of 365 cases and 315 controls. Imputed callsets resulted in 1) 3043464 high confidence imputed genotypes 2) 7277174 genotypes with medium level, and 3) 8964589 genotypes with a low imputation accuracy. We identified PCs 1, 2, 3 and 4 as significant predictors and used them as covariates in the NORDIC-NOR GWAS. The analysis resulted in a lambda of 1.00 and a lambda1000 of 1.01 across a total of 7518582 tested SNPs. The NORDIC-NOR study was approved by the Norwegian Regional Committee for Medical and Health Research Ethics (REC-West) under project number 2018/52 REKVest (PI Bjarne Hansen) and project number: 2014/75 REKVest (PI: Jan Haavik).

*OCGAS-nestadt | Nestadt G., OCGAS Consortium | USA*

The OCGAS study was conducted at one of the five participating recruitment sites of the National Institute of Mental Health. The study was approved by the IRB boards at: Johns Hopkins University School of Medicine, Brown Medical School, New York State Psychiatric Institute and College of Physicians and Surgeons at Columbia University, University of California Los Angeles (UCLA) School of Medicine, Massachusetts General Hospital and Harvard Medical School, National Institute of Mental Health, and Keck School of Medicine at the University of Southern California. Samples were collected between 2007 and 2014. The sample comprised of trios (including an affected proband and both parents) or in some cases a proband and an unaffected sibling. Each case was evaluated by a MD- or PhD-level clinical psychologist using the Structured Clinical Interview for DSM-IV (SCID). The checklist of obsessions and compulsions from the Y-BOCS, refined to include the age of onset, offset, and severity of each symptom, as well as the Y-BOCS scores for the worst episode (lifetime) was recorded. Course and treatment response variables were also included. A similar model was used for evaluating tics and Tourette disorder. Axis I disorder diagnoses were assigned using the JHU Diagnostic Assignment Checklist, an instrument that documents the criteria for over 20 DSM-IV disorders; this instrument also was the primary tool for the diagnostic consensus procedure. The SCID-II was used to evaluate four personality disorders (schizotypic, obsessive-compulsive, avoidant, and dependent), and the FISC was used to obtain additional information about each participant from a knowledgeable informant. Children over the age of eight were assessed in the same way, except that the Kiddie-SADS was used in place of the SCID. Final diagnostic status was assigned based on the consensus of two psychiatrists or psychologists reviewing the case independently. The agreement between diagnosticians using the Diagnostic Assignment Checklist has been studied and found to be excellent for variables such as age at onset of OCD. The chance-corrected percent agreement between the diagnosticians for the diagnosis of OCD was $K = 0.92$; for age at onset of OCD, $K = 0.88$ (for age +/- 5 years), and Pearson's $r = 0.71$. The diagnostic information from each site was reviewed by one of the five members of the JHU diagnostic consensus committee to ensure comparability across sites. For study inclusion, probands were required to meet DSM-IV criteria for OCD - with onset of obsessions and/or compulsions before the age of 18 years (*mean* = 9.4 years; *SD* = 6.35). Subjects with an age-of-onset >17 years, schizophrenia, severe mental retardation that does not permit an evaluation to characterize the psychiatric disorder, Tourette disorder or OCD occurring exclusively in the context of depression (secondary OCD) were excluded. In addition, individuals were removed from the sample if they were previously diagnosed with brain pathology including brain tumors, Huntington's disease, Parkinson's disease, or Alzheimer's disease. This resulted in a final sample of 212 cases and 212 controls. Samples were genotyped on the

DRAFT

Ilumina PsychChip array at USC. Ethics approvals for the OCGAS study were obtained from the Hopkins Medicine Institutional Review Boards.

The OCD Collaborative Genetics Association Study (OCGAS) is a collaborative research study and was funded by the following NIMH Grant Numbers: MH071507, MH079489, MH079487, MH079488 and MH079494.

*OCGAS-ab | Arnold P., Burton C. | Canada | 27777633, 31772171*

Samples in the OCGAS-ab cohort were collected between 2008-2015 and consist of trios. The study was approved by REB at Hospital for Sick Children. Cases were required to meet DSM-IV criteria for OCD and were selected based on the K-SADS and CY-BOCS questionnaires. Exclusion criteria for cases were age of onset <18 years of age, psychosis, and history of severe neurological disorders other than Tourette's disorder. Of the 55 cases and 55 controls, 50% are female, the mean age was 16.38 ($SD$ = 3.78). One individual presented a co-morbid diagnosis of ASD, 11 of ADHD, six of Tics/Tourette's syndrome, five of anxiety disorders, 1 of eating disorder, 2 of depression, and two presented learning difficulties. Samples were genotyped on the Ilumina PsychChip array at USC.Ethics approvals for the OCGAS study were obtained from the Hopkins Medicine Institutional Review Boards.

*OCGAS-gh |Grünblatt E., Walitza S. | Switzerland and Germany | 28065182, 29102815*

Participants in the early-onset OCD cohort were recruited at the Departments of Child and Adolescent Psychiatry of the Universities of Würzburg, Marburg, Aachen, and Freiburg in Germany and Zurich in Switzerland. Informed written consent was obtained in all cases by the participants or their parents. The study was approved by the ethical commissions of all involved universities in accordance with the latest version of the Declaration of Helsinki, including an ethical permission granted by the Ethic Committees from Aachen, Würzburg, Marburg, Freiburg and the Cantonal Ethic Commission of Zürich (Ref. Nr. 39/97, 140/3 and EK: KEK-ZH-Nr. 2010-0340/3). Samples were collected between 2000 and 2016 and resulted in 56 cases and 56 controls in trios or case-control samples. Patients were included if they fulfilled the diagnostic criteria for current OCD according to DSM-4 and ICD-10. To assess OCD diagnostic criteria, early-onset OCD patients and parents were interviewed separately by senior clinicians with a semi-structured diagnostic interview of psychiatric disorders in children and adolescents (Kinder-DIPS; children and parents version)[27]; the patients and parents located in Zürich underwent the German version of a semi-structured clinical interview (K-SADS-PL)[28]. In addition, severity and additional characteristics of OCD symptoms were evaluated with the Children's Yale-Brown Obsessive Compulsive Scale (CY-BOCS)[29]. Kinder-DIPS or K-SADS-PL was used to screen for the existence of comorbid disorders (affective-, anxiety-, eating- and tic disorders, attention-deficit/hyperactivity disorder, conduct-, oppositional disorder, as well as substance use, abuse, psychosis and somatic diseases) in children and adolescents. CASCAP-D was used to screen for autistic spectrum disorders[30]. Present and lifetime Tourette´s syndrome and tic disorders were assessed with the adapted German version[31] of the Child and Adult Schedule for Tourette and Other Behavioral Syndromes (STOBS)[32] and the Yale Global Tic Severity Scale (YGTSS)[33] in the Zürich patients. Case exclusion criteria were a lifetime history of Tourette's syndrome, psychotic disorder, autism spectrum disorder, mentar retardation (IQ < 70) or alcohol dependence. Patients with co-morbid disorders were only included if OCD was the primary diagnosis. Controls were excluded if they presented a major psychiatric disorder or an IQ < 70. All pediatric OCD patients received cognitive behavior therapy, while when insufficient, drug treatment was added, most commonly with an SSRI. Samples were genotyped on the ILMN PsychChip array and genotyped at USC in one batch.

*BioVU | Davis, L. K. | Nashville, Tennessee, USA*

Vanderbilt University Medical Center (VUMC) is a tertiary care center that provides inpatient and outpatient care in Nashville, TN. The VUMC electronic health record (EHR) system was established in 1990 and includes data on billing codes from the International Classification of Diseases, 9th and 10th editions (ICD-9 and ICD-10), Current Procedural Terminology (CPT) codes, laboratory values, reports, and clinical documentation. In 2007, VUMC launched a biobank, BioVU, which links a patient's DNA sample to their EHR. The BioVU Consent form is provided to patients in the outpatient clinic environments at VUMC. The form states policies on data sharing and privacy, and should a signature be obtained, makes any blood leftover from clinical care eligible for BioVU banking[34]. The Vanderbilt University Medical Center Institutional Review Board oversees BioVU and approved this project (IRB201609). OCD case status was determined through a combination of ICD codes, medications, and natural language processing for EHR notes. Using ICD codes, cases were defined as individuals with any codes for OCD (ICD9: 300.3, ICD10: F42, F63.3, F45.22). Additional cases were gathered by first finding individuals with "obsessive-compulsive" in clinic notes, problem lists, discharge summaries or clinical communications. Next, these individuals were required to have OCD medication or cognitive behavioral therapy in their EHR. Finally, individuals with metabolic disorder codes (ICD9: 277.89, 277.99, ICD10: E88.9, E88.89) were excluded. Controls were defined as any individual without OCD codes, metabolism disorder codes, "obsessive- compulsive" in their clinical notes, and evidence of OCD medication. The initial sample included 1062 cases (median age in years across EHR: 32.75, 58.6% female) and 40316 controls (median age in years across EHR: 53, 57% female). We obtained genotype information on 94,474 BioVU individuals genotyped on the Illumina MEGA EX array. Using PLINK v1.9[5], genotypes were filtered for SNP and individual call rates, sex discrepancies, and excessive heterozygosity. We selected individuals of European ancestry

DRAFT

using principal component analysis implemented in FlashPCA[35] and confirmed the absence of genotyping batch effects through logistic regression with *batch* as the phenotype. Autosomes were imputed to the HRC panel using Michigan Imputation Server[4] in five batches. After imputation, genotypes were converted to hard calls with PLINK using the default threshold settings. SNPs with multiple alleles or imputation quality less than $R^2$ of 0.3 were excluded. Next, SNPs with minor allele frequency less than 0.005 or genotyping rates less than 0.98 were excluded. Individuals with call rates less than 0.98 were excluded. We ran a series of principal component analyses (PCA) to determine BioVU individuals of European genetic ancestry. First, we performed PCA using FlashPCA on BioVU combined with CEU, YRI, and CHB reference sets from 1000 Genomes Project Phase 3[36]. Principal components were scaled so that the axes could be interpreted as proportions of genetic ancestry. We selected BioVU individuals who were within 40% of the CEU cluster along the CEU-CHB axis and within 30% of the CEU cluster on the CEU-YRI axis, generating a once-PCA filtered European set. To ensure subsequent steps would remove SNPs associated with reduced quality rather than cryptic population substructure, we filtered the previously identified BioVU European cluster to identify individuals falling within the CEU, TSI, and GIH 1000 genomes populations, producing a twice-filtered European set. Using the twice-filtered European set we conducted a series of SNP checks. First, we filtered individuals with IBS greater than 0.2 and calculated principal components to use as covariates. Next, we checked for imputation batch effects by conducting pairwise logistic regression of the five imputation batches using sex and top 10 principal components as covariates. SNPs with p-values less than 0.001 in the additive model were flagged. We then compared MAF between BioVU and the CEU reference population. Any SNPs with a MAF difference greater than 0.1 were flagged. SNPs with a Hardy-Weinberg Equilibrium p-value less than 10E-10 were flagged. Finally, the flagged SNPs from the batch effect, MAF difference, and HWE were excluded from the once-PCA filtered BioVU European set, resulting in 9,386,383 SNPs for analysis. The final dataset comprised of 1041 cases and 38613 controls. To account for the large case-control imbalance, we used SAIGE for the GWAS. Covariates included were sex, median age across medical record, and top 10 prinicpal components.

*UKBB | Breen, G. | United Kingdom*

The UK Biobank sample consists of 776 OCD cases and 125729 controls, see Bycroft et al.[37] for a general description of the UK Biobank resource. The here included data has not been published previously. The data was derived from a online mental health questionnaire, completed by participants between July 2016 and July 2017. Research on the UK Biobank is conducted under a generic Research Tissue Bank approval from the UK North West Multi-centre Research Ethics Committee (MREC). This research was approved to be conducted under that approval by the governing Research Ethics Committee of the UK Biobank. The analyses in this paper were performed under an approved extension to project 16577. Cases were defined by self-report of a professional diagnosis of OCD ("Have you been diagnosed with [Obsessive compulsive disorder (OCD)] by a professional, even if you don't have it currently?"). The median age of cases at the time of report (not age at diagnosis) was 62 (inter quantile range (*IQR* = 55-68), 59% of cases are female. All participants who did not report a OCD diagnosis were included as controls. The median age of controls at the time of report was 65 (*IQR* = 58-70), 56% of controls are female. Genotyping of cases and controls was performed on the Affymetrix Axiom UK Biobank Array / Affymetrix Axiom UK BiLEVE array[37] and genotyped in the Affymetrix Research Services Laboratory, Santa Clara, CA in several batches. For genotype quality control the following filters were applied to the data: MAF$>$0.01 and call-rate$>$98%. 4-means clustering was applied on the first two principle components to determine participants of European ancestries (as described by Warren et al.[38]). Further, quality assurance outliers marked by UKBB were removed. Relatives (KING relatedness$>$0.044) greedily (e.g. keeping the parents in a parent-child trio) and participants with mismatched sex were removed (reported females with FX $\geq$ 0.6, reported males with FX $\leq$ 0.85; FX determined using chrX SNPs in approximate linkage equilibrium, r2$<$0.2). Imputation was performed with IMPUTE4 using a combined HRC/UK-10k reference panel. Imputed data was filtered on MAF $\geq$ 0.01 and INFO $\geq$ 0.4. GWAS was performed using SAIGE, including the first six principle components from participants of European ancestries, factors for genotyping batch and assessment centre as covariates.

*EstBB, | Metspalu, A. | Estonia*

The Estonian Biobank (EstBB) is a population-based cohort with a rich variety of phenotypic and health-related information collected for each participant[39]. At recruitment, participants have signed a consent to allow follow-up linkage of their electronic health records (EHR), thereby providing a longitudinal collection of phenotypic information. The health data of the participants is continuously updated through periodical linking with records from the national Health Insurance Fund Treatment Bills (from 2004 onwards), Tartu University Hospital (from 2008), and North Estonia Medical Center (from 2005). For every participant there is information on diagnoses in ICD-10 coding and drug dispensing data, including drug ATC codes, prescription status and purchase date (if available). Individual level data analysis in the EstBB was carried out under ethical approval from the Estonian Committee on Bioethics and Human Research (Estonian Ministry of Social Affairs) and data release N05 from the EstBB.

The samples from the Estonian Biobank were genotyped at the Genotyping Core Facility of the Institute of Genomics, University of Tartu using the Global Screening Array (GSAv1.0, GSAv2.0, and GSAv2.0_EST) from Illumina. Altogether 155,772 samples were genotyped and PLINK format files[17] exported using GenomeStudio v2.0.4. Individuals were excluded from the analysis if their call-rate was $<$ 95% or if the sex defined based on heterozygosity of

DRAFT

the X chromosome did not match the sex in the phenotype data. Variants were excluded if the call-rate was $< 95\%$ and HWE p-value $< 1E-4$ (autosomal variants only). Variant positions were updated to genome build 37 and all alleles were switched to the TOP strand using tools and reference files provided at https://www.well.ox.ac.uk/ wrayner/strand/. After QC the dataset contained 154,201 samples for imputation. Before imputation variants with MAF $< 1\%$ and indels were removed. Prephasing was done using the Eagle v2.3 software[40]. The number of conditioning haplotypes Eagle2 uses when phasing each sample was set to: –Kpbwt=20000. Imputation was done using Beagle v.28Sep18.793[41] with effective population size ne=20,000. An Estonian population specific imputation reference of 2297 WGS samples was used[42].

For the genome-wide study of OCD in EstBB cases were defined as participants with F42* ICD10 diagnosis codes in their EHRs. We conducted a GWAS on 138,508 individuals of European ancestry, including 493 cases and 138,015 controls. The analysis was performed with the SAIGE software, including related individuals and adjusting for the first 10 principal components (PCs) of the genotype matrix, as well as for birth year, birth year squared and sex.

#### Supplementary Tables

Supplementary Tables S1-S10 can be found in the following online document (click here).

DRAFT

#### Supplementary Figures

**Fig. S1. QQ (quantile-quantile) plot** for the 14140 cases and 562117 controls in the OCD meta-analysis. Association quantiles of the -log(10) p-values, received from inverse variant weighted meta-analysis, are plotted against the quantiles expected under the null hypothesis. The shading indicates 95% confidence interval. Lambda, the genomic inflation factor, is the observed median $\chi^2$ test statistic under the null. Lambda1000 indicates the lambda if the sample contained 500 cases and 500 controls.

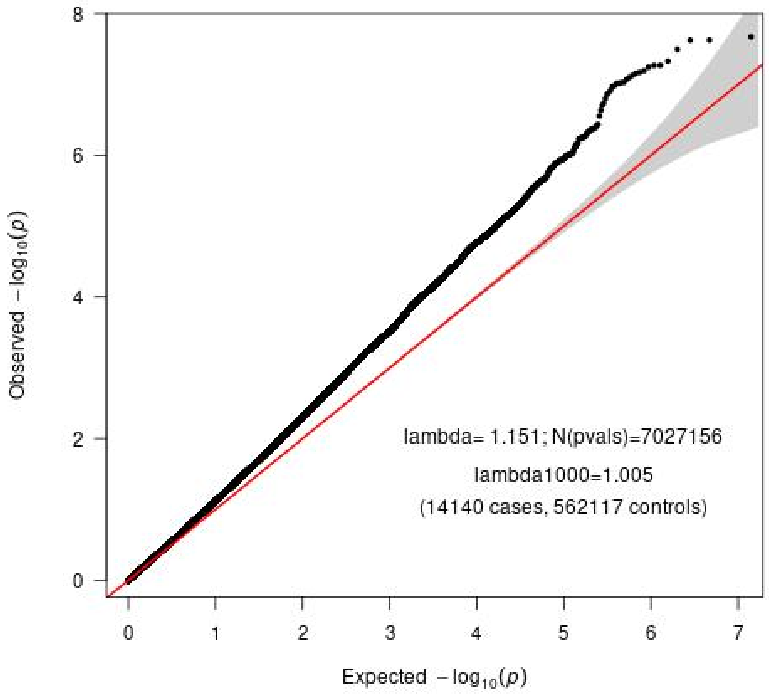


DRAFT

**Fig. S2. Regional plot of association signal near rs13262595** The –log10(P) of SNPs in the OCD meta-analysis GWAS is shown on the left y axis. The recombination rates expressed in centimorgans (cM) per Mb (Megabase) (blue line) are shown on the right y axis. Position in Mb is on the x axis. Only the SNPs with association p-value less than 0.1 were plotted. The most associated SNP is shown as a purple diamond.

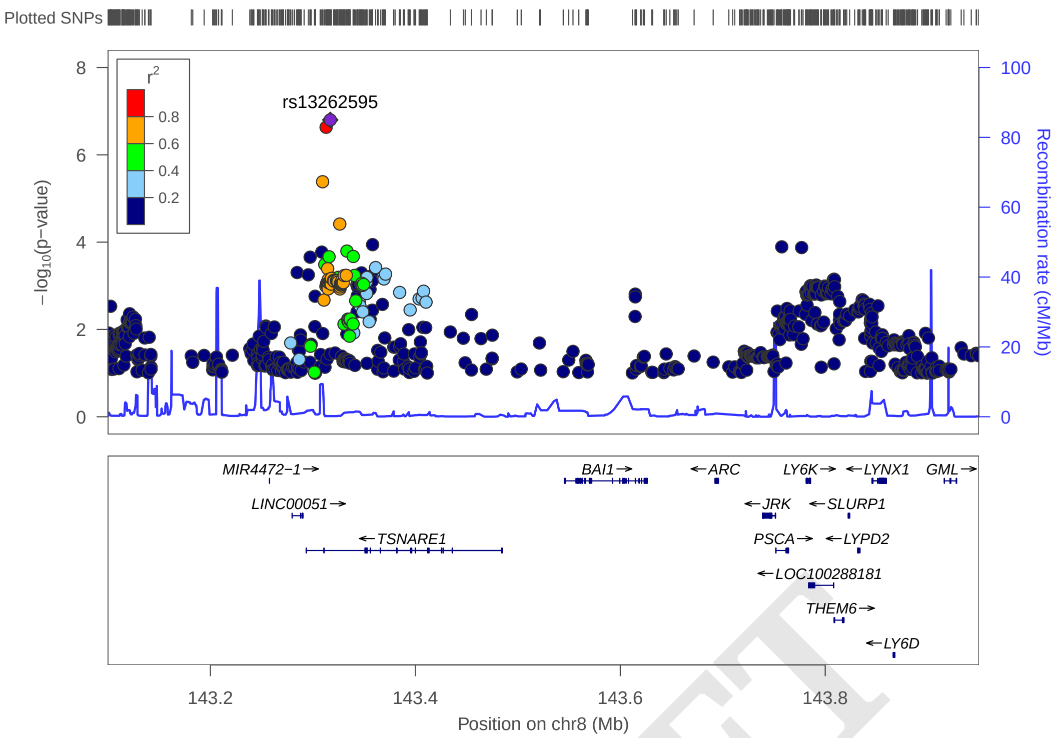


**Fig. S3. Regional plot of association signal near rs674094** The –log10(P) of SNPs in the OCD meta-analysis GWAS is shown on the left y axis. The recombination rates expressed in centimorgans (cM) per Mb (Megabase) (blue line) are shown on the right y axis. Position in Mb is on the x axis. Only the SNPs with association p-value less than 0.1 were plotted. The most associated SNP is shown as a purple diamond.

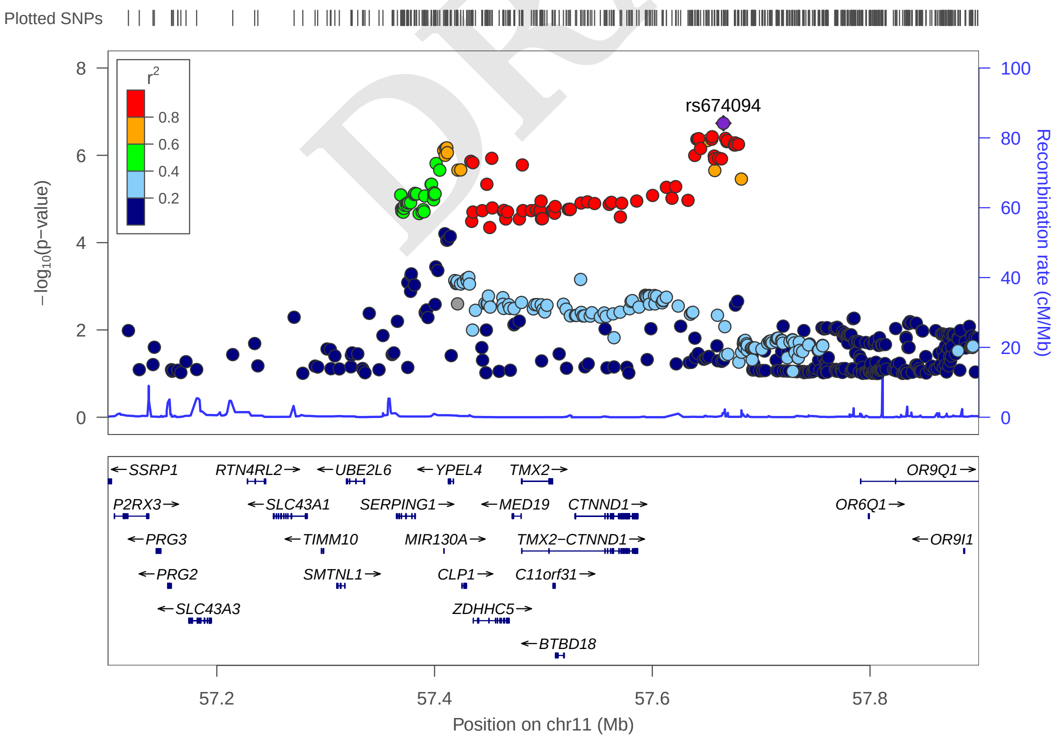


DRAFT

**Fig. S4. Regional plot of association signal near rs79712033** The –log10(P) of SNPs in the OCD meta-analysis GWAS is shown on the left y axis. The recombination rates expressed in centimorgans (cM) per Mb (Megabase) (blue line) are shown on the right y axis. Position in Mb is on the x axis. Only the SNPs with association p-value less than 0.1 were plotted. The most associated SNP is shown as a purple diamond.

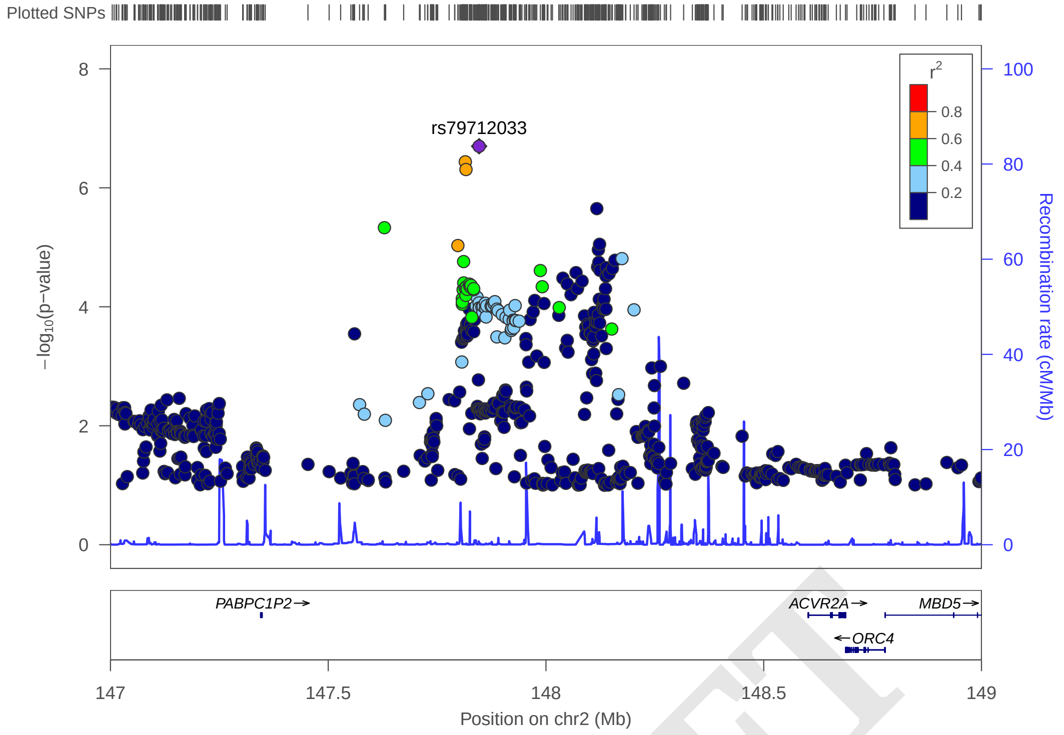


**Fig. S5. Regional plot of association signal near rs7219489** The –log10(P) of SNPs in the OCD meta-analysis GWAS is shown on the left y axis. The recombination rates expressed in centimorgans (cM) per Mb (Megabase) (blue line) are shown on the right y axis. Position in Mb is on the x axis. Only the SNPs with association p-value less than 0.1 were plotted. The most associated SNP is shown as a purple diamond.

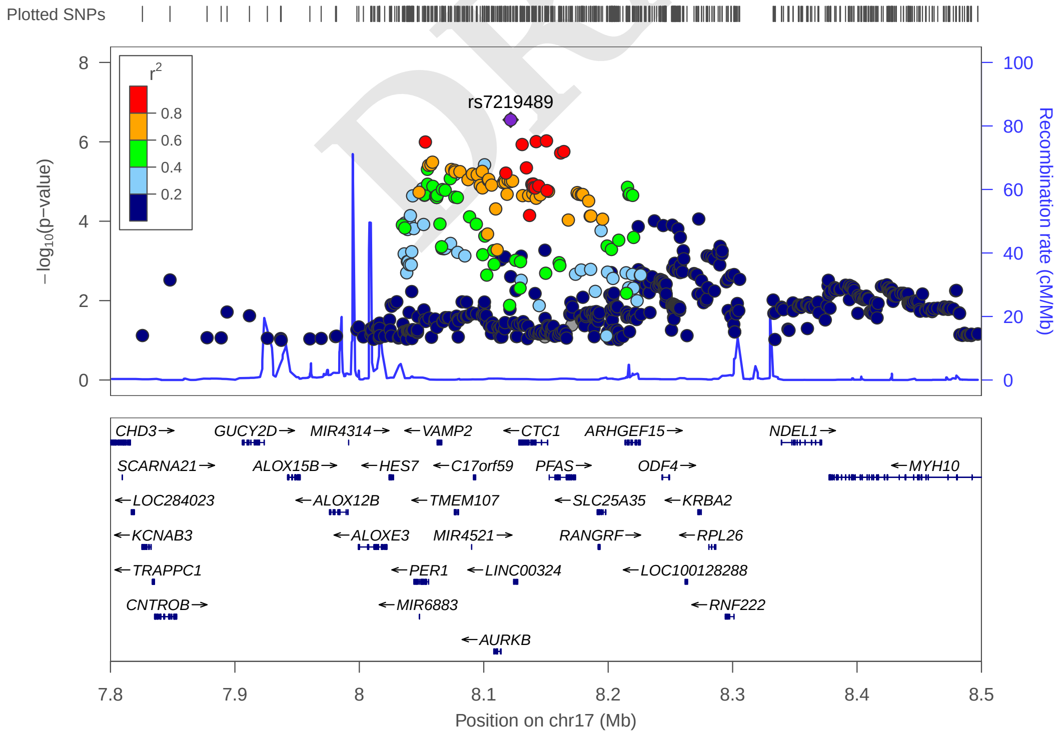


DRAFT

**Fig. S6. Regional plot of association signal near rs9535127** The –log10(P) of SNPs in the OCD meta-analysis GWAS is shown on the left y axis. The recombination rates expressed in centimorgans (cM) per Mb (Megabase) (blue line) are shown on the right y axis. Position in Mb is on the x axis. Only the SNPs with association p-value less than 0.1 were plotted. The most associated SNP is shown as a purple diamond.

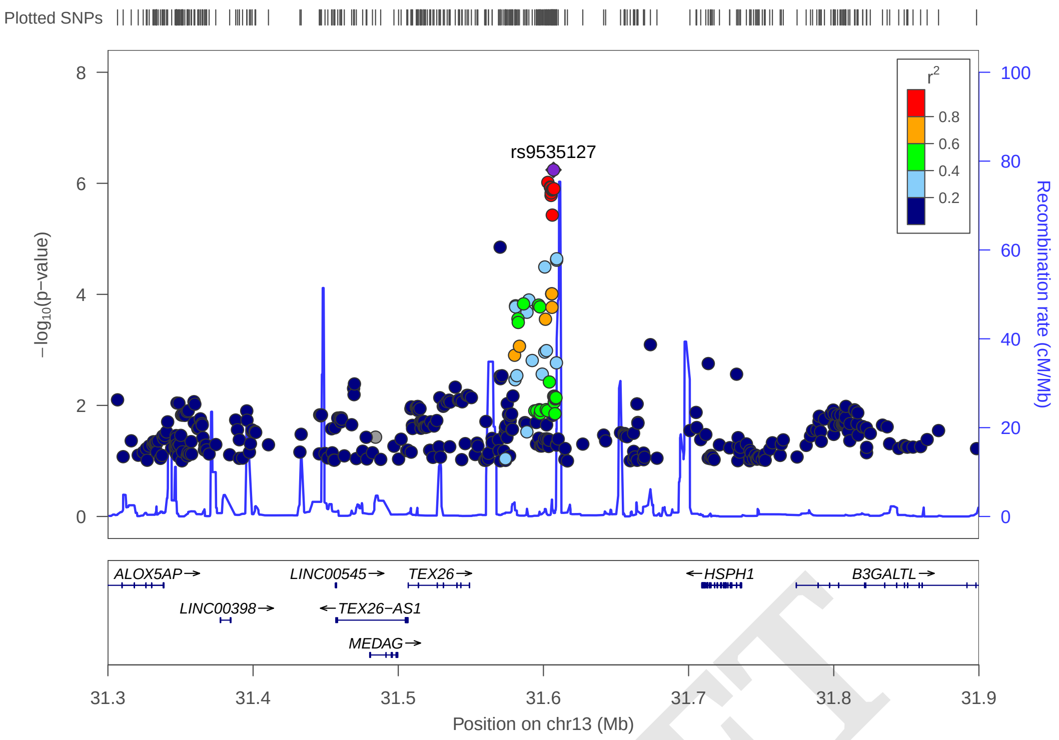


DRAFT

**Fig. S7. Regional plot of association signal near rs7128224** The –log10(P) of SNPs in the OCD meta-analysis GWAS is shown on the left y axis. The recombination rates expressed in centimorgans (cM) per Mb (Megabase) (blue line) are shown on the right y axis. Position in Mb is on the x axis. Only the SNPs with association p-value less than 0.1 were plotted. The most associated SNP is shown as a purple diamond.

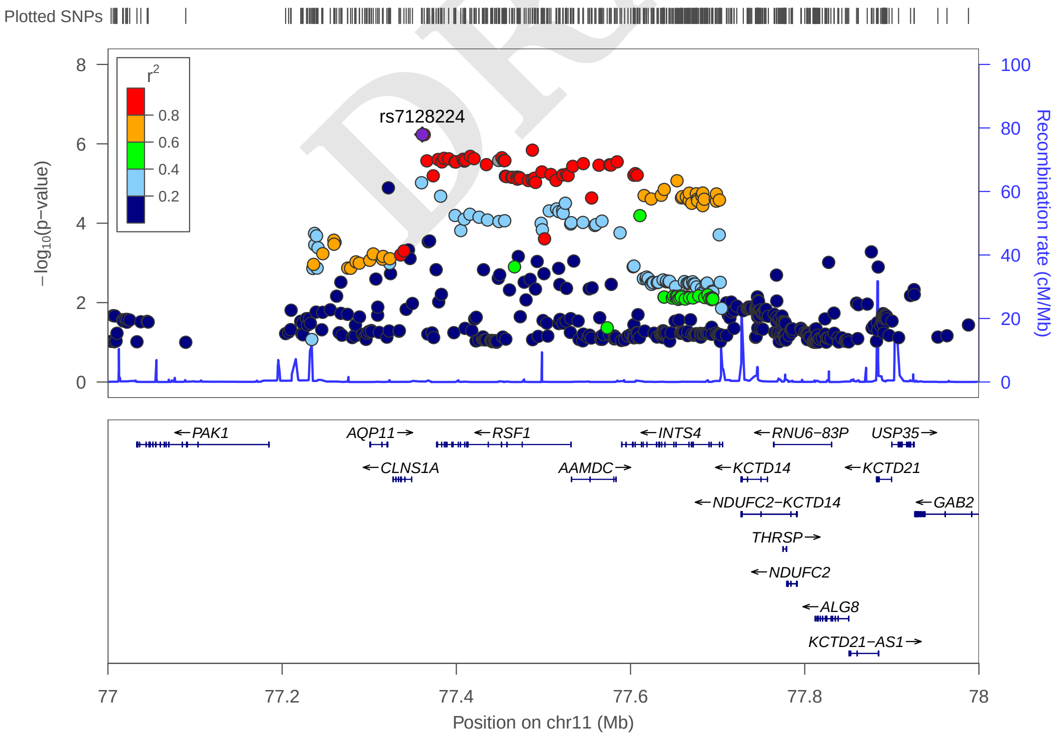

**Fig. S8. Regional plot of association signal near rs424541** The –log10(P) of SNPs in the OCD meta-analysis GWAS is shown on the left y axis. The recombination rates expressed in centimorgans (cM) per Mb (Megabase) (blue line) are shown on the right y axis. Position in Mb is on the x axis. Only the SNPs with association p-value less than 0.1 were plotted. The most associated SNP is shown as a purple diamond.

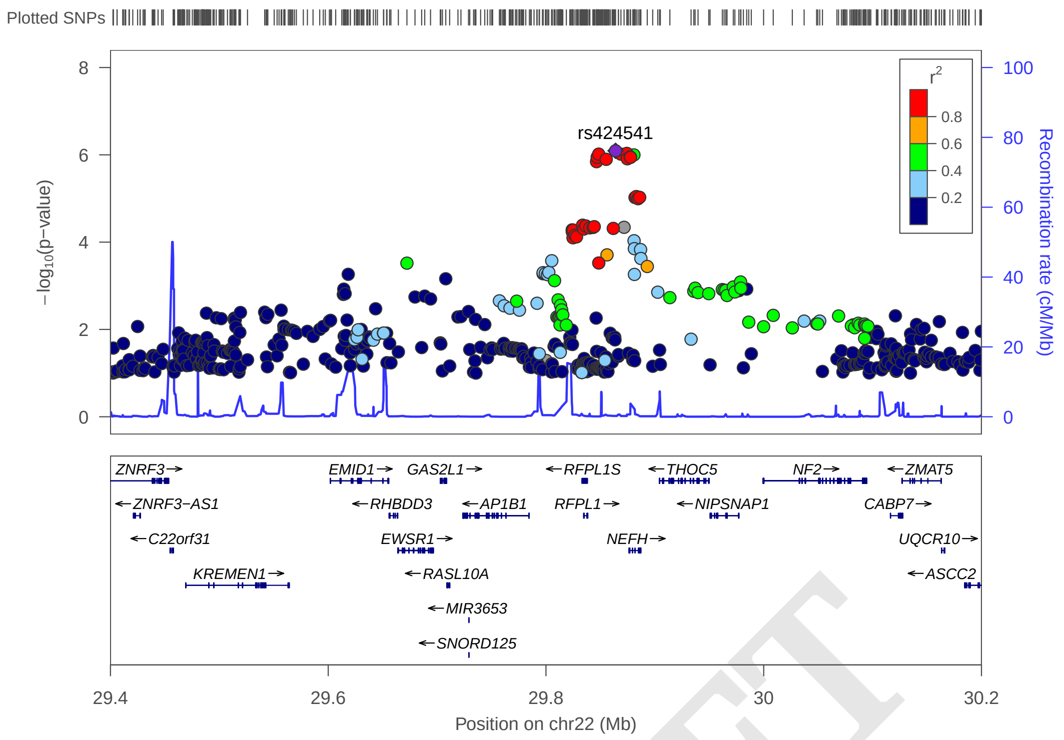


DRAFT

**Fig. S9. Regional plot of association signal near rs34289388** The –log10(P) of SNPs in the OCD meta-analysis GWAS is shown on the left y axis. The recombination rates expressed in centimorgans (cM) per Mb (Megabase) (blue line) are shown on the right y axis. Position in Mb is on the x axis. Only the SNPs with association p-value less than 0.1 were plotted. The most associated SNP is shown as a purple diamond

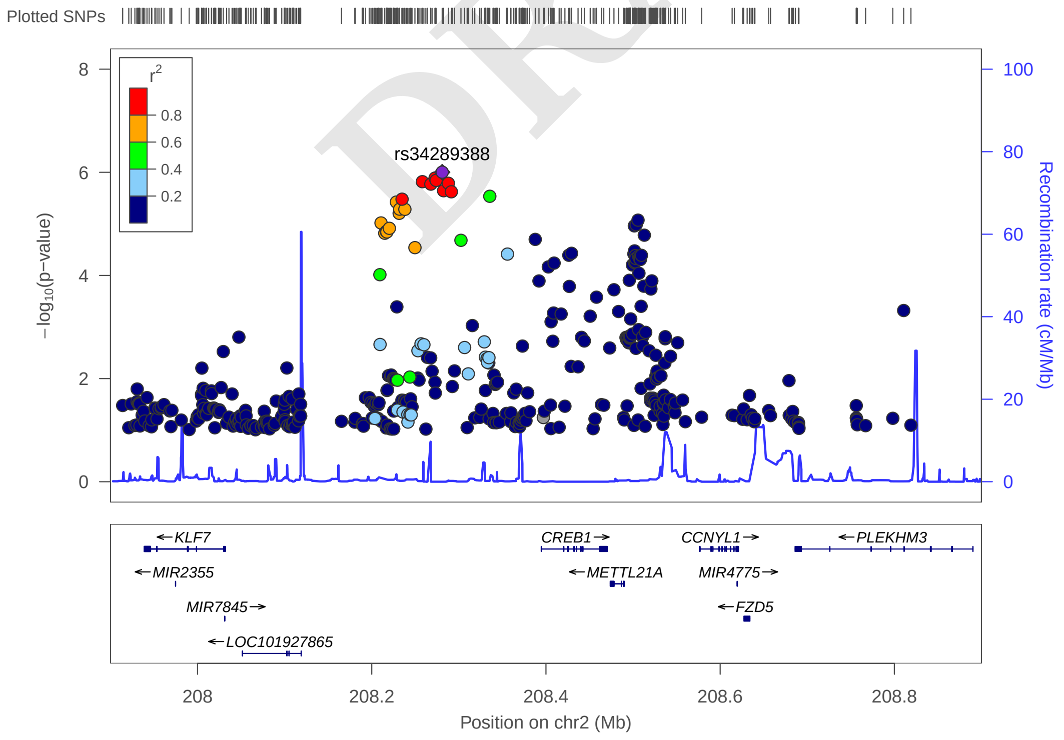

**Fig. S10. Forest plot of SNP rs13262595.** The plot shows the effect estimate with 95%-confidence interval for each cohort contributing to the meta-analysis and for the inverse variance weighted meta analysis. The table lists INFO (imputation score), p-value, f_ca(n) (frequency cases), f_co(n) (frequency controls), ln(OR), and STDerr (standard error) for each of the contributing cohorts and for the meta-analysis. At the top, + indicates a positive direction of effect, - a negative direction of effect while ? indicates that the SNP was not contained in the respective cohort.

| **rs13262595** | | **A/G** | | **8:143316970** | | | |
|---|---|---|---|---|---|---|---|
| | | +++++−+−+−++−++−+++ | | het_P: | 0.261 | het_I: | 15.8 |
| | **ngt** | **info** | **p_value** | **f_ca(n)** | **f_co(n)** | **ln(OR)** | **STDerr** |
| dan_daner_610_final. | 0 | 1.04 | 0.0626 | 0.480(323) | 0.427(323) | 0.205 | 0.11 |
| dan_daner_AUS_refstrict_final. | 0 | 0.99 | 0.338 | 0.445(757) | 0.433(5368) | 0.0538 | 0.0561 |
| dan_daner_DOTS_final. | 0 | 0.98 | 0.0935 | 0.451(2678) | 0.439(10410) | 0.0524 | 0.0312 |
| dan_daner_EPOC_final. | 0 | 0.99 | 0.858 | 0.423(195) | 0.424(204) | 0.0253 | 0.142 |
| dan_daner_EstBB_final. | 0 | 0.99 | 0.728 | 0.411(493) | 0.416(138015) | −0.0227 | 0.0653 |
| dan_daner_FinnGen_final. | 0 | 1.00 | 0.0794 | 0.426(790) | 0.401(161390) | 0.0934 | 0.0532 |
| dan_daner_HUNT_final. | 0 | 0.99 | 0.929 | 0.455(284) | 0.460(66192) | −0.0075 | 0.0847 |
| dan_daner_MoBa_final. | 0 | 0.98 | 0.265 | 0.399(104) | 0.437(2193) | −0.164 | 0.147 |
| dan_daner_Nordic_norway_HRC_final. | 0 | 0.97 | 0.96 | 0.444(365) | 0.432(315) | 0.00608 | 0.123 |
| dan_daner_Nordic_sweden_HRC_final. | 0 | 1.00 | 0.0106 | 0.455(1997) | 0.424(3943) | 0.103 | 0.0401 |
| dan_daner_UKBB_saige_final. | 1 | 1.00 | 0.554 | 0.441(776) | 0.434(125729) | 0.0305 | 0.0516 |
| dan_daner_bioVU_saige_strict_final. | 0 | 1.00 | 0.478 | 0.457(1014) | 0.448(38613) | 0.0324 | 0.0457 |
| dan_daner_iocd_final. | 0 | 1.01 | 0.000164 | 0.466(1519) | 0.424(3541) | 0.169 | 0.0448 |
| dan_daner_ocgas−ab_final. | 0 | 0.90 | 0.618 | 0.419(55) | 0.466(55) | −0.172 | 0.345 |
| dan_daner_ocgas−gh_final. | 0 | 1.17 | 0.6 | 0.499(56) | 0.446(56) | 0.135 | 0.257 |
| dan_daner_ocgas−nestadt_final. | 0 | 1.04 | 0.307 | 0.448(212) | 0.478(212) | −0.14 | 0.137 |
| dan_daner_ocgas_all2_final. | 0 | 1.03 | 0.00314 | 0.482(986) | 0.434(1023) | 0.185 | 0.0628 |
| dan_daner_psych_broad_final. | 0 | 0.98 | 0.0247 | 0.477(1396) | 0.458(4009) | 0.102 | 0.0456 |
| dan_daner_wue_final. | 0 | 0.94 | 0.625 | 0.454(140) | 0.440(526) | 0.0688 | 0.141 |
| **dan_** | **1** | **0.99** | **1.58e−07** | **0.454(14140)** | **0.424(562117)** | **0.073** | **0.0139** |

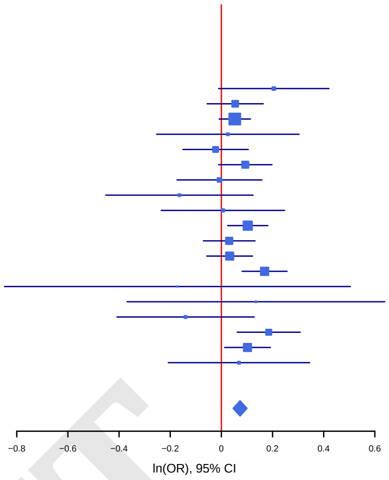


DRAFT

**Fig. S11. Forest plot of SNP rs34289388.** The plot shows the effect estimate with 95%-confidence interval for each cohort contributing to the meta-analysis and for the inverse variance weighted meta analysis. The table lists INFO (imputation score), p-value, f_ca(n) (frequency cases), f_co(n) (frequency controls), ln(OR), and STDerr (standard error) for each of the contributing cohorts and for the meta-analysis. At the top, + indicates a positive direction of effect, - a negative direction of effect while ? indicates that the SNP was not contained in the respective cohort.

| **rs34289388** | | **G/A** | | **2:208280861** | | | |
|---|---|---|---|---|---|---|---|
| | | +−−−−−−+−−+−−−−−−−− | | het_P: | 0.175 | het_I: | 23.2 |
| | **ngt** | **info** | **p_value** | **f_ca(n)** | **f_co(n)** | **ln(OR)** | **STDerr** |
| IOCDF-GC TR | 0 | 1.15 | 0.0461 | 0.784(323) | 0.831(323) | −0.265 | 0.133 |
| AUS | 0 | 0.97 | 0.127 | 0.818(757) | 0.801(5368) | 0.11 | 0.0723 |
| DOTS | 0 | 1.00 | 0.00249 | 0.797(2678) | 0.778(10410) | 0.115 | 0.038 |
| EPOC | 0 | 0.97 | 0.899 | 0.799(195) | 0.796(204) | 0.022 | 0.174 |
| Estonia | 0 | 1.00 | 0.318 | 0.817(493) | 0.804(138015) | 0.0807 | 0.0809 |
| FinnGen | 0 | 1.00 | 0.132 | 0.792(790) | 0.773(161390) | 0.0941 | 0.0625 |
| HUNT | 0 | 1.00 | 0.293 | 0.743(284) | 0.761(66192) | −0.104 | 0.0986 |
| MoBa | 0 | 1.00 | 0.181 | 0.817(104) | 0.775(2193) | 0.227 | 0.17 |
| NORDiC-nor | 0 | 1.00 | 0.464 | 0.766(365) | 0.787(315) | −0.106 | 0.145 |
| NORDiC-swe_EGOS | 0 | 1.01 | 0.26 | 0.783(1997) | 0.772(3943) | 0.0538 | 0.0478 |
| UKBB | 1 | 1.00 | 0.147 | 0.817(776) | 0.801(125729) | 0.093 | 0.0642 |
| bioVU | 0 | 0.99 | 0.488 | 0.802(1014) | 0.796(38613) | 0.0394 | 0.0569 |
| IOCDF-GC CC | 0 | 1.00 | 0.0125 | 0.805(1519) | 0.789(3541) | 0.14 | 0.0562 |
| ocgas-ab | 0 | 1.04 | 0.526 | 0.742(55) | 0.704(55) | 0.219 | 0.345 |
| ocgas-gh | 0 | 1.01 | 0.0622 | 0.828(56) | 0.715(56) | 0.63 | 0.338 |
| ocgas-nes | 0 | 0.99 | 0.0551 | 0.837(212) | 0.788(212) | 0.35 | 0.183 |
| OCGAS | 0 | 0.99 | 0.0301 | 0.823(986) | 0.796(1023) | 0.177 | 0.0815 |
| Psych_Broad | 0 | 0.97 | 0.392 | 0.804(1396) | 0.794(4009) | 0.0489 | 0.0571 |
| WWF | 0 | 1.00 | 0.144 | 0.804(140) | 0.763(526) | 0.246 | 0.169 |
| **Meta - analysis** | **1** | **1.00** | **9.93e−07** | **0.800(14140)** | **0.788(562117)** | **0.0838** | **0.0171** |

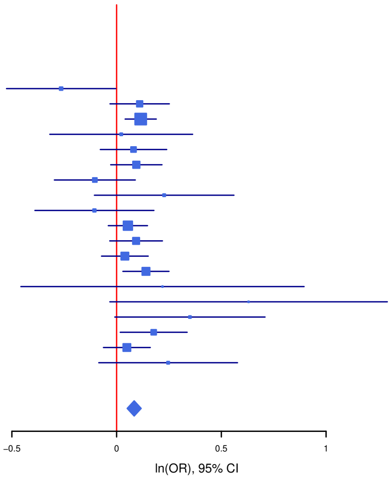

**Fig. S12. Forest plot of SNP rs424541.** The plot shows the effect estimate with 95%-confidence interval for each cohort contributing to the meta-analysis and for the inverse variance weighted meta analysis. The table lists INFO (imputation score), p-value, f_ca(n) (frequency cases), f_co(n) (frequency controls), ln(OR), and STDerr (standard error) for each of the contributing cohorts and for the meta-analysis. At the top, + indicates a positive direction of effect, - a negative direction of effect while ? indicates that the SNP was not contained in the respective cohort.

| **rs424541** | | **C/T** | | **22:29863922** | | | |
|---|---|---|---|---|---|---|---|
| | | ++++−++−++++++++++− | | het_P: | 0.596 | het_I: | 0.0 |
| | **ngt** | **info** | **p_value** | **f_ca(n)** | **f_co(n)** | **ln(OR)** | **STDerr** |
| IOCDF-GC TR | 0 | 0.98 | 0.982 | 0.833(323) | 0.833(323) | −0.00351 | 0.15 |
| AUS | 0 | 1.00 | 0.0836 | 0.820(757) | 0.835(5368) | −0.125 | 0.072 |
| DOTS | 0 | 0.99 | 0.0729 | 0.827(2678) | 0.838(10410) | −0.0736 | 0.0411 |
| EPOC | 0 | 0.98 | 0.736 | 0.857(195) | 0.837(204) | 0.0666 | 0.198 |
| Estonia | 0 | 1.00 | 0.0412 | 0.741(493) | 0.769(138015) | −0.155 | 0.0761 |
| FinnGen | 0 | 1.00 | 0.315 | 0.731(790) | 0.746(161390) | −0.0599 | 0.0595 |
| HUNT | 0 | 1.00 | 0.836 | 0.798(284) | 0.791(66192) | 0.0214 | 0.104 |
| MoBa | 0 | 0.99 | 0.674 | 0.788(104) | 0.801(2193) | −0.0749 | 0.178 |
| NORDiC-nor | 0 | 0.95 | 0.594 | 0.817(365) | 0.806(315) | −0.0832 | 0.156 |
| NORDiC-swe_EGOS | 0 | 1.01 | 0.00032 | 0.791(1997) | 0.817(3943) | −0.179 | 0.0498 |
| UKBB | 1 | 1.00 | 0.858 | 0.835(776) | 0.837(125729) | −0.0124 | 0.0693 |
| bioVU | 0 | 0.99 | 0.00177 | 0.818(1014) | 0.843(38613) | −0.196 | 0.0628 |
| IOCDF-GC CC | 0 | 1.01 | 0.481 | 0.843(1519) | 0.843(3541) | −0.0432 | 0.0613 |
| ocgas-ab | 0 | 1.07 | 0.196 | 0.801(55) | 0.881(55) | −0.579 | 0.448 |
| ocgas-gh | 0 | 0.93 | 0.304 | 0.786(56) | 0.836(56) | −0.373 | 0.363 |
| ocgas-nes | 0 | 0.96 | 0.831 | 0.842(212) | 0.840(212) | −0.0423 | 0.198 |
| OCGAS | 0 | 0.99 | 0.306 | 0.847(986) | 0.856(1023) | −0.0926 | 0.0903 |
| Psych_Broad | 0 | 1.00 | 0.571 | 0.852(1396) | 0.855(4009) | −0.0361 | 0.0638 |
| WWF | 0 | 0.91 | 0.345 | 0.852(140) | 0.831(526) | 0.186 | 0.197 |
| **Meta - analysis** | **1** | **1.00** | **8.09e−07** | **0.818(14140)** | **0.789(562117)** | **−0.0895** | **0.0181** |

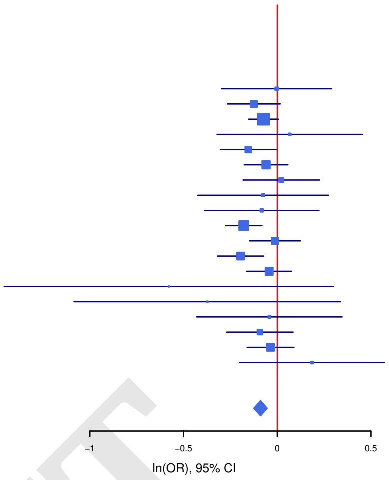


DRAFT

**Fig. S13. Forest plot of SNP rs674094.** The plot shows the effect estimate with 95%-confidence interval for each cohort contributing to the meta-analysis and for the inverse variance weighted meta analysis. The table lists INFO (imputation score), p-value, f_ca(n) (frequency cases), f_co(n) (frequency controls), ln(OR), and STDerr (standard error) for each of the contributing cohorts and for the meta-analysis. At the top, + indicates a positive direction of effect, - a negative direction of effect while ? indicates that the SNP was not contained in the respective cohort.

| **rs674094** | | **A/C** | | **11:57665336** | | | |
|---|---|---|---|---|---|---|---|
| | | ++?+++++++++?+??+++ | | het_P: | 0.574 | het_I: | 0.0 |
| | **ngt** | **info** | **p_value** | **f_ca(n)** | **f_co(n)** | **ln(OR)** | **STDerr** |
| IOCDF-GC TR | 0 | 0.95 | 0.079 | 0.324(323) | 0.280(323) | 0.22 | 0.125 |
| AUS | 0 | 0.94 | 0.333 | 0.319(757) | 0.308(5368) | 0.0594 | 0.0614 |
| DOTS | 0 | 0.99 | 0.000244 | 0.327(2678) | 0.301(10410) | 0.122 | 0.0331 |
| EPOC | 0 | 0.86 | 0.0383 | 0.319(195) | 0.249(204) | 0.345 | 0.168 |
| Estonia | 0 | 0.99 | 0.856 | 0.357(493) | 0.354(138015) | 0.0122 | 0.0675 |
| FinnGen | 0 | 1.00 | 0.517 | 0.362(790) | 0.357(161390) | 0.0357 | 0.0551 |
| HUNT | 0 | 0.99 | 0.192 | 0.340(284) | 0.312(66192) | 0.12 | 0.0917 |
| MoBa | 0 | 0.97 | 0.838 | 0.327(104) | 0.321(2193) | 0.0308 | 0.151 |
| NORDiC-nor | 0 | 1.00 | 0.808 | 0.318(365) | 0.291(315) | 0.0323 | 0.133 |
| NORDiC-swe_EGOS | 0 | 0.96 | 0.0993 | 0.318(1997) | 0.308(3943) | 0.0722 | 0.0438 |
| UKBB | 1 | 0.99 | 0.0322 | 0.321(776) | 0.296(125729) | 0.12 | 0.0561 |
| IOCDF-GC CC | 0 | 1.01 | 0.124 | 0.296(1519) | 0.288(3541) | 0.0754 | 0.049 |
| OCGAS | 0 | 1.05 | 0.108 | 0.301(986) | 0.276(1023) | 0.11 | 0.0686 |
| Psych_Broad | 0 | 1.00 | 0.959 | 0.298(1396) | 0.304(4009) | 0.0025 | 0.0492 |
| WWF | 0 | 0.91 | 0.067 | 0.351(140) | 0.299(526) | 0.276 | 0.151 |
| **Meta - analysis** | **1** | **0.99** | **1.84e−07** | **0.319(14140)** | **0.332(562117)** | **0.0827** | **0.0159** |

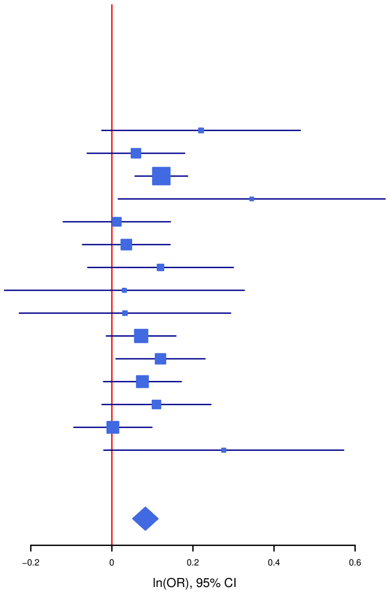

**Fig. S14. Forest plot of SNP rs7128224.** The plot shows the effect estimate with 95%-confidence interval for each cohort contributing to the meta-analysis and for the inverse variance weighted meta analysis. The table lists INFO (imputation score), p-value, f_ca(n) (frequency cases), f_co(n) (frequency controls), ln(OR), and STDerr (standard error) for each of the contributing cohorts and for the meta-analysis. At the top, + indicates a positive direction of effect, - a negative direction of effect while ? indicates that the SNP was not contained in the respective cohort.

| **rs7128224** | | **T/G** | | **11:77360928** | | | |
|---|---|---|---|---|---|---|---|
| | | ++?+−−++++++?+??+++ | | het_P: | 0.21 | het_I: | 21.9 |
| | **ngt** | **info** | **p_value** | **f_ca(n)** | **f_co(n)** | **ln(OR)** | **STDerr** |
| IOCDF-GC TR | 0 | 0.98 | 0.713 | 0.324(323) | 0.315(323) | 0.0443 | 0.121 |
| AUS | 0 | 0.96 | 0.433 | 0.325(757) | 0.312(5368) | 0.0473 | 0.0602 |
| DOTS | 0 | 0.96 | 0.0856 | 0.297(2678) | 0.285(10410) | 0.0592 | 0.0344 |
| EPOC | 0 | 0.90 | 0.781 | 0.304(195) | 0.310(204) | −0.0464 | 0.167 |
| Estonia | 0 | 0.99 | 0.575 | 0.360(493) | 0.369(138015) | −0.0375 | 0.0669 |
| FinnGen | 0 | 1.00 | 0.413 | 0.268(790) | 0.265(161390) | 0.0488 | 0.0596 |
| HUNT | 0 | 0.97 | 0.00547 | 0.361(284) | 0.308(66192) | 0.258 | 0.0927 |
| MoBa | 0 | 0.97 | 0.352 | 0.337(104) | 0.301(2193) | 0.143 | 0.153 |
| NORDiC-nor | 0 | 0.95 | 0.00708 | 0.340(365) | 0.282(315) | 0.363 | 0.135 |
| NORDiC-swe_EGOS | 0 | 0.99 | 0.198 | 0.318(1997) | 0.302(3943) | 0.0558 | 0.0433 |
| UKBB | 1 | 0.99 | 0.417 | 0.311(776) | 0.301(125729) | 0.0454 | 0.056 |
| IOCDF-GC CC | 0 | 0.99 | 0.0953 | 0.313(1519) | 0.296(3541) | 0.0815 | 0.0488 |
| OCGAS | 0 | 0.98 | 0.0263 | 0.319(986) | 0.286(1023) | 0.155 | 0.0697 |
| Psych_Broad | 0 | 0.96 | 0.000828 | 0.337(1396) | 0.301(4009) | 0.163 | 0.0488 |
| WWF | 0 | 0.90 | 0.298 | 0.338(140) | 0.303(526) | 0.158 | 0.152 |
| **Meta - analysis** | **1** | **0.99** | **5.82e−07** | **0.316(14140)** | **0.308(562117)** | **0.0802** | **0.016** |

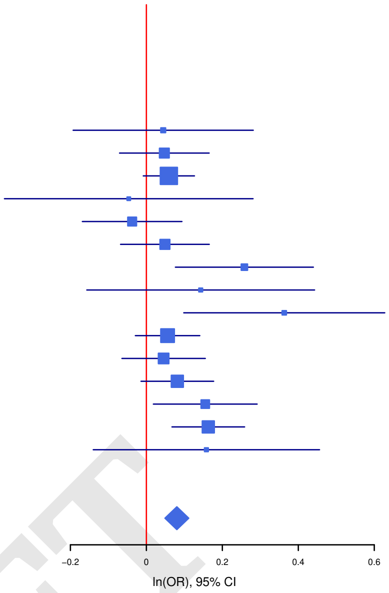


**Fig. S15. Forest plot of SNP rs7219489.** The plot shows the effect estimate with 95%-confidence interval for each cohort contributing to the meta-analysis and for the inverse variance weighted meta analysis. The table lists INFO (imputation score), p-value, f_ca(n) (frequency cases), f_co(n) (frequency controls), ln(OR), and STDerr (standard error) for each of the contributing cohorts and for the meta-analysis. At the top, + indicates a positive direction of effect, - a negative direction of effect while ? indicates that the SNP was not contained in the respective cohort.

| **rs7219489** | | **G/T** | | **17:8121622** | | | |
|---|---|---|---|---|---|---|---|
| | | −−−−−+−−−−−+−−?−??−−+ | | het_P: | 0.605 | het_I: | 0.0 |
| | **ngt** | **info** | **p_value** | **f_ca(n)** | **f_co(n)** | **ln(OR)** | **STDerr** |
| IOCDF-GC TR | 0 | 1.03 | 0.3 | 0.705(323) | 0.678(323) | 0.123 | 0.119 |
| AUS | 0 | 0.97 | 0.0647 | 0.750(757) | 0.729(5368) | 0.119 | 0.0647 |
| DOTS | 0 | 0.98 | 0.091 | 0.730(2678) | 0.718(10410) | 0.0589 | 0.0349 |
| EPOC | 0 | 0.89 | 0.555 | 0.677(195) | 0.703(204) | −0.0979 | 0.166 |
| Estonia | 0 | 0.99 | 0.121 | 0.742(493) | 0.719(138015) | 0.111 | 0.0716 |
| FinnGen | 0 | 1.00 | 0.0688 | 0.769(790) | 0.744(161390) | 0.109 | 0.06 |
| HUNT | 0 | 1.00 | 0.847 | 0.732(284) | 0.725(66192) | 0.0183 | 0.0947 |
| MoBa | 0 | 1.00 | 0.987 | 0.716(104) | 0.716(2193) | −0.00258 | 0.162 |
| NORDiC-nor | 0 | 0.95 | 0.0412 | 0.743(365) | 0.697(315) | 0.281 | 0.138 |
| NORDiC-swe_EGOS | 0 | 0.95 | 7.31e−05 | 0.751(1997) | 0.717(3943) | 0.184 | 0.0465 |
| UKBB | 1 | 1.00 | 0.0662 | 0.739(776) | 0.718(125729) | 0.105 | 0.0572 |
| bioVU | 0 | 0.99 | 0.376 | 0.729(1014) | 0.720(38613) | 0.045 | 0.0508 |
| IOCDF-GC CC | 0 | 0.98 | 0.266 | 0.721(1519) | 0.713(3541) | 0.0558 | 0.0502 |
| OCGAS | 0 | 1.00 | 0.571 | 0.712(986) | 0.704(1023) | 0.0395 | 0.0698 |
| Psych_Broad | 0 | 1.01 | 0.58 | 0.714(1396) | 0.708(4009) | 0.0272 | 0.0492 |
| WWF | 0 | 0.89 | 0.981 | 0.710(140) | 0.709(526) | −0.00371 | 0.158 |
| **Meta - analysis** | **1** | **0.99** | **2.79e−07** | **0.732(14140)** | **0.727(562117)** | **0.0807** | **0.0157** |

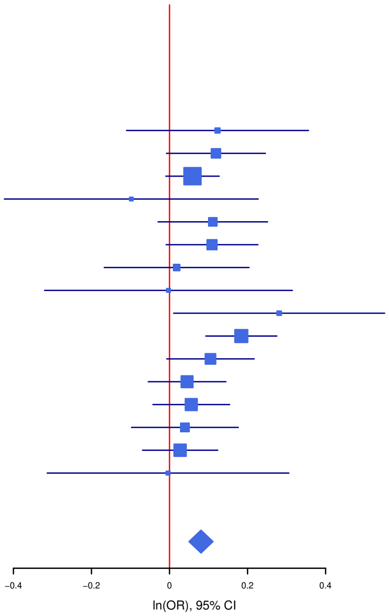

**Fig. S16. Forest plot of SNP rs79712033.** The plot shows the effect estimate with 95%-confidence interval for each cohort contributing to the meta-analysis and for the inverse variance weighted meta analysis. The table lists INFO (imputation score), p-value, f_ca(n) (frequency cases), f_co(n) (frequency controls), ln(OR), and STDerr (standard error) for each of the contributing cohorts and for the meta-analysis. At the top, + indicates a positive direction of effect, - a negative direction of effect while ? indicates that the SNP was not contained in the respective cohort.

| **rs79712033** | | **A/G** | | **2:147846501** | | | |
|---|---|---|---|---|---|---|---|
| | | ++++?+?–+––+?+??+++ | | het_P: | 0.105 | het_I: | 33.7 |
| | **ngt** | **info** | **p_value** | **f_ca(n)** | **f_co(n)** | **ln(OR)** | **STDerr** |
| IOCDF-GC TR | 0 | 0.93 | 0.302 | 0.949(323) | 0.936(323) | 0.257 | 0.249 |
| AUS | 0 | 0.88 | 0.24 | 0.948(757) | 0.941(5368) | 0.156 | 0.133 |
| DOTS | 0 | 0.95 | 0.00328 | 0.945(2678) | 0.934(10410) | 0.2 | 0.0679 |
| Estonia | 0 | 0.97 | 0.0514 | 0.952(493) | 0.937(138015) | 0.261 | 0.134 |
| HUNT | 0 | 0.98 | 0.518 | 0.919(284) | 0.926(66192) | –0.105 | 0.162 |
| MoBa | 0 | 0.95 | 0.00781 | 0.875(104) | 0.926(2193) | –0.713 | 0.268 |
| NORDiC-nor | 0 | 0.91 | 0.504 | 0.923(365) | 0.930(315) | –0.16 | 0.24 |
| NORDiC-swe_EGOS | 0 | 0.96 | 0.0285 | 0.939(1997) | 0.927(3943) | 0.181 | 0.0827 |
| UKBB | 1 | 0.98 | 0.332 | 0.950(776) | 0.944(125729) | 0.109 | 0.112 |
| bioVU | 0 | 0.97 | 0.204 | 0.949(1014) | 0.941(38613) | 0.125 | 0.0983 |
| IOCDF-GC CC | 0 | 0.93 | 0.00209 | 0.951(1519) | 0.937(3541) | 0.317 | 0.103 |
| OCGAS | 0 | 0.94 | 0.38 | 0.951(986) | 0.944(1023) | 0.129 | 0.147 |
| Psych_Broad | 0 | 0.94 | 0.0152 | 0.948(1396) | 0.933(4009) | 0.246 | 0.102 |
| WWF | 0 | 0.82 | 0.57 | 0.950(140) | 0.944(526) | 0.194 | 0.341 |
| **Meta - analysis** | **1** | **0.97** | **2e–07** | **0.945(14140)** | **0.937(562117)** | **0.165** | **0.0318** |

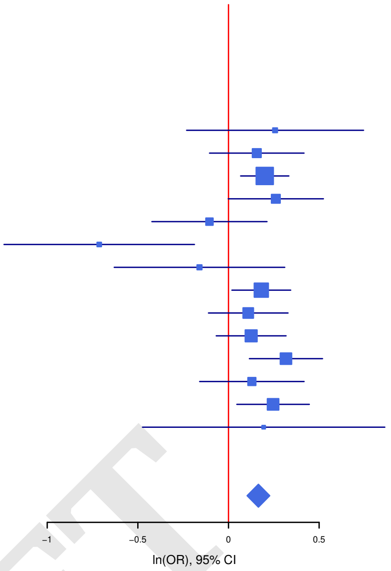


DRAFT

**Fig. S17. Forest plot of SNP rs9535127.** The plot shows the effect estimate with 95%-confidence interval for each cohort contributing to the meta-analysis and for the inverse variance weighted meta analysis. The table lists INFO (imputation score), p-value, f_ca(n) (frequency cases), f_co(n) (frequency controls), ln(OR), and STDerr (standard error) for each of the contributing cohorts and for the meta-analysis. At the top, + indicates a positive direction of effect, - a negative direction of effect while ? indicates that the SNP was not contained in the respective cohort.

| **rs9535127** | | **T/C** | | **13:31606961** | | | |
|---|---|---|---|---|---|---|---|
| | | ++?++++++?–+?+??+++ | | het_P: | 0.858 | het_I: | 0.0 |
| | **ngt** | **info** | **p_value** | **f_ca(n)** | **f_co(n)** | **ln(OR)** | **STDerr** |
| dan_daner_610_final. | 0 | 0.98 | 0.16 | 0.498(323) | 0.460(323) | 0.159 | 0.113 |
| dan_daner_AUS_refstrict_final. | 0 | 0.96 | 0.0853 | 0.496(757) | 0.471(5368) | 0.097 | 0.0563 |
| dan_daner_DOTS_final. | 0 | 0.98 | 0.0105 | 0.486(2678) | 0.467(10410) | 0.0796 | 0.0311 |
| dan_daner_EPOC_final. | 0 | 0.98 | 0.0349 | 0.545(195) | 0.472(204) | 0.301 | 0.144 |
| dan_daner_EstBB_final. | 0 | 0.99 | 0.0292 | 0.524(493) | 0.490(138015) | 0.141 | 0.0646 |
| dan_daner_FinnGen_final. | 0 | 1.00 | 0.133 | 0.541(790) | 0.526(161390) | 0.0779 | 0.0519 |
| dan_daner_HUNT_final. | 0 | 0.99 | 0.458 | 0.471(284) | 0.456(66192) | 0.0631 | 0.0851 |
| dan_daner_Nordic_norway_HRC_final. | 0 | 0.94 | 0.782 | 0.438(365) | 0.461(315) | –0.0341 | 0.123 |
| dan_daner_Nordic_sweden_HRC_final. | 0 | 0.99 | 0.0993 | 0.494(1997) | 0.481(3943) | 0.0661 | 0.0401 |
| dan_daner_UKBB_saige_final. | 1 | 0.99 | 0.609 | 0.473(776) | 0.466(125729) | 0.0263 | 0.0515 |
| dan_daner_iocd_final. | 0 | 1.02 | 0.276 | 0.470(1519) | 0.461(3541) | 0.0481 | 0.0442 |
| dan_daner_ocgas_all2_final. | 0 | 0.97 | 0.099 | 0.481(986) | 0.456(1023) | 0.106 | 0.0644 |
| dan_daner_psych_broad_final. | 0 | 1.00 | 0.386 | 0.466(1396) | 0.450(4009) | 0.0391 | 0.0451 |
| dan_daner_wue_final. | 0 | 0.93 | 0.205 | 0.497(140) | 0.461(526) | 0.18 | 0.142 |
| **dan_** | **1** | **0.99** | **5.73e–07** | **0.487(14140)** | **0.490(562117)** | **0.0738** | **0.0148** |

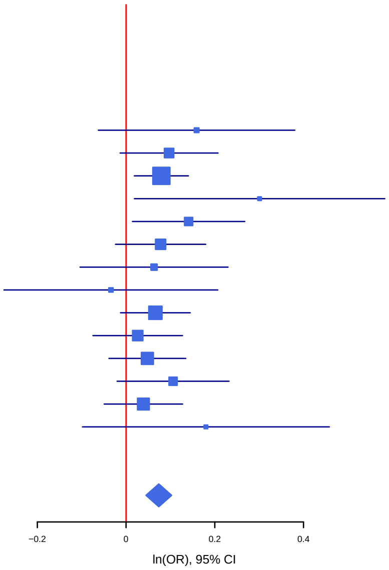

**Fig. S18. Polygenic risk score analysis in OCD samples** Variance explained in the target samples based on scores derived in the discovery samples for 8 significance thresholds ($P_T$ < 0.001, 0.05, 0.1, 0.2, 0.3, 0.4, 0.5, and 1, plotted left to right for each target sample). The y-axis indicates Nagelkerke's pseudo-$R^2$; the number above each set of bars is the best P-value for the target sample analysis. Numbers of cases and controls in each target sample are listed below each set of bars. The discovery sample is the overall PGC OCD samples minus the target sample.

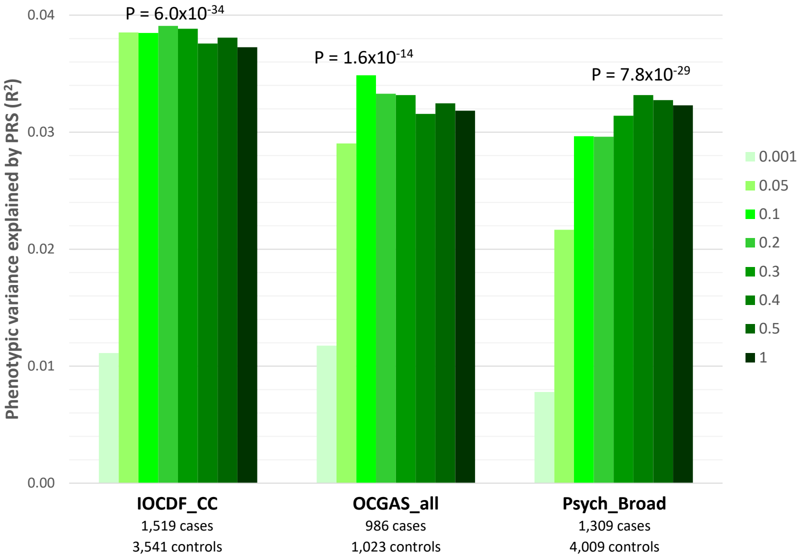


DRAFT

**Fig. S19. Gene overlap in the number of significant (Bonferroni-corrected) results for 4 gene-based methods (C-MAGMA, H-MAGMA, E-MAGMA, and S-PrediXcan).** P value thresholds: C-MAGMA: $2.50 \times 10^{-6}$ (N=20,031 tests); E-MAGMA: P<$2.51 \times 10^{-7}$ (N=199,421 tests); H-MAGMA: P<$2.05 \times 10^{-6}$ (N=24,358 tests); S-MultiXcan: P<$2.31 \times 10^{-6}$ (N=21,601 tests).

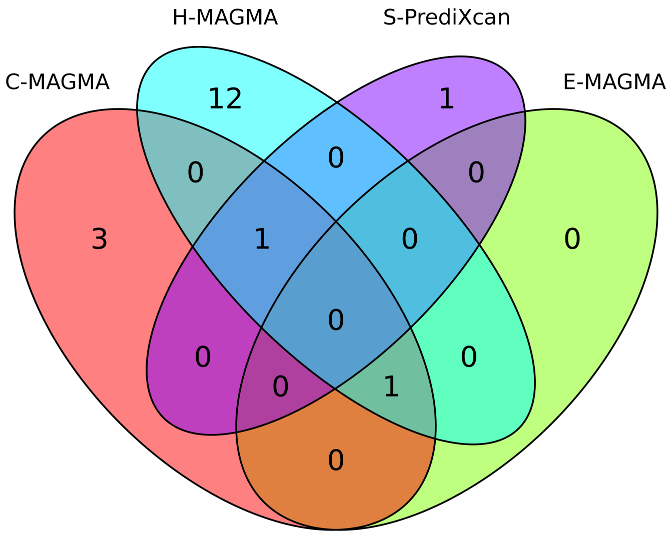

**Fig. S20. Regional plot of the top lead SNP (rs2581789) and nearest genes.** A) Zoomed in regional plot with, from the top, GWAS p values, Combined Annotation Dependent Depletion (CADD) score, RegulomeDB score and expression quantitative trait loci (eQTL) -log10 p values. SNPs in genomic risk loci are color-coded as a function of their maximum r² to one of the independent significant SNPs in the locus. SNPs that are not in LD with any of the independent significant SNPs (r²<=0.2) are grey. Only SNPs with P < 0.05 are displayed. eQTLs are plotted per gene and colored based on tissue type. In the plots of CADD score, RegulomeDB score and eQTLS, unmapped SNPs are displayed in gray. B) Is the extension of A. The top part with zoomed-in regional plot with GWAS p-values is the same as in A), followed by further eQTL -log10 p values. Colour coding is the same as in A).

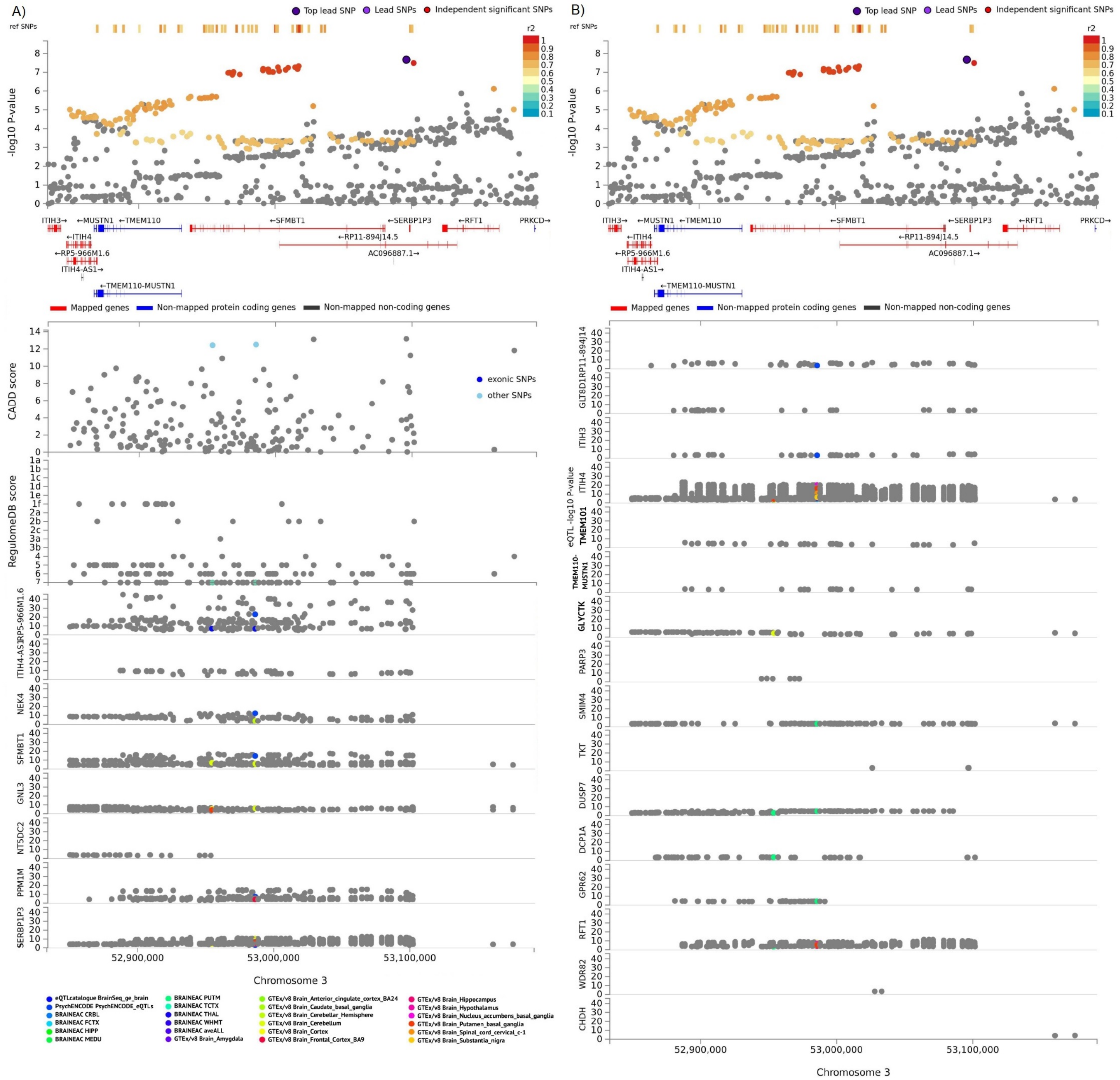

**Fig. S21. FUMA GTEX expression heatmap** displaying the average expression per tissue type per OCD associated gene (except for LOC101928274 due to lack of matching gene ID) following winsorization at 50 and log 2 transformation with pseudocount 1. The expression value is given in TPM (Transcripts Per Million). Cells filled in red represent higher expression compared to cells filled in blue across genes and labels. They are ordered by hierarchical clustering.

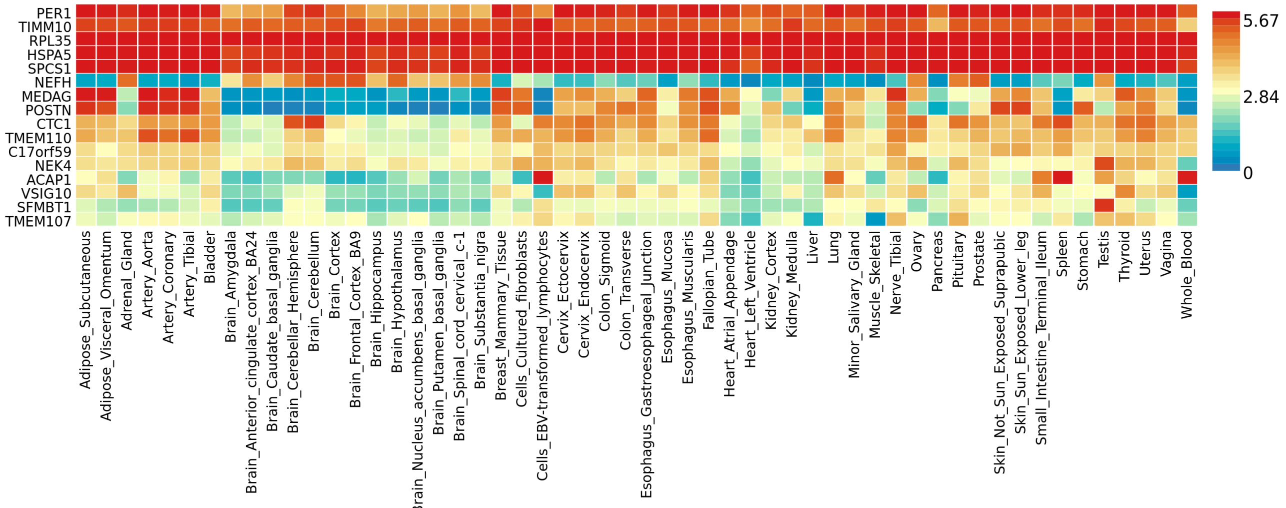


**Fig. S22. FUMA Brain Span gene expression data**: Expression heatmap displaying the average expression for 11 general developmental stages per OCD associated gene (except for LOC101928274 due to lack of matching gene ID) following winsorization at 50 and log 2 transformation with pseudocount 1. The expression value is given in TPM RPKM (Read Per Kilobase per Million). Cells filled in red represent higher expression compared to cells filled in blue across genes and labels. They are ordered by hierarchical clustering.

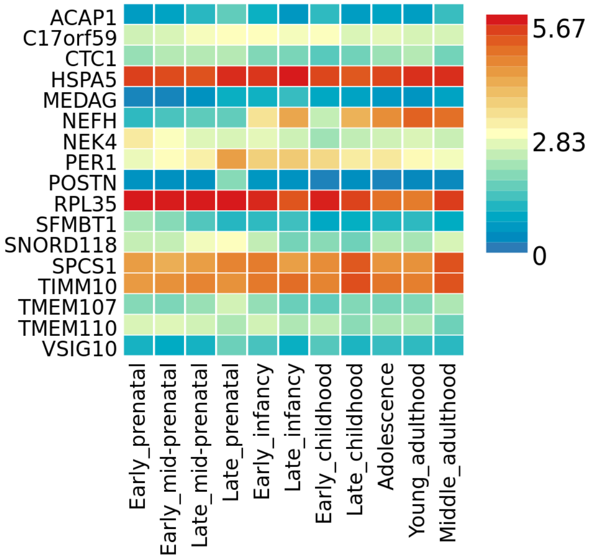

**Fig. S23. FUMA Brain Span gene expression data**: Expression heatmap displaying the average expression for 29 different ages per OCD associated gene (except for LOC101928274 due to lack of matching gene ID) following winsorization at 50 and log 2 transformation with pseudocount 1. The expression value is given in TPM RPKM (Read Per Kilobase per Million). Cells filled in red represent higher expression compared to cells filled in blue across genes and labels. They are ordered by hierarchical clustering.

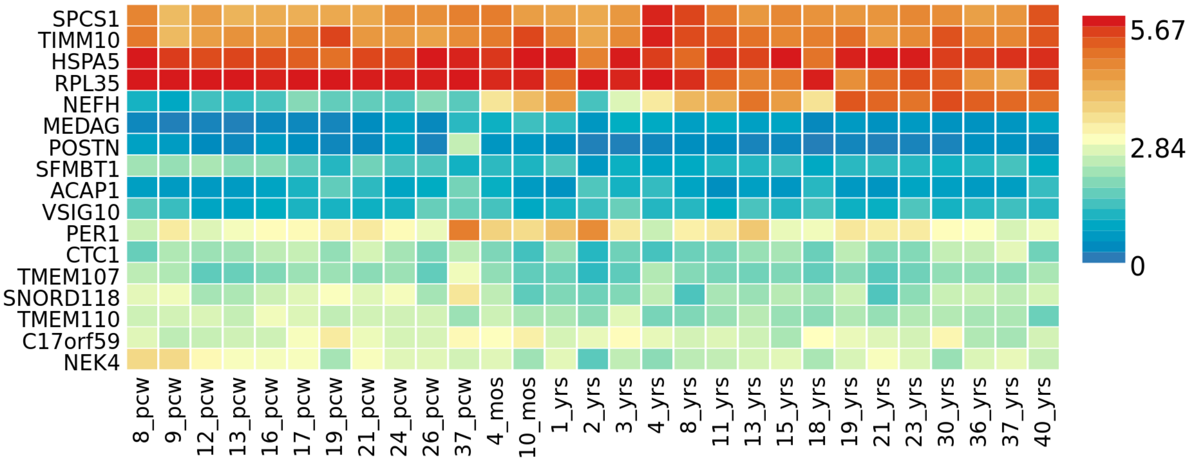

**Fig. S24. FUMA DEG GTEX gene expression data- both-sided-, up- or down regulation.** Differentially Expressed Gene (DEG) sets were pre-calculated by performing two-sided t-tests for any one of the labels against all others. Genes with p value 0.05 after Bonferroni correction and absolute log fold change 0.58 were defined as differentially expressed genes in a given label compared to others. Tissues are ordered by two-sided DEG P-value. No associated sets were observed for GTEx v8 54 tissues. On top of DEG, up-regulated DEG and down-regulated DEG were also pre-calculated by taking the sign of t-statistics into account. Input genes were tested against each of the DEG sets using the hypergeometric test. The background genes are genes that have average expression value > 1 in at least one of the labels and exist in the user selected background genes. Significant enrichment at Bonferroni corrected P-value 0.05 are coloured in red. Bonferroni correction was performed for each of both-sided, up- and down-regulated DEG sets separately.

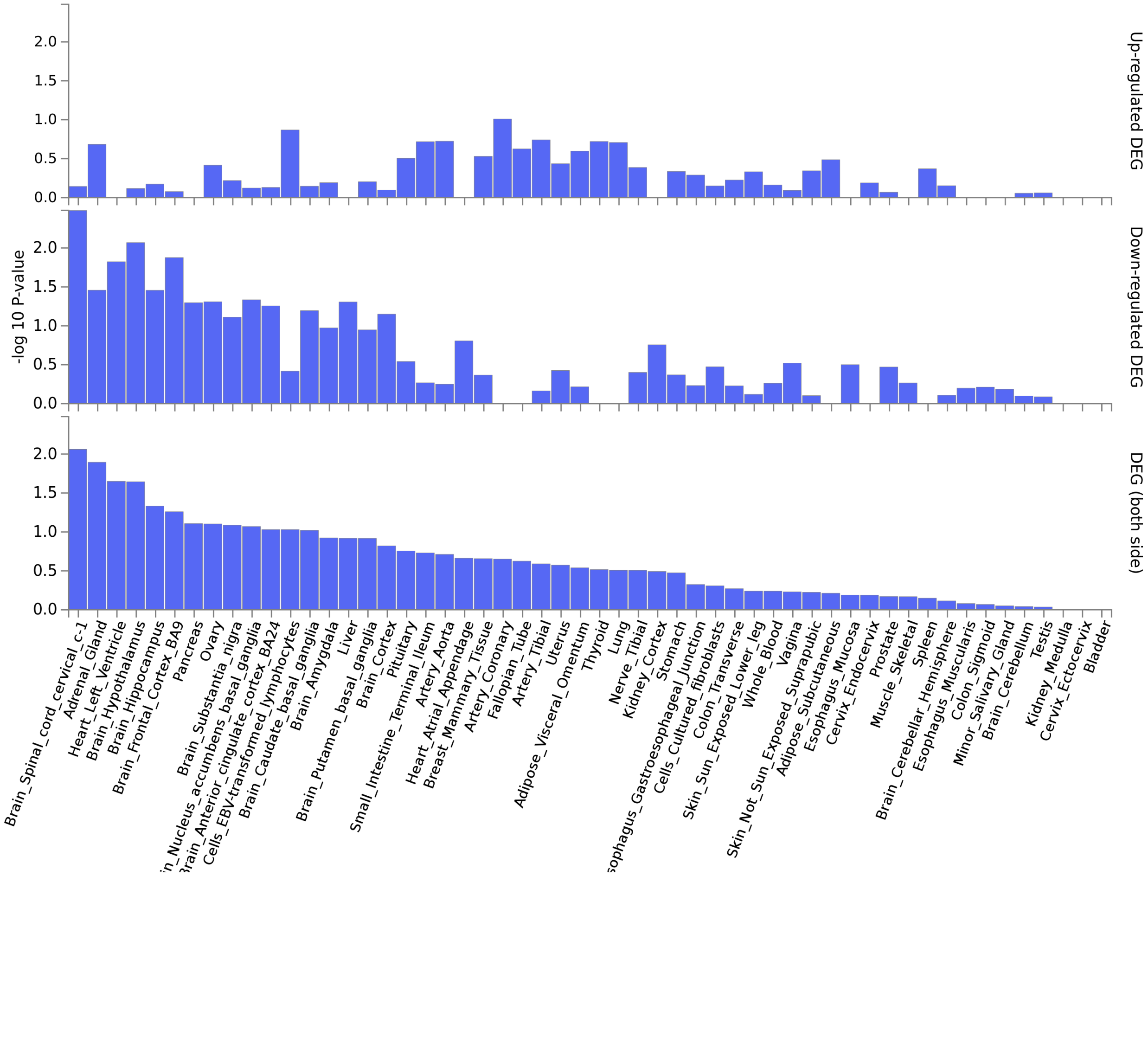

**Fig. S25. FUMA DEG GTEX gene expression data- both-sided-, up- or down regulation.** Differentially Expressed Gene (DEG) sets were pre-calculated by performing two-sided t-tests for any one of the labels against all others. Genes with p value  0.05 after Bonferroni correction and absolute log fold change  0.58 were defined as differentially expressed genes in a given label compared to others. Tissues are ordered by two-sided DEG P-value. No associated sets were observed for 11 general developmental stages in the Brainspan data. On top of DEG, up-regulated DEG and down-regulated DEG were also pre-calculated by taking the sign of t-statistics into account. Input genes were tested against each of the DEG sets using the hypergeometric test. The background genes are genes that have average expression value > 1 in at least one of the labels and exist in the user selected background genes. Significant enrichment at Bonferroni corrected P-value  0.05 are coloured in red. Bonferroni correction was performed for each of both-sided, up- and down-regulated DEG sets separately.

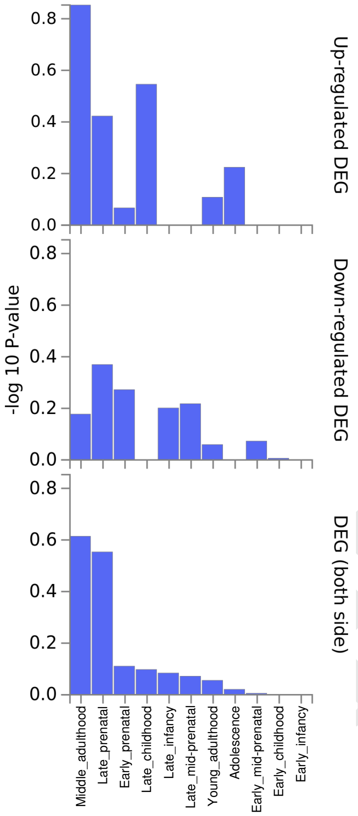


DRAFT

**Fig. S26. FUMA DEG GTEX gene expression data- both-sided-, up- or down regulation.** Differentially Expressed Gene (DEG) sets were pre-calculated by performing two-sided t-tests for any one of the labels against all others. Genes with p value  0.05 after Bonferroni correction and absolute log fold change  0.58 were defined as differentially expressed genes in a given label compared to others. Tissues are ordered by two-sided DEG P-value. No associated sets were observed for 29 different ages of brain samples in the Brainspan. On top of DEG, up-regulated DEG and down-regulated DEG were also pre-calculated by taking the sign of t-statistics into account. Input genes were tested against each of the DEG sets using the hypergeometric test. The background genes are genes that have average expression value > 1 in at least one of the labels and exist in the user selected background genes. Significant enrichment at Bonferroni corrected P-value  0.05 are coloured in red. Bonferroni correction was performed for each of both-sided, up- and down-regulated DEG sets separately.

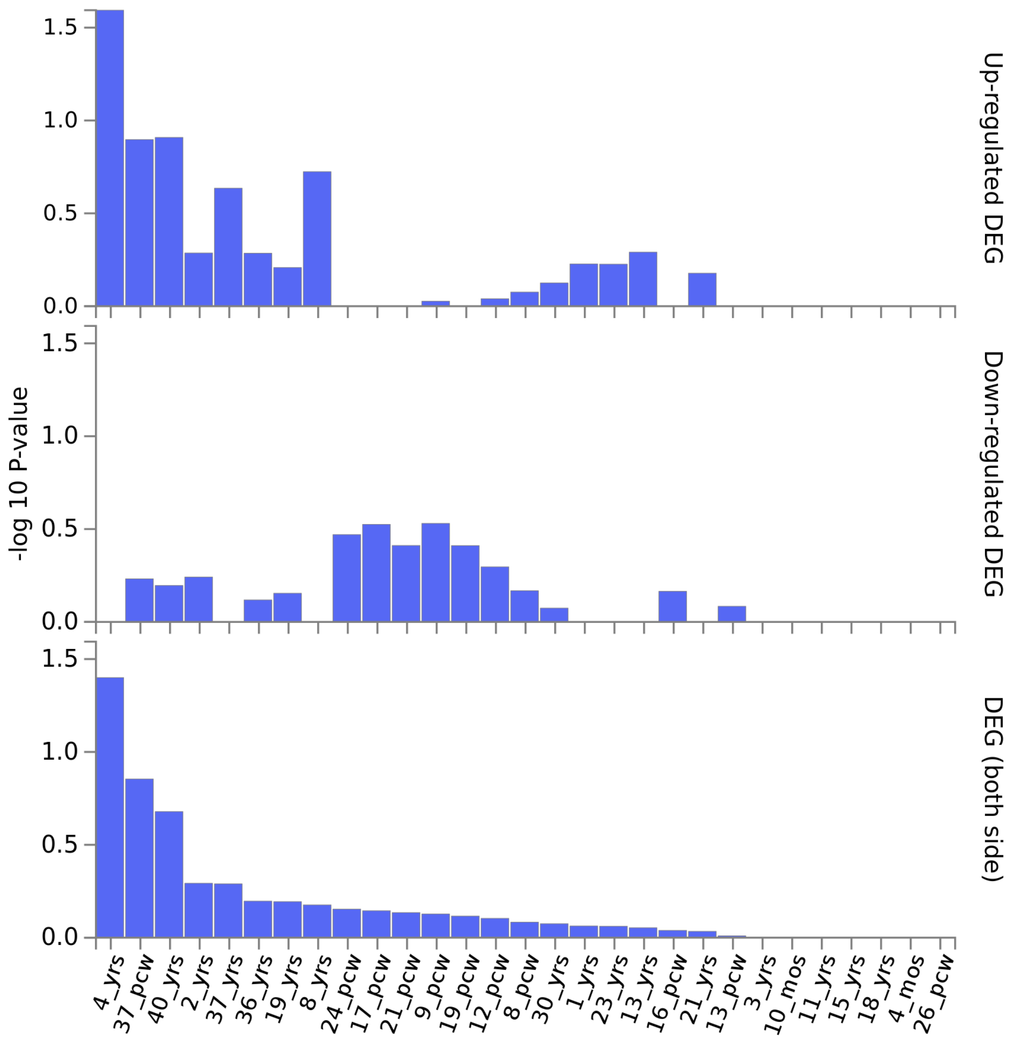

**Fig. S27. Mouse tissue / broad celltype enrichment analysis.** Depicted are the results for enrichment analyses of the OCD summary statistics in regions of genes that are preferentially expressed in the test tissue / broad celltype group. The -log10 p values shown are the mean of the -log10 p values resulting from LDSC-analysis and from MAGMA-analysis. In general, tissues / celltype groups are only considered significant if their p-values survive FDR correction in both the LDSC and MAGMA analyses. Based on this criterium we fail to detect an individual celltype or broad celltype group from the mouse nervous system with a significant enrichment of OCD genetic risk.

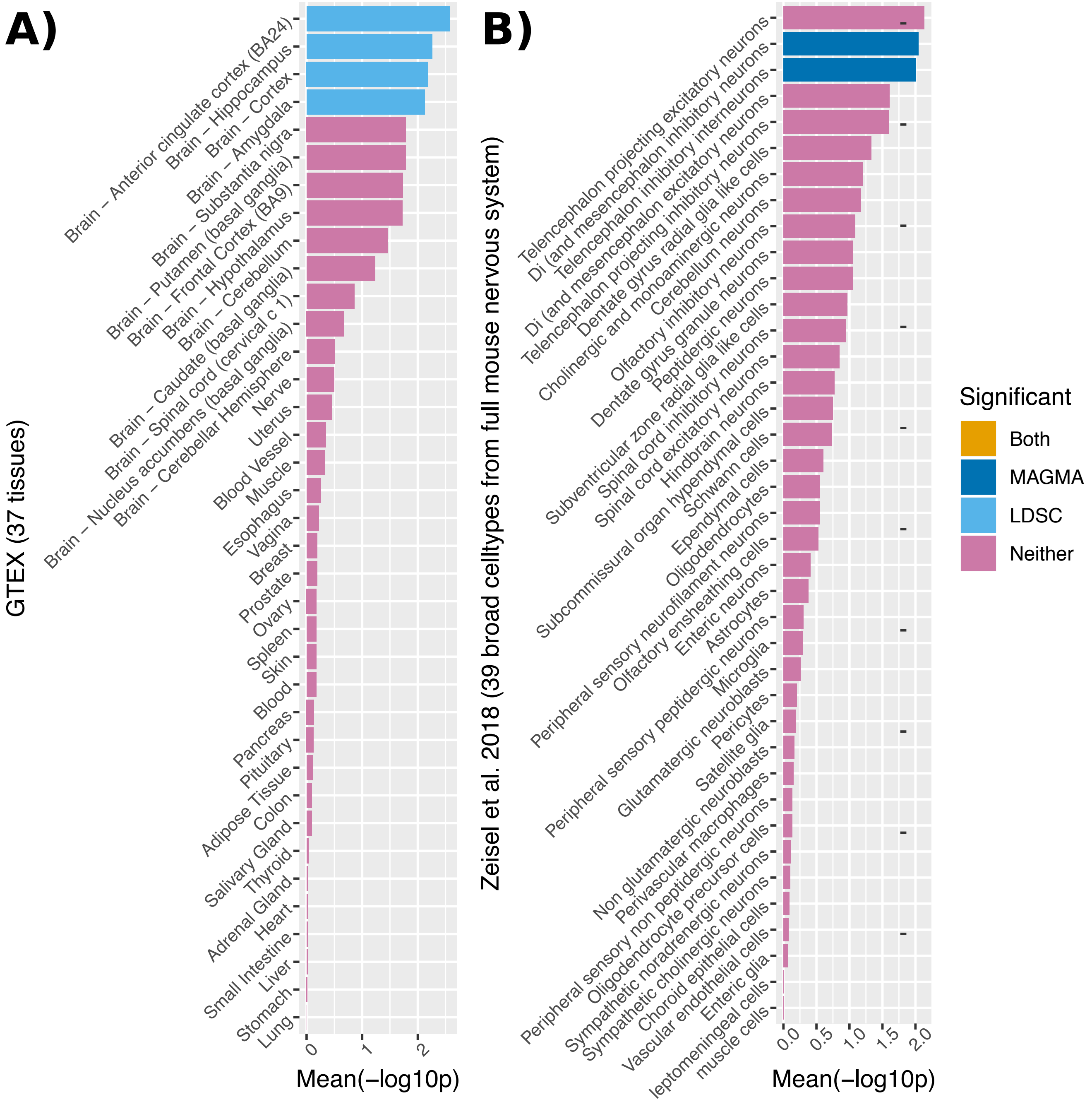

**Fig. S28. Specific celltype enrichment analysis in mouse nervous system.** The results from enrichment analyses across the entire mouse nervous system using MAGMA are depicted (265 celltypes total). A result is considered significant if the FDR-adjusted p-value is less than 0.05. We observe a total of 9 individual celltypes that are significant based on these requirements.

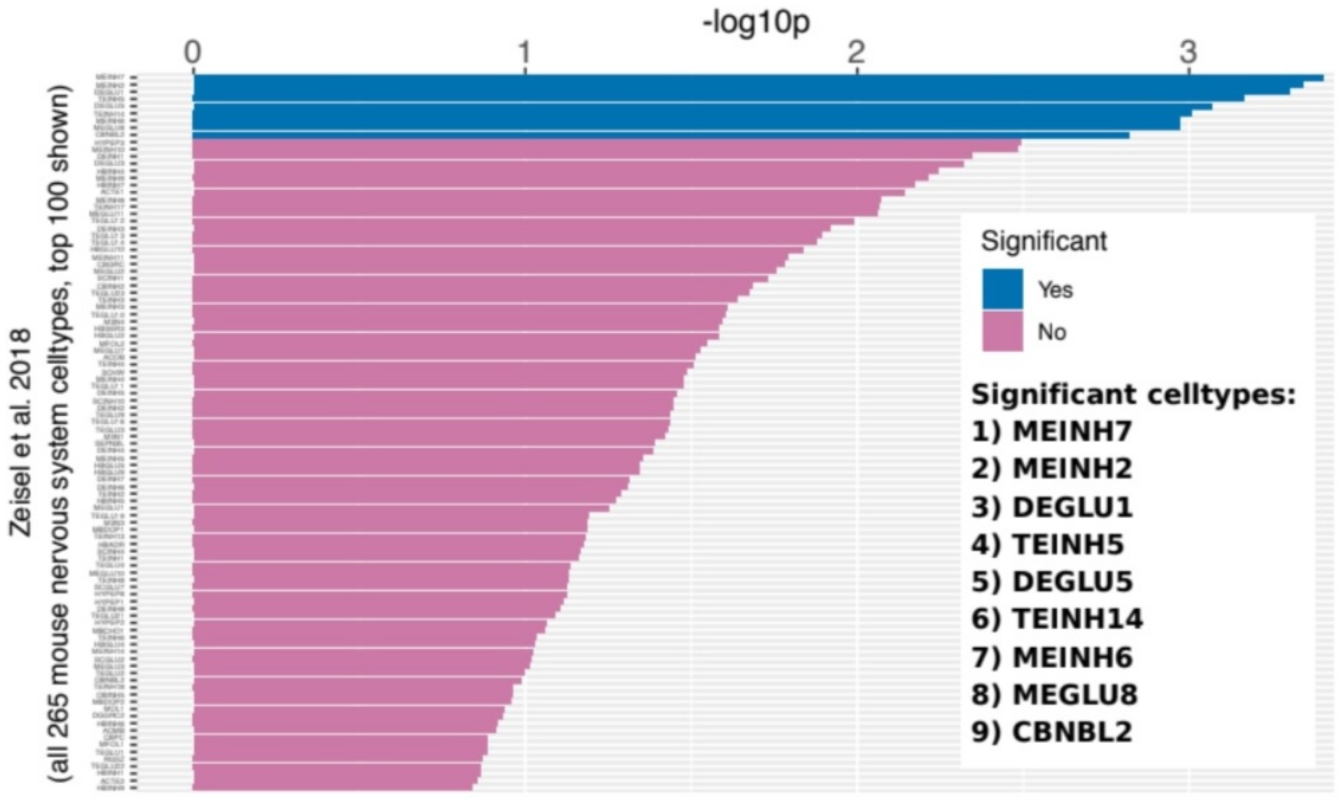

**Fig. S29. Overlap of genome-wide significant locus with brain-derived chromatin interactions.** The webserver FUMA was used to assess and plot the overlap of the genome-wide significant locus detected in this study with high confidence chromatin interactions (orange) detected in the psychENCODE study and Giusti-Rodriguez et al. No genes that were linked to the locus via these interactions had a clear obvious role in OCD.

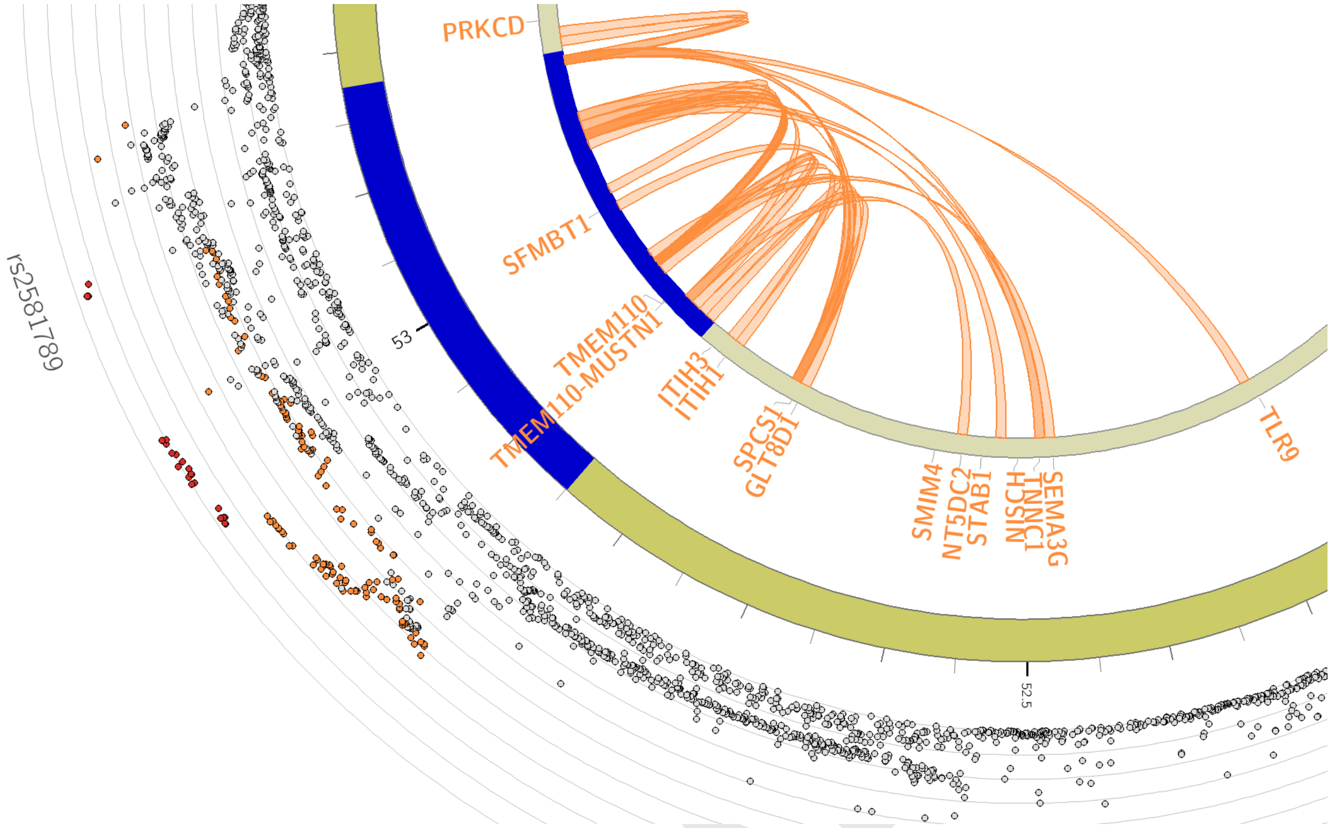


DRAFT

**Fig. S30. Genetic correlations (rg) between three OCD datasets and a broad range (N=82) of other phenotypes.** Light green indicates rg with OCD excluding biobanks, blue indicates rg with OCD excluding the Australian dataset and the danish iPSYCH dataset as they were not primarily ascertained for OCD, and black indicates rg with OCD datasets from biobanks. Error bars represent 95% confidence intervals and asterisks indicate significant associations after FDR correction for multiple testing. Non-significant correlations with SE $>$ 0.5 are excluded from display.

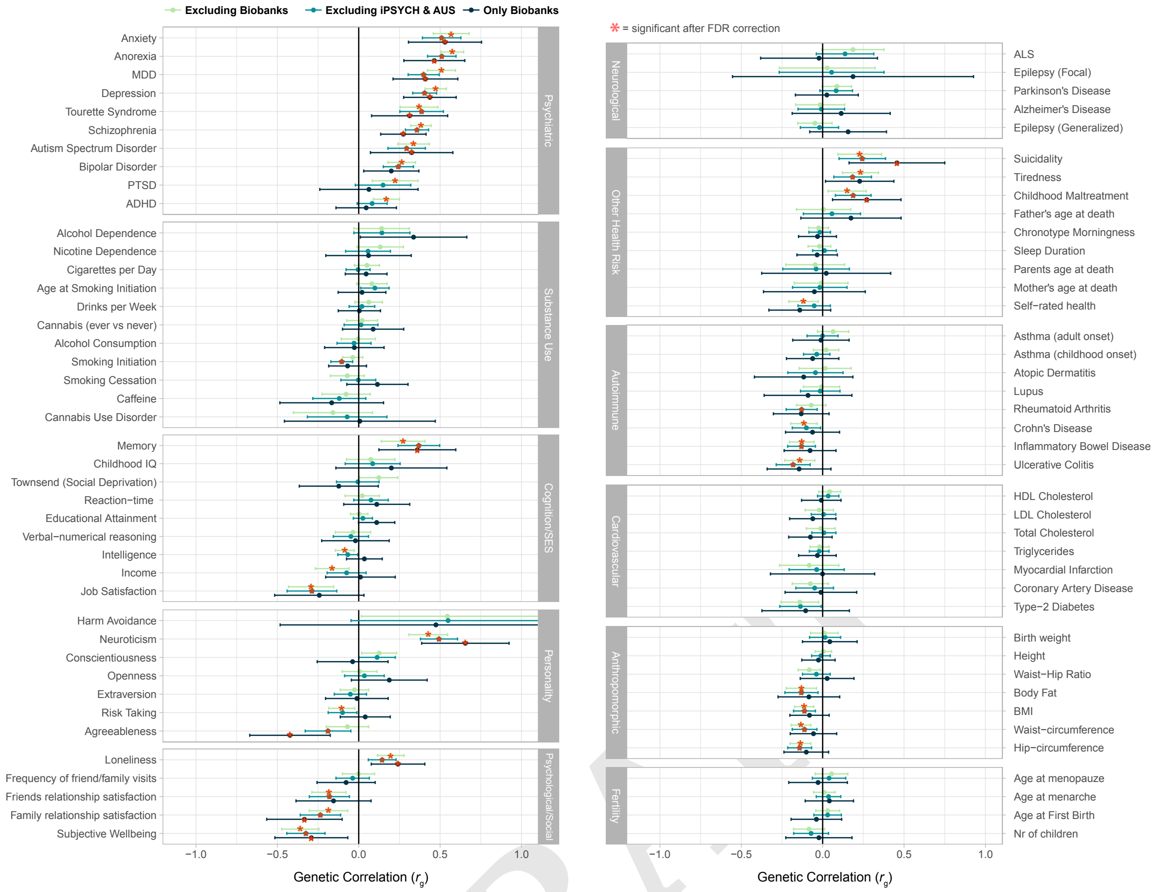


DRAFT


DRAFT
